# SARS-CoV-2 infection in Africa: A systematic review and meta-analysis of standardised seroprevalence studies, from January 2020 to December 2021

**DOI:** 10.1101/2022.02.14.22270934

**Authors:** HC Lewis, H Ware, M Whelan, L Subissi, Z Li, X Ma, A Nardone, M Valenciano, B Cheng, K Noel, C Cao, M Yanes-Lane, B Herring, A Talisuna, N Nsenga, T Balde, DA Clifton, M Van Kerkhove, DL Buckeridge, N Bobrovitz, J Okeibunor, RK Arora, I Bergeri, the UNITY Studies Collaborator Group

**Affiliations:** World Health Organization, Regional Office for Africa, Brazzaville, Congo; World Health Organization, Geneva, Switzerland; Centre for Health Informatics, Cumming School of Medicine, University of Calgary, Canada; Faculty of Engineering, University of Waterloo, Canada; Institute of Health Policy Management and Evaluation, University of Toronto, Toronto, ON, Canada; Epiconcept, Paris, France; School of Population and Global Health, McGill University, Montreal, Quebec, Canada; Department of Epidemiology, Biostatistics and Occupational Health, McGill University, Montreal, Canada; COVID-19 Immunity Task Force Secretariat, McGill University, Montreal, Canad; Temerty Faculty of Medicine, University of Toronto, Toronto, Ontario, Canada; Institute of Biomedical Engineering, University of Oxford, UK

## Abstract

**Introduction:** Estimating COVID-19 cumulative incidence in Africa remains problematic due to challenges in contact tracing, routine surveillance systems and laboratory testing capacities and strategies. We undertook a meta-analysis of population-based seroprevalence studies to estimate SARS-CoV-2 seroprevalence in Africa to inform evidence-based decision making on Public Health and Social Measures (PHSM) and vaccine strategy.

**Methods:** We searched for seroprevalence studies conducted in Africa published 01-01-2020 to 30-12-2021 in Medline, Embase, Web of Science, and Europe PMC (preprints), grey literature, media releases and early results from WHO Unity studies. All studies were screened, extracted, assessed for risk of bias and evaluated for alignment with the WHO Unity protocol for seroepidemiological investigations. We conducted descriptive analyses of seroprevalence and meta-analysed seroprevalence differences by demographic groups, place and time. We estimated the extent of undetected infections by comparing seroprevalence and cumulative incidence of confirmed cases reported to WHO. PROSPERO: CRD42020183634.

**Results:** We identified 54 full texts or early results, reporting 151 distinct seroprevalence studies in Africa Of these, 95 (63%) were low/moderate risk of bias studies. SARS-CoV-2 seroprevalence rose from 3.0% [95% CI: 1.0-9.2%] in Q2 2020 to 65.1% [95% CI: 56.3-73.0%] in Q3 2021. The ratios of seroprevalence from infection to cumulative incidence of confirmed cases was large (overall: 97:1, ranging from 10:1 to 958:1) and steady over time. Seroprevalence was highly heterogeneous both within countries - urban vs. rural (lower seroprevalence for rural geographic areas), children vs. adults (children aged 0-9 years had the lowest seroprevalence) - and between countries and African sub-regions (Middle, Western and Eastern Africa associated with higher seroprevalence).

**Conclusion:** We report high seroprevalence in Africa suggesting greater population exposure to SARS-CoV-2 and protection against COVID-19 disease than indicated by surveillance data. As seroprevalence was heterogeneous, targeted PHSM and vaccination strategies need to be tailored to local epidemiological situations.

## Introduction

Africa is experiencing unprecedented challenges due to the Coronavirus disease 2019 (COVID-19) pandemic, both directly and from the compounding effects of other health, economic and social factors.[1,2] As of 9 February 2022, 11.1 million COVID-19 cases and 238,845 deaths were confirmed on the African Continent.[3] To date, the pandemic has progressed in four main waves, with the most recent wave largely due to the Omicron variant circulating in a number of countries. However, reported data indicates a less severe disease profile in Africa compared to other regions globally: fewer cases, proportionally fewer patients with severe outcomes and death, and proportionally more asymptomatic cases.[3–5]

Precisely estimating COVID-19 cumulative incidence in Africa remains problematic due to challenges in contact tracing, routine surveillance systems and laboratory testing capacities and strategies in many countries. Furthermore, Africa is a large, complex and heterogeneous continent with a range of different economies, countries impacted by humanitarian crises, vulnerable population groups and unique public health challenges. With low vaccine coverage in Africa (17% as of 9 February 2022),[6] using seroprevalence data to understand the true dynamics of SARS-CoV-2 infection and case under-ascertainment is key to map the extent of protection in the general population and inform the public health response.

SARS-CoV-2 seroprevalence studies in Africa have been under-represented in previous systematic reviews and meta-analyses due to sparse seroprevalence data. The one previous meta-analysis in Africa [7] pooled results from studies published to April 2021 which included only a small number of studies (n=23 studies) with limited geographical coverage and scope (no nationwide studies). Seroprevalence data are now emerging from completed field investigations, in part enabled by the support of the World Health Organization’s UNITY Studies. At the start of the pandemic, UNITY Studies developed a population-based, age-stratified seroepidemiological investigation protocol (SEROPREV) and supported countries to plan and implement robust and standardised seroprevalence studies, and to analyse and publish their results.[8] These studies provide us with the opportunity to understand SARS-CoV-2 epidemiology in Africa despite low vaccination coverage and case under-ascertainment. To note, our investigative team has also conducted an analysis of SEROPREV studies globally, including regional comparison, which serves as the foundational source for the methods and analyses further developed in this paper [9] to provide more detailed analyses for Africa.

We aimed to describe the COVID-19 pandemic in Africa up to and before the emergence of the Omicron variant. We undertook a meta-analysis of population-based seroprevalence studies that were aligned with WHO’s standardised SEROPREV protocol to estimate SARS-CoV-2 seroprevalence in Africa to inform evidence-based decision making on Public Health and Social Measures (PHSM) and vaccine strategy. Our primary objectives were to: (i) estimate SARS-CoV-2 seroprevalence and changes over time in Africa as a whole and by UN sub-regions (ii) estimate the extent of undetected infections by comparing seroprevalence and cumulative incidence of confirmed cases and (iii) identify heterogeneity in seroprevalence attributable to demographic factors, country-level factors, and study design.

## Methods

### Search strategy, selection criteria, and data extraction

We conducted a systematic review of seroprevalence data sources in the African continent published from 1 January 2020 to 30 December 2021. We searched MEDLINE, Embase, Web of Science, and Europe PMC for published articles, preprints, grey literature, and media reports. We also accepted early results from ongoing studies in Africa shared by study teams with WHO via a standardised template [10] up to 20 January 2022, which have been made available on an open-access repository.[11] This review is registered with PROSPERO (CRD42020183634) where further details on our search strategy, inclusion criteria, screening and extraction protocol are available.[12]

Full details on our inclusion and exclusion criteria are described elsewhere.[9] Briefly, we included SARS-CoV-2 seroprevalence studies in the general population aligned with the WHO SEROPREV protocol [8,9] conducted on the African Continent. We included cohort and cross-sectional study designs. We also specified assay performance criteria for inclusion of at least 90% sensitivity and 97% specificity determined by the manufacturer. Exceptions to the assay performance criteria were made to accommodate resource limitations in COVID-19 emergency humanitarian settings (herein referred to as HRP settings or status, defined by the Global COVID-19 Humanitarian Response Plan).[13] We excluded studies without a clear numerator or denominator; without study dates and seroprevalence estimate; studies sampling closed populations (prisons, schools, etc); and studies that excluded participants with previous COVID-19 diagnosis or vaccination. Sources identified through our search and results submitted directly to us through the Unity Studies Initiative were screened using the same selection criteria.

Screening was conducted in two stages by two independent reviewers: title and abstract screening, followed by full text screening. Conflicts were resolved by a third reviewer. Articles selected for inclusion were then extracted and verified by two reviewers. In cases where sources contained multiple primary estimates of seroprevalence (i.e. non-overlapping populations, separate methodologies, etc), the source (full text) was split into multiple individual studies for extraction. Reviewers collected information on the source’s key characteristics (journal/venue, publication date), study information (demographics, sampling criteria, testing strategy) as well as seroprevalence estimates.

### Risk of bias assessment

All studies were critically appraised using a modified Joanna Briggs Institute (JBI) 9-point checklist for seroprevalence studies. [14–16] Each study’s JBI rating was completed by two independent reviewers. Conflicts were resolved by consensus. An automated decision rule based on combinations of each reviewer’s JBI checklist was applied to determine overall study risk of bias (low, moderate, or high). This automated decision rule is described in detail elsewhere. [17]

### Data synthesis and analysis

Seroprevalence studies were classified by study design, sample frame, sampling method, type of serological assay, and geographic scope (local, sub-national, or national) (Table S1). We further classified local scope into national capitals and other cities or towns over or under 300,000 inhabitants (Table S2), based on urban agglomerations defined by the UN.[18] Countries were classified according to the UN’s African sub-regions,[19] HRP status,[13] and World Bank income level. Each round of cohort or repeated cross-sectional studies were classified as separate studies. Where there were multiple estimates per study unrelated to time, estimates were prioritised based on adjustment, antibody isotypes, test type, and antibody targets (full details: Supplement S3.1). To best reflect the period covered by the estimate, we anchored each estimate to the date halfway between start of sampling and end (“sampling midpoint date”). We also identified studies with seroprevalence estimates by urban and rural areas within cities or towns (as defined by study authors), males and females, and 10-year age groups up to 60 years and over. Data was analysed using R statistical software version 4.1.2.[20] There was no public or patient involvement in this research.

We summarised the characteristics of all identified studies (Dataset 0). We used studies rated low or moderate risk of bias to estimate seroprevalence in the general population over time and conduct subgroup meta-analysis and meta-regression (Dataset 1) and national studies rated low or moderate risk of bias to calculate seroprevalence to cumulative incidence ratios (Dataset 2). We summarised the proportion of seropositives in each individual study (Dataset 1) by country and over time, highlighting the scope of the result and which results derived from the same cohort or repeated cross-sectional data sources. We summarised other relevant variables in each country, including smoothed daily cases, cumulative incidence of cases reported to WHO,[3] and relative variant genome frequency shared via the GISAID initiative.[21] To estimate combined seroprevalence from infection or vaccination in the general population, we meta-analysed reported seroprevalence by UN sub-region and quarter to reduce heterogeneity between studies, pooling studies using a random-effects model (metaprop in R). We used the Clopper-Pearson method to produce 95% confidence intervals reflecting uncertainty.[22,23] To estimate seroprevalence attributable to infection only, we adjusted anti-Spike reported seroprevalence using a standard formula before pooling studies.[24]

We also estimated the magnitude by which confirmed SARS-CoV-2 cases underestimated the true burden of disease. To do so, we applied our overall meta-analysis estimates of seroprevalence from infection to the African population to estimate true infections, and divided them by the number of laboratory confirmed cases at the time.[3] To examine differences by country, we calculated the ratio between estimated seroprevalence from infection and the cumulative incidence of COVID-19 cases for each national study in Dataset 2. Finally, to put the ratios into context of different health systems across countries, we calculated Pearson’s correlation coefficient between the under-ascertainment ratios in each study and four indices of national health system functionality (access, quality, demand, and resilience).[25] To estimate asymptomatic seroprevalence, we summarised the proportion of seropositives that reported no COVID-19 symptoms during the study period in the aggregated results shared by UNITY collaborators. We tested for differences in the distribution across age and sex groups using the Kruskal-Wallis H-test.

To explore possible causes of heterogeneity among study results, we conducted subgroup analysis and meta-regression. First, to quantify demographic differences in seroprevalence, we calculated the seroprevalence ratio between subgroups within each study with available data in Dataset 1. We compared each 10-year age group to adults 20-29, males to females, and, within local studies, urban to rural areas. To produce summary estimates, we aggregated the ratios across studies using inverse variance-weighted meta-analysis. Heterogeneity was quantified using the I^2^ statistic. We also constructed a Poisson generalised linear mixed-effects regression to explore associations between seroprevalence and study and country factors.[26] Independent categorical predictors were selected *a priori* as UN sub-region, sample frame, geographic scope, low or high population density in the study setting (thresholds and sources in Supplement S3.2), and type of serological test. Cumulative incidence of confirmed cases was selected as an independent continuous predictor.

Our main analysis used seroprevalence estimates uncorrected for test characteristics. As a sensitivity analysis, we also produced results adjusting for test characteristics through Bayesian measurement error models, with binomial sensitivity and specificity distributions. The sensitivity and specificity values for correction were prioritised from the WHO SARS-CoV-2 Test Kit Comparative Study conducted at the NRL Australia,[27] followed by a multicentre evaluation of 47 commercial SARS-CoV-2 immunoassays by 41 Dutch laboratories,[28] and from independent evaluations by study authors where author-designed assays were used.

## Results

### Selection of studies

We identified 73,348 titles and abstracts in our search. Of these, 4,221 full text articles were included in full text screening. We identified 54 data sources reporting studies aligned with the SEROPREV protocol in the African Continent, 42 published and 12 aggregated results from collaborators, which contained a total of 151 unique seroprevalence studies (detailed information on each study: Supplement S4.2, Tables S5-S7; references: Supplement S4.4).

### Study characteristics and quality assessment

The 151 identified studies represented 41% (22/54) of WHO African Continent Member States (MS), which make up 71% of the continent’s population (957 million/1.3 billion inhabitants). The data included 9 of 18 MS in Eastern Africa, 1 of 5 MS in Southern Africa (South Africa), 6 of 16 MS in Western Africa, 5 of 9 MS in Middle Africa, 1 of 6 MS in Northern Africa (Egypt), and 55% (n=16/29) vulnerable HRP countries. Early results from UNITY study collaborators in nine countries made up over one third (34%, n=51/151) of identified studies.

Among the 151 studies included in the descriptive analysis (Table 1, column 1), 25% (n=38) reported results at a national level and 7% at a sub-national level. The remaining two-thirds of studies reported results at a local level, with 25% in the national capital, 7% in another major city over 300,000 inhabitants, and 34% in a smaller city or town under 300,000 inhabitants. Of studies reporting results at a local level, 36% indicated the city was selected because of suspected high SARS-CoV-2 transmission, 62% mentioned convenience, and 65% mentioned high population density.

**Table 1:** Study characteristics.

|  | **Dataset 0: ALL STUDIES** | **Dataset 1: LOW AND MODERATE RISK OF BIAS STUDIES** | **Dataset 2: LOW AND MODERATE RISK OF BIAS STUDIES; NATIONAL SCOPE** |
| --- | --- | --- | --- |
|  | **Used in descriptive analysis** | **Used to estimate seroprevalence over time and identify associated factors** | **Used to estimate ascertainment** |
| **Study Characteristic** | **N = 151<sup>1</sup>** | **N = 95<sup>1</sup>** | **N = 36<sup>1</sup>** |
| **World Bank income level** |  |  |  |
| Low-income country | 95 (63%) | 53 (56%) | 23 (64%) |
| Lower-middle-income country | 33 (22%) | 22 (23%) | 11 (31%) |
| Upper-middle-income country | 23 (15%) | 20 (21%) | 2 (5.6%) |
| High-income country | 0 (0%) | 0 (0%) | 0 (0%) |
| **Humanitarian response plan (HRP)** |  |  |  |
| Vulnerable HRP country | 62 (41%) | 50 (53%) | 9 (25%) |
| **UN sub-region** |  |  |  |
| Eastern Africa | 103 (68%) | 54 (57%) | 26 (72%) |
| Southern Africa | 22 (15%) | 20 (21%) | 2 (5.6%) |
| Western Africa | 17 (11%) | 16 (17%) | 7 (19%) |
| Middle Africa | 7 (4.6%) | 5 (5.3%) | 1 (2.8%) |
| Northern Africa | 2 (1.3%) | 0 (0%) | 0 (0%) |
| **Source type** |  |  |  |
| Journal Article (Peer-Reviewed) | 71 (47%) | 30 (32%) | 5 (14%) |
| Preprint | 13 (8.6%) | 8 (8.4%) | 4 (11%) |
| Presentation, Conference, or Institutional Report | 16 (11%) | 7 (7.4%) | 0 (0%) |
| Early results from UNITY study collaborators | 51 (34%) | 50 (53%) | 27 (75%) |
| **Geographic scope** |  |  |  |
| National | 38 (25%) | 36 (38%) | 36 (100%) |
| Subnational | 10 (6.6%) | 9 (9.5%) | 0 (0%) |
| Capital city | 37 (25%) | 15 (16%) | 0 (0%) |
| Other city or town > 300,000 people | 11 (7.3%) | 4 (4.2%) | 0 (0%) |
| Other city or town < 300,000 people | 51 (34%) | 28 (29%) | 0 (0%) |
| Multiple cities or towns | 4 (2.6%) | 3 (3.2%) | 0 (0%) |
| **Study population** |  |  |  |
| Blood donors | 68 (45%) | 22 (23%) | 22 (61%) |
| Residual sera | 9 (6.0%) | 4 (4.2%) | 2 (5.6%) |
| Household and community samples | 68 (45%) | 68 (72%) | 12 (33%) |
| Pregnant or parturient women | 6 (4.0%) | 1 (1.1%) | 0 (0%) |
| **Sampling method** |  |  |  |
| Convenience | 59 (39%) | 18 (19%) | 3 (8.3%) |
| Probability | 76 (50%) | 75 (79%) | 31 (86%) |
| Sequential | 16 (11%) | 2 (2.1%) | 2 (5.6%) |
| **Study design** |  |  |  |
| Cross-sectional survey | 40 (26%) | 33 (35%) | 11 (31%) |
| Prospective cohort | 19 (13%) | 19 (20%) | 0 (0%) |
| Repeated cross-sectional study | 90 (60%) | 43 (45%) | 25 (69%) |
| Retrospective cohort | 2 (1.3%) | 0 (0%) | 0 (0%) |
| **Serological test type** |  |  |  |
| CLIA | 37 (25%) | 34 (36%) | 2 (5.6%) |
| WHO-standardised SARS-CoV-2<br>Total Ab ELISA | 41 (27%) | 33 (35%) | 28 (78%) |
| Other ELISA | 50 (33%) | 7 (7.4%) | 4 (11%) |
| LFIA | 16 (11%) | 15 (16%) | 2 (5.6%) |
| Multiple Types | 5 (3.3%) | 4 (4.2%) | 0 (0%) |
| Other | 2 (1.3%) | 2 (2.1%) | 0 (0%) |
| **Overall risk of bias** |  |  |  |
| Low | 35 (23%) | 35 (37%) | 24 (67%) |
| Moderate | 60 (40%) | 60 (63%) | 12 (33%) |
| High | 56 (37%) | 0 (0%) | 0 (0%) |
| **Percent vaccinated at sampling midpoint** |  |  |  |
| 0% | 129 (85%) | 75 (79%) | 26 (72%) |
| Above 0% up to 5% | 17 (11%) | 15 (16%) | 9 (25%) |
| Above 5% up to 10% | 3 (2.0%) | 3 (3.2%) | 1 (2.8%) |
| Above 10% | 2 (1.3%) | 2 (2.1%) | 0 (0%) |

<sup>1</sup>n (%)
<sup>2</sup>United Nations, Department of Economic and Social Affairs, Population Division (2018). World Urbanization Prospects: The 2018 Revision, Online Edition. File 13: Population of Capital Cities in 2018 (thousands) and File 16: Percentage of the Total Population Residing in Each Urban Agglomeration with 300,000 Inhabitants or More in 2018, by Country, 1950-2035.

Half of the studies used probability sampling (50%) and most sampled blood donors (45%, n=68) or households and communities (45%, n=68). The remaining studies sampled pregnant or parturient women (4%, n=6) and residual sera from outpatient clinics or primary health care facilities (6%, n=9). The most common study design was repeated cross-sectional (60%). Three studies in three countries (Burkina Faso, Mali, South Africa) used a longitudinal cohort design with probability sampling. Among the testing strategies used to measure seroprevalence, most studies used the Wantai SARS-CoV-2 Total Ab ELISA procured by WHO for standardisation (27%, n=41), other ELISA (33%, n=50), or CLIA (25%, n=37) assays. Sixteen studies used a lateral flow immunoassay (11%). Five studies (3.3%) had more than five percent of the national population vaccinated by the sampling midpoint, all in South Africa. The most frequent risk of bias rating was moderate (40%, n=60), followed by high (37%, n=56) and low risk of bias (23%, n=35). Risk of bias ratings for individual studies are reported in Supplement S4.3.

Ninety-five studies with low (37%, n=35) or moderate (63%, n=60) risk of bias (dataset 1) were included in the subsequent results (listed in Supplement S.4.2, Tables S5-S6).

### Seroprevalence and its geographic and temporal variation

Seroprevalence ranged from 0% to 87% in all studies. For countries in Eastern and Southern Africa, this range was 0% in a national blood donor study in Malawi conducted in January 2020,[29] to 73% in a local household sample in Addis Ababa, Ethiopia in April 2021.[30] For countries in Western and Middle Africa, this was 3% in a national household sample in Sierra Leone, March 2021,[31] to 87% in a national household sample in Ghana, December 2021.[32]

There was considerable variation in reported seroprevalence in the 17 countries in Dataset 1 (Figure S2). Countries with notable cohort or repeated cross-sectional studies (Malawi, Kenya, Ghana, and South Africa) are highlighted in Figure 2, indicating sharp increases in seroprevalence in the general population over time (Figure 2).

**Figure 1:**
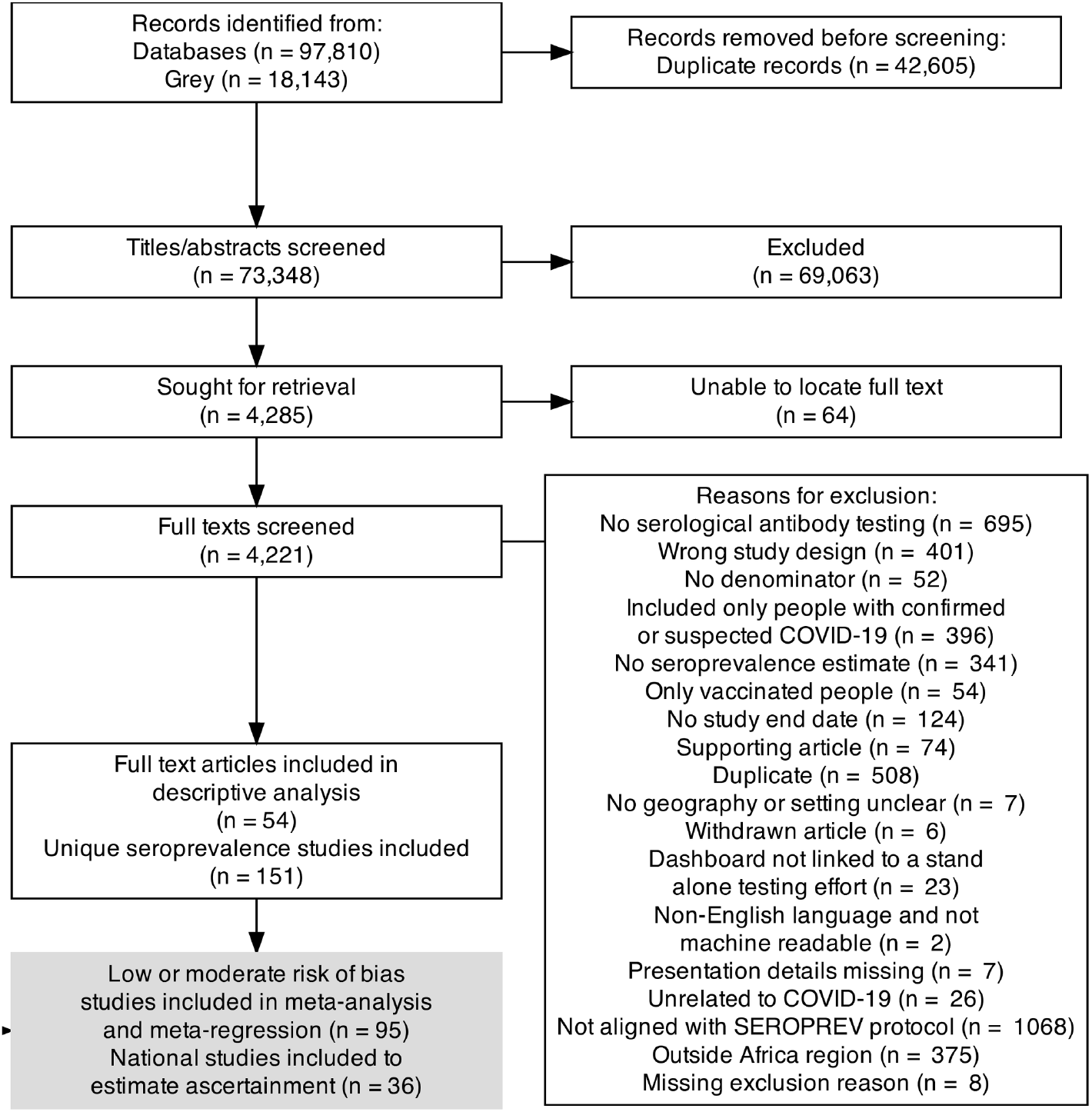
PRISMA Flow Diagram.

**Figure 2:**
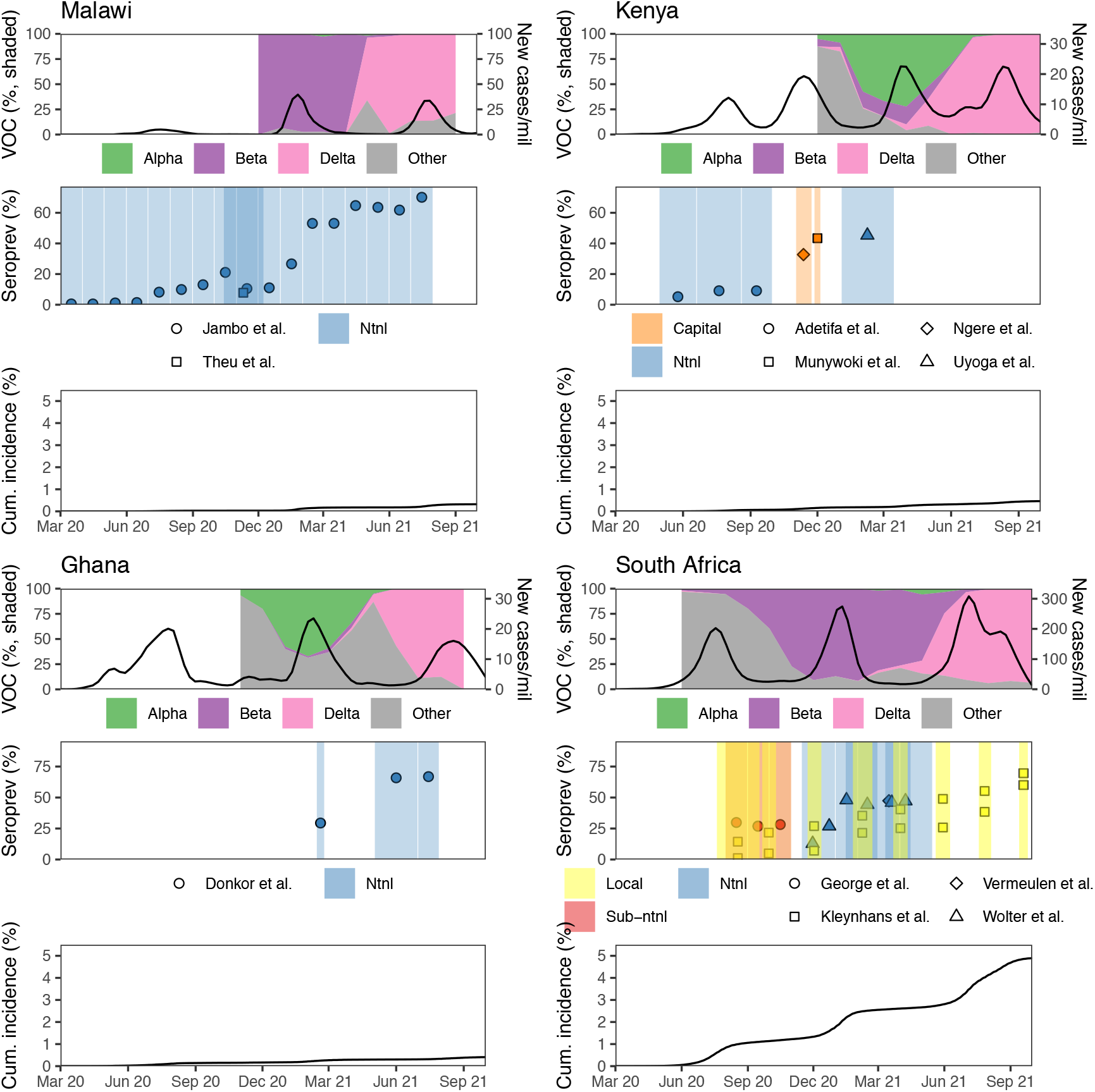
Reported seroprevalence, variants of concern, and cumulative incidence by selected countries over time, Mar 2020 - Sep 2021. • Plots for all countries are reported in Figure S2. • **Top panel, left axis:** Shaded areas represent the relative frequency of major variants of concern (VOC) circulating, based on weekly counts of hCoV-19 genomes submitted to GISAID we have aggregated by month. Weeks with fewer than 10 total submissions in a given country were excluded from the analysis. • **Top panel, right axis:** Daily confirmed cases reported to WHO on a national level per million people, smoothed using local regression (LOESS). • **Middle panel:** Each point is an individual seroprevalence study, and identical shapes represent studies originating from the same cohort or repeated cross-sectional data source. • **Bottom panel:** Cumulative incidence of confirmed cases per 100 people.

**Figure 3:**
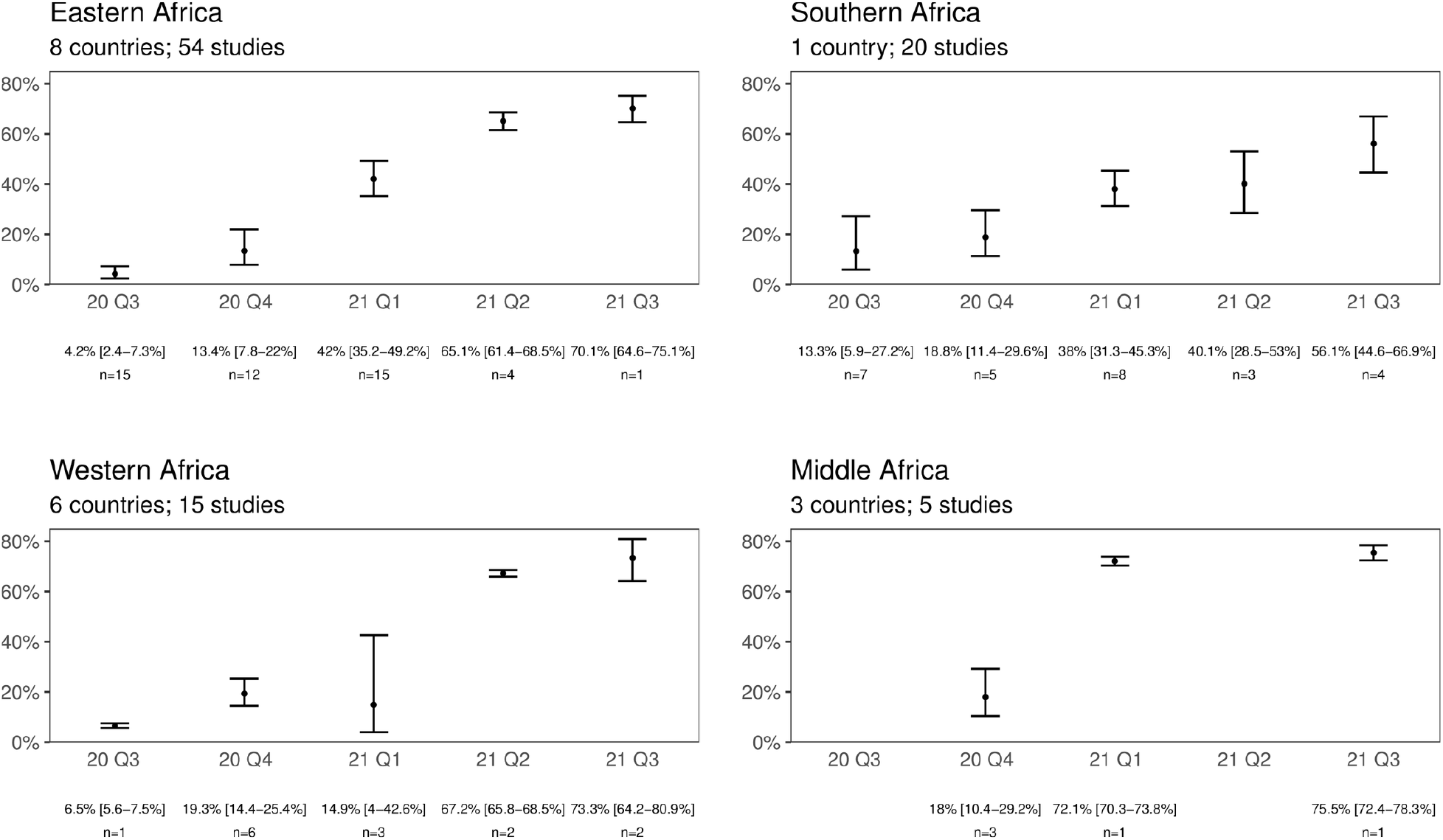
Pooled seroprevalence from infection or vaccination (random-effects model) by UN sub-region and quarter, Jul 2020 - Sep 2021. • Point estimates and 95% confidence intervals (error bars) are reported for each quarter, as well as the number of studies pooled (n).

### Pooled seroprevalence from infection or vaccination (random-effects model)

In the random-effects model, the pooled seroprevalence from infection or vaccination across Africa rose from 3.0% [95% CI: 1.0-9.2%] (number of studies=6, I2=96.0) as of Q2 2020 to 65.1%, [56.3-73.0%] (n=8, I2=98.6) as of Q3 2021.

Sub-regional meta-analyses were conducted for each quarter from 2020 Q2 to 2021 Q3, the period with available data (Q2 2020 results in Supplement Table S4). Pooled seroprevalence [95% CI] was 70.1% [64.6-75.1%] (n=1) in Eastern Africa, 56.1% [44.6-66.9%] (n=4) in Southern Africa, 73.3% [64.2-80.9%] (n=2) in Western Africa, and 75.5% [72.4-78.3%] (n=1) in Middle Africa as of Q3 2021.

### Seroprevalence to cumulative incidence ratios

There was considerable variation by country in the ratios of seroprevalence from infection (based on national studies) to cumulative incidence (Supplement Table S3). Ratios across all studies ranged from 10:1 to 958:1, with the highest under-ascertainment from 2020 Q4 to 2021 Q3 found in Nigeria (958:1, July 2021) and Malawi (696:1, October 2020), and the lowest in Sierra Leone (57:1, March 2021) and South Africa (10:1, November 2020). The ratios were moderately negatively correlated with health system access (Pearson’s r = -0.70), demand (−0.60), and resilience (−0.60). Health system quality was weakly negatively correlated with ascertainment (−0.26). Applying our overall estimated seroprevalence from infection to the African population suggests that true infections were 97 times larger than confirmed cases as of September 2021 (800 million infections compared to 8.2 million confirmed cases).

### Demographic, study, and country factors associated with seroprevalence

The percentage of seropositive individuals who did not report symptoms prior to sampling (asymptomatic seroprevalence) overall and by age and sex subgroups for fourteen studies reporting symptoms are shown in the supplement. Asymptomatic prevalence in Africa was 67.0% overall (IQR 47.8-80.9%, n=14) (Figure S3). Median asymptomatic prevalence was similar across age groups, ranging from 51.1% in ages 60+ to 71.0% in ages 20-29 (Kruskal-Wallis (KW) H-test p = 0.45). Median asymptomatic prevalence in males was 64.1%, compared to 70.1% in females (KW H-test p = 0.53).

Within studies, seroprevalence was lower for children 0-9 years compared to aged 20-29 years (prevalence ratio (PR) 0.72 [0.65-0.80]). There were no differences between other age groups and 20-29 years, nor between males and females (Figure 4). Within local studies, compared to urban geographical areas, seroprevalence was lower for rural geographical areas (PR 0.58 [0.45-0.74]).

**Figure 4:**
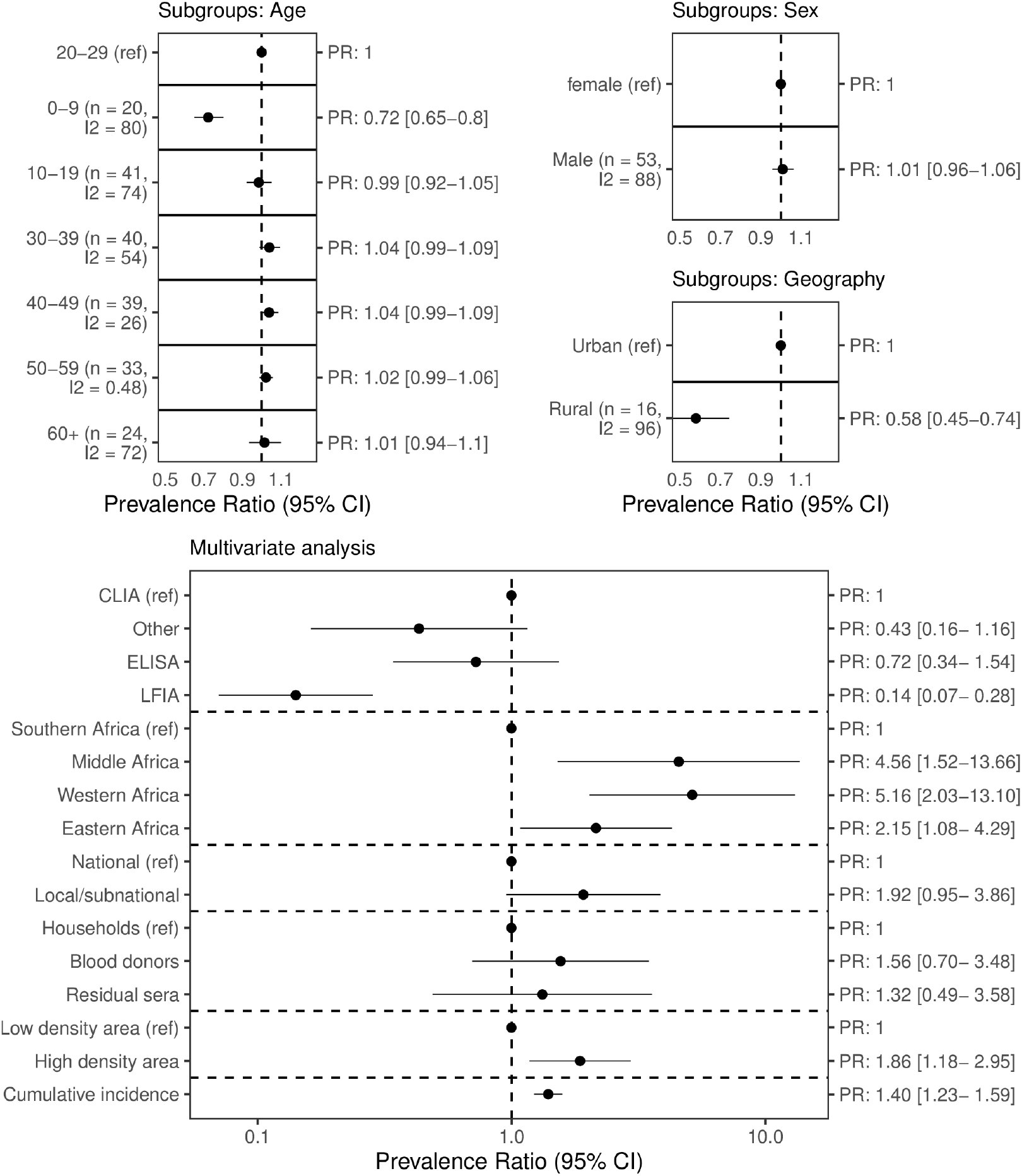
Factors associated with seroprevalence. • **Top panel: Meta-analysis results**. We calculated the ratio in prevalence between subgroups within each study then aggregated the ratios across studies using inverse variance-weighted random-effects meta-analysis. Heterogeneity was quantified using the I^2^ statistic. Each row represents a separate meta-analysis. • **Right panel:** Multivariate analysis using a Poisson generalised linear mixed-effects regression to model seroprevalence. • CLIA = Chemiluminescent immunoassay; ELISA = Enzyme-linked immunosorbent assay; LFIA = Lateral flow immunoassay. Cumulative incidence = Cumulative incidence of confirmed cases per 100 people.

In the meta-regression, high population density was associated with an increase in seroprevalence compared to studies in areas of low population density (PR 1.86 [118-2.95]). Compared to Southern Africa, Eastern Africa (PR 2.15 [1.08-4.29]), Western Africa (PR 5.16 [2.03-13.1]), and Middle Africa (PR 4.56 [1.52- 13.7]) were associated with higher seroprevalence. Higher cumulative incidence of confirmed cases was associated with an increase in seroprevalence (PR 1.40 [1.23-1.59]). Compared to studies that sampled households and communities, there were no differences between seroprevalence in studies that sampled blood donors nor residual sera. Finally, lateral flow assays were associated with lower seroprevalence compared to CLIA assays (PR 0.14 [0.07-0.28]), while there were no differences between ELISA nor other assays compared to CLIA assays, respectively.

### Sensitivity analysis

Sensitivity analyses accounting for serological test performance showed no qualitative differences from the primary results (Figures S4-S7).

## Discussion

We estimate that overall seroprevalence is now high in Africa (65.1% [56.3-73.0%] in Q3 2021) and has risen considerably over time, from 3.0% [1.0-9.2%] in Q2 2020, marked by sharp increases after the emergence of the Beta and Delta variants. Seroprevalence was highly heterogeneous both within countries - urban vs. rural (lower seroprevalence for rural geographic areas), children vs. adults (children aged 0-9 years had the lowest seroprevalence) - and between countries and African sub-regions (Eastern, Western, and Middle Africa regions associated with higher seroprevalence). Our results suggest that the ratio of seroprevalence to cumulative incidence of confirmed cases is large (97:1) in Africa overall, varying from 10:1 to 958:1 by country over the study period.

### Strengths and limitations

Previous assessments of COVID-19 seroprevalence studies highlight methodological heterogeneity as a key barrier to synthesising data.[33–35] The distinctive dataset described here is standardised, representative and granular, enabling unique insights into seroprevalence in Africa. Recently, in part via WHO’s UNITY Studies Initiative, more seroprevalence data has become available, disaggregated by demographic groups (age, sex), place (e.g. sub-region, country) and time (quarterly periods). In line with the equity principles of the UNITY Studies Initiative, our dataset 0 included a broad range of studies in low-income countries (62%, n=95), lower-middle-income countries (LMIC) (22%, n=33) and vulnerable HRP countries (41%, n=62). Around one quarter of studies were conducted at the national level, which is unique to this analysis. UNITY study collaborators shared timely evidence, facilitating geographic coverage and reducing publication bias. Additionally, standardised epidemiological and serological methods (including the supply of a well-performing assay to LMIC) enabled through the UNITY Studies Initiative means that the estimates included in our meta-analysis are robust and comparable. Finally, recognizing that assay performance is a key determinant of seroprevalence, we linked our data to independent test kit evaluations[27,28] of serological assay performance to correct seroprevalence estimates in a sensitivity analysis, helping ensure the robustness of our results to this source of bias.

This review should be considered in light of its limitations. Firstly, many local studies (29% in a city/town <300,000 people) were included in our estimates of seroprevalence over time (Dataset 1). These studies may not reflect the seroprevalence across an entire country: local studies are often conducted in large, dense, and interconnected urban centres to investigate suspected high transmission or for convenience. Indeed, our results show that local urban areas are associated with higher seroprevalence estimates. Secondly, our results are based only on studies aligned with the SEROPREV protocol and these may differ if using studies with other criteria. The SEROPREV protocol is broad, requiring a defined population, age standardisation, and a robust test, which tend to be indicators of higher quality seroprevalence studies. Based on studies identified through SeroTracker’s living systematic review, only 11 general population studies in Africa at low or moderate risk of bias were excluded from this study because they did not meet the eligibility criteria, and thus would likely have minimal impact on the main findings.[36] Thirdly, our meta-analysis estimates in certain quarters were driven by specific countries with a disproportionate number of studies, often in the same population over time, in Eastern Africa (Ethiopia: 17 studies, Malawi: 20 studies), Southern Africa (South Africa: 20 studies), and Western Africa (Ghana: 4 studies, Nigeria: 7 studies). Other sub-regions were underrepresented due to scarce data (e.g. no studies in North Africa, 5 studies in Middle Africa). Fourthly, we did not account for antibody waning, so the present study likely underestimates the true extent of past infection; similarly, case under-ascertainment is likely to be underestimated. Finally, due to delays in releasing seroprevalence study results, we could not produce estimates for Q4 2021, including since the emergence of the Omicron variant.

### Methodological quality of the included studies

We evaluated study risk of bias using a validated, seroprevalence-specific tool based on the Joanna Briggs Institute (JBI) Checklist for seroprevalence studies,[17] and found that 37% of studies were at high risk of bias. The most common reasons for this included an unrepresentative sample frame, non-probability sampling, and not adjusting estimates for population characteristics. We found a higher proportion of high-risk studies in our study compared to another SRMA in Africa (26%) by Chisale *et al*.[7] The interpretation of items in the JBI checklist is subjective and can vary considerably within and between study teams. Moreover, Chisale *et al*. evaluated overall risk of bias by summing each item’s score; the automated decision rule in our tool puts more weight on items more likely to bias results, such as whether a representative sample frame was chosen and a well-performing antibody test was used.[17]

To minimise the risk of bias in our summary estimates, we included only studies at low/moderate risk in our meta-analysis of seroprevalence in the general population over time, subgroup meta-analysis and meta-regression, and case ascertainment estimates. Despite this effort, there are still methodological differences between the meta-analysed studies that may reduce their comparability. For example, one fifth of studies in our analysis dataset (19%, n=18/95) were convenience samples, which are less representative than population-based probability samples. To limit this bias, we required UNITY-Aligned convenience samples to have a clearly defined sample frame (i.e., sampling of volunteers excluded).

### Context

The pooled seroprevalence in Africa estimated in this study (65.1% in Q3 2021) is among the highest in the world (comparable to the South-East Asia region).[9] With vaccination coverage in Africa being low during the study period (4.8% in September 2021),[6] this is mostly driven by infections. Our time- and region-specific estimates shed light on the trajectory of the pandemic. Of interest, we observe sharp increases in seroprevalence in 2021 in certain countries and regions, signifying the considerable number of infections caused by more transmissible variants [21]: for example, an increase of 11% (Dec 2020) to 65% (Apr 2021) seroprevalence in Malawi following the emergence of the Beta variant; and 26% (May 2021) to 60% (Sept 2021) in rural South Africa following Delta variant emergence. While case counts also increased during this time, this systematic comparison of seroprevalence data from different countries demonstrates the inexorable spread of infection in regions with limited or variable PHSM [37] and vaccination roll-out, particularly where highly transmissible variants are circulating.

One possible explanation for high seroprevalence is potential cross-reactivity of antibodies against P. falciparum or common cold coronaviruses (CCC) In areas of Africa with a high incidence of malaria,[38,39] malaria cross-reactivity could follow two potential mechanisms: a) SARS-CoV-2 antibody tests falsely reacting in malaria hyperendemic areas, [40] or b) cross-protection through CD8+ t-cell activation from *P. falciparum* antigens [41] Broad anti-CCC antibodies have been identified following SARS-CoV-2 infection and vaccination.[42,43] The exact role of cross-reactivity on seroprevalence estimates warrant further investigation.

In multivariate analysis, seroprevalence was heterogeneous between sub-regions: higher in Eastern, Western, and Middle Africa compared to Southern Africa. The exact reasons for this heterogeneity remain unknown but could be related to mitigation strategy, health infrastructure, and the effectiveness of PHSM implementation. The capacity to isolate has been shown to vary greatly in Africa, [44] and challenges have been reported with social distancing in the Western and Middle sub-regions, especially in high density areas.[45,46] This is also consistent with findings by Chisale *et al*.[7], who observed higher seroprevalence in studies conducted in Central Africa compared to other regions.

We observed heterogeneity within countries by age group and urban/rural geography. We observed lower seroprevalence in children 0-9, perhaps attributable to milder infections in this group which are associated with lower antibody titres [47] and school closures which have been common and lengthy in some African countries.[48] In contrast to our global analysis,[9] we did not find lower seroprevalence in adults 60+ years compared to adults 20-29 years. These results may be due to increased intergenerational mixing at the household level, as prior research in Africa has shown that households sharing space with persons ages 60+ may have increased transmission risk [44]. We also found lower seroprevalence was associated with rural geographical areas compared to urban areas, in line with other hypotheses and modelling associating rural areas with a potentially lower spread of infection due to decreased population density.[49]

The use of lateral flow immunoassays (LFIA) was associated with lower seroprevalence estimates compared to CLIA assays in the multivariate meta-regression. This may be explained by the lower sensitivity of LFIA, which can lead to more false negatives and underestimated seroprevalence.[50]

We observed greatly varying ratios between seroprevalence to cumulative confirmed case incidence by country, ranging from 10:1 in South Africa to 958:1 in Nigeria. The large fraction of asymptomatic cases in Africa is one possible reason for this — our estimates suggest that 67.0% (IQR 47.8-80.9%) of cases have no symptoms, which accords with previous work documenting higher rates of asymptomatic infections in Africa compared to other regions.[35,51] Under-ascertainment is well documented in African countries with low capacity for surveillance and laboratory testing,[4] and indeed, lower levels of under-ascertainment were observed in countries with higher health system functionality indices. We estimate larger ratios compared to other parts of the world during the same period.[9]

### Implications for practice, policy and future research

In contrast to routine surveillance data that rely on case reports, recently available seroprevalence studies have provided a more accurate understanding of the true extent of SARS-CoV-2 infection across Africa, amid low vaccine coverage. There is a need to strengthen surveillance infrastructure for priority diseases, including collaborative government-researcher efforts and timely reporting mechanisms. Seroprevalence data needs to be used alongside other sources of epidemiological data for policy-decision making. This will collectively inform effective, and tailored disease prevention control programmes, which must be deployed alongside other investments in public health and health system strengthening (e.g. trained and motivated health workers, a well-maintained infrastructure, and a reliable supply of medicines and technologies) to support their implementation and uptake in Africa for COVID-19 and other future or existing infectious disease threats.

The geographic distribution of early unpublished results shared with WHO for inclusion in this systematic review and meta-analysis demonstrates the need for standardised initiatives to help build enhanced surveillance and research capacity in LMIC. In this study, there were only two Unity-Aligned studies in Northern Africa (both in Egypt) identified here. Several populous and/or HRP countries lack study results to determine seroprevalence in the general population (e.g., Ethiopia: no nationwide study results available since July 2020; Angola and DRC: no nationwide study results; Algeria, United Republic of Tanzania, Morocco, Somalia, and Tunisia: no nationwide or local study results). Furthermore, there were no results available from island nations. We are aware of studies ongoing in some of these countries, emphasising the importance of open data practises, sharing early results, and collation and analysis of timely data. Improved quality and transparency of reporting studies would also help address this [52]: several studies were excluded from this SRMA due to insufficient information (e.g. no denominator stated, no end date stated, setting unclear).

Given the burden of SARS-CoV-2 varies across regions in Africa, there is also a need for studies representing different contexts (e.g. heterogeneous access to health services, fragile environments, HRP countries) and vulnerable populations (e.g. those with endemic infections like HIV and comorbidities, those living in high density urban settlements, refugee populations). Furthermore, there is a need to continue studying population differences in SARS-CoV-2 infection (e.g. age, sex, geography, race etc.) to identify susceptible sub-populations to inform priorities for vaccination coverage and prevention and control measures. There is sparse data over time in many countries, which indicates a need to conduct more spatiotemporal studies (e.g. longitudinal cohort, repeated cross-sectional). The use of well-designed convenience samples can help achieve this — for example, studies in blood donors represent over half of identified studies that used a cohort or repeated cross-sectional design (57%, n=64/112). Our results suggest that blood donors are a good proxy for the general population, as we found no statistical difference between seroprevalence estimates in blood donors and those in households and communities.

Population-based seroprevalence studies primarily give a reliable estimate of the exposure to SARS-CoV-2 infection in the general population. Where studies measure antibodies quantitatively, they can also estimate correlates for protection against infection.[53] The risk of reinfection with the Omicron variant is reported to be much higher than for previous variants in both SARS-CoV-2 infected and vaccinated individuals.[54] This implies that the presence of SARS-CoV-2 antibodies may no longer be a correlate of protection against infection for Omicron. However, seroprevalence estimates remain indicative of protection against severe disease, as cellular immunity is unlikely to be affected even in the case of highly mutated variants such as Omicron.

## Conclusion

This updated meta-analysis in Africa provides robust and representative seroprevalence results from over 40% of the continent’s Member States, enabled through the standardisation and adaptability of the WHO Unity Studies. The substantial under-ascertainment of SARS-CoV-2 infection indicates that the majority of cases of SARS-CoV-2 in Africa are not captured by national surveillance systems, emphasising the continued need for comparable, aggregated, and timely seroprevalence data that accounts for changing vaccination coverage. Our work provides a platform to develop Africa’s surveillance portfolio, building on existing local capacity to enable targeted and regular seroprevalence studies in sentinel countries. SARS-CoV-2 seroprevalence is high and heterogenous in Africa suggesting greater population exposure to SARS-CoV-2 and protection against COVID-19 disease than has been previously indicated by confirmed case data and vaccine coverage. As such, PHSM and vaccination strategies tailored to local settings and specific populations are warranted.

## Data Availability

Detailed information on each study is available in Supplement S4.2 and references are cited in the Supplementary File (S4.4). Primary seroprevalence estimates and statistical code relevant to the study are available in a public, open access repository:
https://github.com/serotracker/Africa-SRMA-seroprevalence/tree/1a7e88ed2679d4027092b2898cf7048426e8b1b6
Seroprevalence estimates by age and sex subgroups relevant to the study are uploaded to Zenodo by UNITY study collaborators and DOIs for each included Zenodo study are cited in the Supplementary File (S4.4). Data are available to download upon reasonable request. https://zenodo.org/communities/unity-sero-2021?page=1&size=20.

https://zenodo.org/communities/unity-sero-2021/?page=1&size=20

## Unity Studies Collaborator Group

<u>**UNITY Study Collaborator Group</u>: Rosemary A Audu^1^, Jacob S Barnor^2^, Enyew Birru^3^, Henry K Bosa^4,5^, Emily L Boucher^6,7^, Annie Chauma-Mwale^8^, Matthew P Cheng^9^, Judy Chen^10^, Cheryl Cohen^11,12^, Tienhan S Dabakuyo-Yonli^13^, Gabriel Deveaux^6^, Boly Diop^14^, Titus H Divala^15^, Emily K Dokubo^16^, Irene O Donkor^17^, Claire Donnici^6^, Nathan Duarte^10^, Natalie A Duarte^18^, Timothy G Evans^19^, Lee Fairlie^20^, Ousmane Faye^21^, Anne von Gottberg^11,22^, Tiffany G Harris^23,24^, Natasha Ilincic^25^, Elsie A Ilori^26^, Jackie Kleynhans^11,12^, Dayoung Kim^27^, Olatunji M Kolawole^28,29^, Jambo C Kondwani^30,31^ Michael Liu^32^, Emma Loeschnik^33^, Sheila Makiala-Mandanda^34,35^, Alexandre Manirakiza^36^, Pinyi N Mawien^37^, Portia C Mutevedzi^38,39^, Jason M Mwenda^1^, Eric M Osoro^40,41^, Samiratou Ouedraogo^10,42^, Sara Perlman-Arrow^7^, Jesse Papenburg^43,44^, Hude Quan^45^, Hannah P Rahim^46^, Karampreet Sachathep^47,48^, Shobna Sawry^49^, Mitchell Segal^25^, Anabel Selemon^6^, Judith Shang^50^, Kristen A Stafford^51,52^, Laura Steinhardt^53^, Cheikh Talla^54^, Halidou Tinto^55^, Isidore T Traore^56,57^, Joe A Theu^58^, Ines Vigan-Womas^59^, Tyler Williamson^60^, Tingting Yan^25^, Cedric P. Yansouni^61,62^, Caseng Zhang^63^

^1^Nigerian Institute of Medical Research, Nigeria

^2^World Health Organization, Regional Office for Africa, Brazzaville, Congo

^3^Ethiopian Public Health Institute, Addis Ababa, Ethiopia

^4^Ministry of Health, Uganda

^5^Kellog College, University of Oxford, Oxford, United Kingdom

^6^Cumming School of Medicine, University of Calgary, Calgary, Canada

^7^Nuffield Department of Clinical Neurosciences, University of Oxford, Oxford, UK

^8^Public Health Institute of Malawi, Ministry of Health, Malawi

^9^Divisions of Infectious Diseases and Medical Microbiology, McGill University Health Centre, Montreal, Canada

^10^McGill University, Montreal, Canada

^11^Center for Respiratory Disease and Meningitis, National Institute for Communicable Diseases, Johanessburg, South Africa

^12^School of Public Health, Faculty of Health Sciences, University of the Witwatersrand, Johannesburg, South Africa

^13^Epidemiology and Quality of Life Research Unit, INSERM U1231, Georges François Leclerc Centre – UNICANCER, Dijon, France

^14^Surveillance Division, Prevention Directorate, Ministry of Health and Social Action, Dakar, Senegal

^15^Kamuzu University of Health Sciences, University in Blantyre, Malawi

^16^United States Centers for Disease Control and Prevention, Atlanta, United States of America

^17^Noguchi Memorial Institute for Medical Research, Accra, Ghana

^18^Faculty of Arts and Science, University of Toronto, Toronto, Canada ^19^COVID-19 Immunity Task Force, McGill University, Montreal, Canada

^20^WITS Reproductive Health and HIV Institute, Faculty of Health Sciences, University of the Witwatersrand, South Africa

^21^Virology Department, Institut Pasteur de Dakar, Dakar, Senegal

^22^School of Pathology, Faculty of Health Sciences, University of the Witwatersrand, Johannesburg, South Africa

^23^The International Center for AIDS Care and Treatment Programs (ICAP), Columbia University, New York, United States

^24^Department of Epidemiology, Mailman School of Public Health, Columbia University, United States of America

^25^University of Toronto, Toronto, Canada

^26^Nigeria Centre for Disease Control, Nigeria

^27^Faculty of Science, University of Calgary, Calgary, Canada

^28^Department of Microbiology, Faculty of Life Sciences, University of Ilorin, Ilorin, Nigeria

^29^Ministerial Expert Advisory Committee on COVID-19, Health Sector Response, Federal Ministry of Health, Abuja

^30^Malawi-Liverpool-Wellcome Clinical Research Programme, Malawi

^31^Liverpool School of Tropical Medicine, United Kingdom

^32^Harvard Medical School, Harvard University, Cambridge, USA

^33^Department of Epidemiology and Biostatistics, Schulich School of Medicine and Dentistry, Western University, Ontario, Canada

^34^Institut National de Recherche Biomédicale, Kinshasa, Democratic Republic of the Congo

^35^Université de Kinshasa, Democratic Republic of the Congo

^36^Institut Pasteur of Bangui, Central African Republic

^37^Preventive Health Services, Ministry of Health, Juba, South Sudan

^38^South African Medical Research Council Vaccines and Infectious Diseases Analytics Research Unit, Faculty of Health Sciences, University of the Witwatersrand, Johannesburg, South Africa

^39^School of Pathology, Faculty of Health Sciences, University of Witwatersrand, South Africa

^40^Washington State University, Global Health Kenya, Nairobi, Kenya

^41^Paul G. Allen School of Global Health, Washington State University, United States of America

^42^National Public Health Institute, Burkina Faso

^43^Montreal Children’s Hospital, McGill University Health Centre, Montreal, Canada

^44^Department of Epidemiology, Biostatistics and Occupational Health, McGill University, Montreal, Canada

^45^Department of Community Health Sciences, University of Calgary, Calgary, Canada

^46^Boston Consulting Group, Boston, USA

^47^The International Center for AIDS Care and Treatment Programs (ICAP), Columbia University, New York, United States of America

^48^Department of Population and Family Health, Mailman School of Public Health, New York, United States of America

^49^WITS Reproductive Health and HIV Institute, Faculty of Health Sciences, University of the Witwatersrand, South Africa

^50^United States Centers for Disease Control and Prevention (US-CDC), Cameroon

^51^Center for International Health, Education, and Biosecurity, University of Maryland School of Medicine, United States of America

^52^Division of Epidemiology and Prevention, Institute of Human Virology, University of Maryland School of Medicine, United States of America

^53^United States Centers for Disease Control and Prevention, Atlanta, United States of America

^54^Epidemiology, Clinical Research and Data Science, Institut Pasteur de Dakar, Senegal ^55^Institut de Recherche en Science de la Santé - Clinical Research Unit of Nanoro, Burkina Faso

^56^Programme de Recherche sur les maladies infectieuses, Centre MURAZ, Bobo-Dioulasso, Burkina Faso

^57^Institut Supérieur des Sciences de la Santé, Université Nazi Boni, Bobo-Dioulasso, Burkina Faso

^58^International Training and Education Center for Health (I-TECH), Malawi

^59^Immunophysiopathology and Infectious Diseases Department, Institut Pasteur de Dakar, Senegal

^60^Department of Community Health Sciences, University of Calgary, Calgary, Canada

^61^Divisions of Infectious Diseases and Medical Microbiology, McGill University Health Centre, Montreal, Canada

^62^J.D. MacLean Centre for Tropical Diseases, McGill University Health Centre

^63^McMaster University, Hamilton, Canada

## Acknowledgements

We thank colleagues at WHO and partner organisations including Kathleen Gallagher, Amen Ben Hamida, Christopher Murrill, Toni Whistler, Venkatachalam Udhayakumar (US Centers for Disease Control and Prevention); Vincent Richard (Institut Pasteur and the Institut Pasteur International Network); individuals to be confirmed (Africa CDC); Robert Blanchard and the WHO/Dubai Logistics Team; Nollascus Ganda, Solome Okware, Joseph Wamala (WHO Country Offices in the Africa Region) and Michael Ryan (WHO HQ)

SeroTracker (led by RKA, including MW, HW, ZL, XM, TY, CC, MYL, JP, MPC, DB, ML, MS, GRD, NI, CZ, SP, HPR, TY, KCN, DK, SAA, ND, CD, NAD, EL, RKI, ASB, ELB, AS, JC). is grateful for support from WHO, Canada’s COVID-19 Immunity Task Force through the Public Health Agency of Canada, the Robert Koch Institute, and the Canadian Medical Association Joule Innovation Fund. RKA additionally thanks the Rhodes Trust for its support.

## Contributors

Conceptualization: HCL, IB, LS, TN, MVK, RKA, NB.

Data curation: HCL, AnV, LA, JCO, TA, LL, AiV, HW, MW, XM, ZL, NB, CC, MYL, ELB, JC, GRD, CD, ND, NAD, NI, DK, ML, EL, KCN, SP, HPR, MJS, AS, TY, CZ.

Formal analysis: HW, RKA, DB, JP, MPC.

Funding acquisition: IB, JCO, MVK, RKA, NB, TY.

Investigation: HCL, IB, LS, AN, MVK, BC, AnV, LA, AR, JCO, TA, PW, LL, AiV, RAA, EB, AC, CCo, TSD, BD, THD, EKD, LF, TGH, EAI, KCJ, JK, OMK, HKB, SM, AM, PCM, EMO, SO, IOD, KKS, SS, KAS, LS, AOT, CT, JAT, HT, ITT, IV, AvG, CPY.

Methodology: HW, RKA, DB, NB, IB, HCL, AN, MV, MVK, CC, MPC, DC, CD, ND, ML, JP, MYL, TW, TE, HQ.

Project administration: HCL, IB, JCO, MVK, MW, NB, RKA, TY, RAA, PNA, TB, JSB, BLH, OMK, PCM, JMM, AN, NN, JDS, LS, AOT, HT, MV, AvG, CPY.

Resources: HCL, LS, BC, AnV, LA, AR, JCO, TA, LL, AiV.

Supervision: IB, AnV, LA, AR, JCO, TA, PRW, LL, RP, MVK, DB, MPC, JP, RKA, NB, TY.

Writing – original draft: HCL, IB, LS, AN, HW, MW, RKA.

Writing – review & editing: all authors.

All authors debated, discussed, edited, and approved the final manuscript. All authors had full access to the full data in the study and accepted responsibility to submit for publication

## Data availability statement

Detailed information on each study is available in Supplement S4.2 and references are cited in the Supplementary File (S4.4). Primary seroprevalence estimates and statistical code relevant to the study are available in a public, open access repository: https://github.com/serotracker/Africa-SRMA-seroprevalence/tree/1a7e88ed2679d4027092b2898cf7048426e8b1b6

Seroprevalence estimates by age and sex subgroups relevant to the study are uploaded to Zenodo by UNITY study collaborators and DOIs for each included Zenodo study are cited in the Supplementary File (S4.4). Data are available to download upon reasonable request. https://zenodo.org/communities/unity-sero-2021?page=1&size=20.[11]

## Supplementary Material

Supplementary material file is attached separately.

