## Supplementary Material for "SARS-CoV-2 infection in Africa: A systematic review and meta-analysis of standardised seroprevalence studies, from January 2020 to December 2021"

**S1: PRISMA Checklist**	**2**

**S2: Supplementary Methods**	**5**

Search strategy, screening, extraction	5

Table S1: Study- and Estimate-level Classification Decisions	6

**S3: Supplementary Analysis**	**8**

S3.1 Estimate prioritization	8

S3.2 Additional analysis description	9

Table S2: Population density classification decisions	9

**S4: Supplementary Results**	**12**

S4.1 Supplemental tables and figures	12

Table S3: Ratios of seroprevalence to cumulative incidence of confirmed cases by subregion and country, 2020 Q4-2021 Q3	12

Table S4: Pooled seroprevalence (random-effects model) by UN sub-region, Mar 2020 - Sep 2021	12

Figure S1: African Member States with seroprevalence data identified, 2020-2021 (dataset 0: all studies)	14

Figure S2: Reported seroprevalence, variants of concern, cumulative incidence, and vaccination by countries over time, Mar 2020 - Sep 2021	15

Figure S3: Asymptomatic seroprevalence by age and sex	25

Figure S4: Adjusted vs. unadjusted seroprevalence data.	25

Figure S5: Pooled seroprevalence from infection or vaccination (random-effects model) by UN sub-region using adjusted and unadjusted seroprevalence data.	26

Figure S6: Seroprevalence to confirmed case ratios using adjusted and unadjusted seroprevalence data.	26

Figure S7: Meta-regression using adjusted seroprevalence data.	27

Figure S8: Risk of bias summary of studies included in descriptive analysis (dataset 0)	28

S4.2 Studies included in ascertainment analysis, meta-analysis, and descriptive analysis	29

Table S5. Studies used in ascertainment analysis, meta-analysis, and descriptive analysis (dataset 2: national studies rated low and moderate risk of bias).	29

Table S6. Studies used in meta-analysis and descriptive analysis (dataset 1: local and sub-national studies rated low and moderate risk of bias. For national studies in dataset 1, see Table S5).	41

Table S7. Studies used in descriptive analysis (dataset 0: studies rated high risk of bias. For studies rated low and moderate risk of bias, see Tables S5 and S6).	56

S4.3 Risk of bias ratings for all studies	67

S4.4 References for all studies	95

**S5: References Cited in Supplementary File**	**99**

### S1: PRISMA Checklist

| **Section and Topic** | **Item #** | **Checklist item** | **Location where item is reported** |
| --- | --- | --- | --- |
| **TITLE** | | |  |
| Title | 1 | Identify the report as a systematic review. | Title page |
| **ABSTRACT** | | |  |
| Abstract | 2 | See the PRISMA 2020 for Abstracts checklist. | P. 2 |
| **INTRODUCTION** | | |  |
| Rationale | 3 | Describe the rationale for the review in the context of existing knowledge. | P. 4 |
| Objectives | 4 | Provide an explicit statement of the objective(s) or question(s) the review addresses. | P. 4 |
| **METHODS** | | |  |
| Eligibility criteria | 5 | Specify the inclusion and exclusion criteria for the review and how studies were grouped for the syntheses. | P. 5 |
| Information sources | 6 | Specify all databases, registers, websites, organisations, reference lists and other sources searched or consulted to identify studies. Specify the date when each source was last searched or consulted. | P. 5 |
| Search strategy | 7 | Present the full search strategies for all databases, registers and websites, including any filters and limits used. | S2 |
| Selection process | 8 | Specify the methods used to decide whether a study met the inclusion criteria of the review, including how many reviewers screened each record and each report retrieved, whether they worked independently, and if applicable, details of automation tools used in the process. | P. 5, S2 |
| Data collection process | 9 | Specify the methods used to collect data from reports, including how many reviewers collected data from each report, whether they worked independently, any processes for obtaining or confirming data from study investigators, and if applicable, details of automation tools used in the process. | P. 5, S2 |
| Data items | 10a | List and define all outcomes for which data were sought. Specify whether all results that were compatible with each outcome domain in each study were sought (e.g. for all measures, time points, analyses), and if not, the methods used to decide which results to collect. | P. 5, S3.1 |
|  | 10b | List and define all other variables for which data were sought (e.g. participant and intervention characteristics, funding sources). Describe any assumptions made about any missing or unclear information. | P. 5-6, Table S1 |
| Study risk of bias assessment | 11 | Specify the methods used to assess risk of bias in the included studies, including details of the tool(s) used, how many reviewers assessed each study and whether they worked independently, and if applicable, details of automation tools used in the process. | P. 5-6 |
| Effect measures | 12 | Specify for each outcome the effect measure(s) (e.g. risk ratio, mean difference) used in the synthesis or presentation of results. | P. 6-7 |
| Synthesis methods | 13a | Describe the processes used to decide which studies were eligible for each synthesis (e.g. tabulating the study intervention characteristics and comparing against the planned groups for each synthesis (item #5)). | P. 6-7 |
|  | 13b | Describe any methods required to prepare the data for presentation or synthesis, such as handling of missing summary statistics, or data conversions. | P. 6-7, S3.1 |
|  | 13c | Describe any methods used to tabulate or visually display results of individual studies and syntheses. | P. 6-7 |
|  | 13d | Describe any methods used to synthesize results and provide a rationale for the choice(s). If meta-analysis was performed, describe the model(s), method(s) to identify the presence and extent of statistical heterogeneity, and software package(s) used. | P. 6-7 |
|  | 13e | Describe any methods used to explore possible causes of heterogeneity among study results (e.g. subgroup analysis, meta-regression). | P. 7 |
|  | 13f | Describe any sensitivity analyses conducted to assess robustness of the synthesized results. | P. 7 |
| Reporting bias assessment | 14 | Describe any methods used to assess risk of bias due to missing results in a synthesis (arising from reporting biases). | N/A |
| Certainty assessment | 15 | Describe any methods used to assess certainty (or confidence) in the body of evidence for an outcome. | P. 6-7 |
| **RESULTS** | | |  |
| Study selection | 16a | Describe the results of the search and selection process, from the number of records identified in the search to the number of studies included in the review, ideally using a flow diagram. | Figure 1 |
|  | 16b | Cite studies that might appear to meet the inclusion criteria, but which were excluded, and explain why they were excluded. | N/A |
| Study characteristics | 17 | Cite each included study and present its characteristics. | Table S5-S7, S4.4 |
| Risk of bias in studies | 18 | Present assessments of risk of bias for each included study. | Table S8 |
| Results of individual studies | 19 | For all outcomes, present, for each study: (a) summary statistics for each group (where appropriate) and (b) an effect estimate and its precision (e.g. confidence/credible interval), ideally using structured tables or plots. | Figure 2, Figure S2, Table S5-S7 |
| Results of syntheses | 20a | For each synthesis, briefly summarise the characteristics and risk of bias among contributing studies. | Table 1 |
|  | 20b | Present results of all statistical syntheses conducted. If meta-analysis was done, present for each the summary estimate and its precision (e.g. confidence/credible interval) and measures of statistical heterogeneity. If comparing groups, describe the direction of the effect. | P. 10-11, Figure 3, Table S4 |
|  | 20c | Present results of all investigations of possible causes of heterogeneity among study results. | P. 11-12, Figure 4 |
|  | 20d | Present results of all sensitivity analyses conducted to assess the robustness of the synthesized results. | P. 12, Figure S4-S7 |
| Reporting biases | 21 | Present assessments of risk of bias due to missing results (arising from reporting biases) for each synthesis assessed. | N/A |
| Certainty of evidence | 22 | Present assessments of certainty (or confidence) in the body of evidence for each outcome assessed. | Figure 3, Figure 4, Table S4, Figure S4-S7 |
| **DISCUSSION** | | |  |
| Discussion | 23a | Provide a general interpretation of the results in the context of other evidence. | P. 14-15 |
|  | 23b | Discuss any limitations of the evidence included in the review. | P. 13 |
|  | 23c | Discuss any limitations of the review processes used. | P. 13 |
|  | 23d | Discuss implications of the results for practice, policy, and future research. | P. 15-16 |
| **OTHER INFORMATION** | | |  |
| Registration and protocol | 24a | Provide registration information for the review, including register name and registration number, or state that the review was not registered. | P. 5 |
|  | 24b | Indicate where the review protocol can be accessed, or state that a protocol was not prepared. | P. 5 |
|  | 24c | Describe and explain any amendments to information provided at registration or in the protocol. | P. 5 |
| Support | 25 | Describe sources of financial or non-financial support for the review, and the role of the funders or sponsors in the review. | P. 18 |
| Competing interests | 26 | Declare any competing interests of review authors. | P. 18-19 |
| Availability of data, code and other materials | 27 | Report which of the following are publicly available and where they can be found: template data collection forms; data extracted from included studies; data used for all analyses; analytic code; any other materials used in the review. | P. 5, P. 19 |

### S2: Supplementary Methods

#### Search strategy, screening, extraction

Full detail on our inclusion and exclusion criteria, screening, and extraction processes are described elsewhere.[[1]](https://www.zotero.org/google-docs/?hqoiMN) Preliminary screening was conducted using Covidence software. Studies were then uploaded to AirTable for extraction of summary-level estimates and study factors.

Database: Ovid MEDLINE(R) and Epub Ahead of Print, In-Process & Other Non-Indexed Citations and Daily

Dates: January 1, 2020 to December 30, 2021

Notes: Covid-19 search terms were adapted from Ovid Expert Searches

| **#** | **Search terms** |
| --- | --- |
| 1 | exp Coronavirus/ |
| 2 | exp Coronavirus Infections/ |
| 3 | (coronavirus* or corona virus* or OC43 or NL63 or 229E or HKU1 or HCoV* or ncov* or covid* or sars-cov* or sarscov* or Sars-coronavirus* or Severe Acute Respiratory Syndrome Coronavirus*).tw,kf.[EB2] |
| 4 | or/1-3 |
| 5 | 4 not ((MERS or MERS-CoV or Middle East respiratory syndrome or camel* or dromedar* or equine or coronary or coronal or covidence* or covidien or influenza virus or HIV or bovine or calves or TGEV or feline or porcine or BCoV or PED or PEDV or PDCoV or FIPV or FCoV or SADS-CoV or canine or CCov or zoonotic or avian influenza or H1N1 or H5N1 or H5N6 or IBV).mp. or (animals/ not humans/)) |
| 6 | ((pneumonia or covid* or coronavirus* or corona virus* or ncov* or 2019-ncov or sars* or virus).tw,kf. or exp pneumonia/) and Wuhan.tw,kf. |
| 7 | (2019-ncov* or 2019nCov* or ncov19 or ncov-19 or 2019-novel CoV or sars-cov2* or sars-cov-2* or sarscov2* or sarscov-2* or Sars-coronavirus2 or Sars-coronavirus-2 or SARS-like coronavirus* or coronavirus 2 or coronavirus2* or corona or coronavirus-19 or covid19 or covid-19 or covid 2019 or ((novel or new or nouveau) adj2 (CoV or nCoV or covid or coronavirus* or corona virus or Pandemi*2)) or ((covid or covid19* or covid-19) and pandemic*2) or (coronavirus* and pneumonia)).tw,kf. |
| 8 | COVID-19.rx,px,ox. or severe acute respiratory syndrome coronavirus 2.os. |
| 9 | or/6-8 |
| 10 | 5 or 9 |
| 11 | immunoglobulins/ or antibodies/ or antibodies, blocking/ or exp antibodies, neutralizing/ or antibodies, viral/ or antigen-antibody complex/ or immune sera/ or exp immunoglobulin isotypes/ or immunoglobulin a/ or immunoglobulin d/ or immunoglobulin e/ or immunoglobulin g/ or immunoglobulin m/ |
| 12 | serologic tests/ or complement fixation tests/ or hemagglutination inhibition tests/ or neutralization tests/ |
| 13 | immunoassay/ or fluoroimmunoassay/ or exp immunoblotting/ or immunoenzyme techniques/ or exp enzyme-linked immunosorbent assay/ or exp enzyme-linked immunospot assay/ or immunosorbent techniques/ or serologic tests/ or complement fixation tests/ or hemagglutination inhibition tests/ or neutralization tests/ or Serology/di |
| 14 | (enzyme linked immunosorbent or enzyme-linked immunosorbent or ELISA or immunofluorescence or complement fixation or hemagglutination inhibition or immunoblot or western blot or neutrali*).tw,kf. |
| 15 | (antibod* or immunoglobulin* or immune globulin* or titer* or isotype* or IgG or IgM or IgA or neutrali* or sera or serum or serolog* or saliva).tw,kf. |
| 16 | or/11-14 |
| 17 | seroepidemiologic studies/ |
| 18 | incidence/ or prevalence/ |
| 19 | (seroconver* or seroprevalence or sero-prevalence or seroincidence or sero-incidence or seroepidemiolog* or sero-epidemiolog*).mp. |
| 20 | (inciden* or prevalen* or count* or rate*).mp. |
| 21 | (serosurvey or sero-survey or screen* or diagnostic).mp. |
| 22 | (seroconver* or seroprevalence or sero-prevalence or seroincidence or sero-incidence or seroepidemiolog* or sero-epidemiolog* or inciden* or prevalen* or silent or asymptomatic or serosurvey or sero-survey).tw,kf. |
| 23 | or/17-21 |
| 24 | 10 and (16 and 23) |
| 25 | 10 and 15 |
| 26 | 10 and 22 |
| 27 | or/24-26 |
| 28 | limit 27 to yr="2020-Current" |
| 29 | remove duplicates from 28 |

#### Table S1: Study- and Estimate-level Classification Decisions

| **Geographic scope** | **Definitions** |
| --- | --- |
| National | Research was conducted across multiple regions within a country |
| Sub-national | Research was conducted across multiple sub-regions or municipalities within the same state or province. |
| Local | Research was conducted in a single county, municipality, or neighborhood. |
| **Sample Frames** | **Definitions** |
| Household and community samples/samples from existing surveillance programs | Population based studies with community or house-hold based surveys derived from nonspecific members of the general population (including members selected from previously established population surveys) |
| Blood donors | Retention samples that have already been taken from banked blood derived from blood donation practices; or samples that are taken from individuals who have donated blood. |
| Residual sera | Samples that have already been taken in health care settings for a variety of reasons other than COVID serological testing; or general groups of non-COVID patients attending healthcare that do not solely belong to a specific patient population |
| Pregnant or parturient women | Samples that have already been taken from pregnant women during routine pregnancy trimester screening; or samples taken from or parturient women. |
| Persons living in slums | Samples taken from people residing in slum dwellings. |
| Representative patient population | Patient population groups where the health condition is common/endemic enough for it to constitute a meaningful sample of the general population (Malaria, HIV) |
| Multiple general populations | Samples that are composed of multiple distinct above-described populations with a single pooled seroprevalence estimate. |
| **Sampling method** | **Definitions** |
| Convenience | Non-probability sampling based on convenient sources of specimens. The sampling method should still have a clear and defined sample frame (i.e., advertisements in public spaces are not included) |
| Sequential | Non-probability sampling based on entire sampling performed within a self-determined time interval. The sampling method should still have a clear and defined sample frame, and is often drawn from convenience samples. |
| Stratified non-probability (Quota) | Non-probability sampling based on groupings of similar units. Groupings are then sampled in a non-randomized fashion, wherein nonresponders may be also ignored and replaced |
| Probability | Probability sampling based on randomized sampling methods wherein all individuals have an equal probability of being selected. |
| **Serological assays** | **Definitions** [**[2]**](https://www.zotero.org/google-docs/?Gwxuly) |
| Multiplex | Assay that simultaneously detects presence of antibodies against multiple viral antigens (i.e., spike, nucleocapsid, envelope). |
| Rapid diagnostic test | Point-of-care diagnostic platform that can be used and evaluated directly at the testing site. Typical features include short turnaround time (<15 minutes), low cost, easy interpretation of results (typically binary), and ease of use with minimal training. |
| ELISA | Enzyme linked immunosorbent assay: Wells coated with the viral antigen of interest. After the formation of immune complexes, an enzyme-linked secondary antibody followed by the enzyme’s substrate are added, which results in chemical modification of the substrate in the presence of immune complexes, which is measured colorimetrically through spectroscopy. The relative light absorption at a particular wavelength is directly proportional to the amount of labeled complexes present. |
| CLIA | Chemiluminescent immunoassay (includes chemiluminescent microparticle immunoassays [CMIA]): Stationary solid particles or wells coated with the viral antigen of interest. After the formation of immune complexes, substrate is added which results in generation of light, the intensity of which is directly proportional to the amount of labeled complexes present. |
| IFA | Immunofluorescence assay. Antibodies bind to the SARS-CoV-2 protein of interest. Fluorescent dyes are coupled to these immune complexes in order to visualize the protein of interest using microscopy or flow cytometry |
| LFIA | Lateral flow immunoassay: A liquid sample containing the analyte of interest moves without the assistance of external forces (capillary action) through various zones of polymeric strips. Antibodies of interest can bind to immobilized antigen of interest in various zones. A secondary reporter antibody (typically functionalized with gold nanoparticles) then binds to these immune complexes resulting in visual band formation for interpretation. |
| Neutralization assay | Heat treated, inactivated serum is added to cell culture (typically Vero E6 cells). Presence of neutralizing antibodies against SARS-CoV-2 is measured by the extent of binding inhibition in cell culture inoculated with the virus . Quantification is determined by the serum dilutions needed to prevent cytopathic effects in the cell monolayer. |

### S3: Supplementary Analysis

#### S3.1 Estimate prioritization

Where there were multiple primary estimates per study unrelated to time, estimates were prioritized based on adjustment (population adjustment), antibody isotypes measured (Total Ab > IgG > IgM > other), test type used (Neutralization > Chemiluminescent immunoassay [CLIA] > Enzyme-linked immunosorbent assay [ELISA] > Lateral flow immunoassay [LFIA] > other), and antibody targets measured (Multiple targets > Spike > Nucleocapsid).

When multiple estimate adjustments were available, we prioritized as follows:

1. Test unadjusted and Population adjusted
2. Both unadjusted
3. Both adjusted
4. Test adjusted and Population unadjusted

When multiple isotypes were available, we prioritized as follows:

1. Total antibodies
2. IgG OR other antibodies
3. IgG only
4. IgM OR other antibodies
5. IgG AND other antibodies
6. IgM only
7. IgM AND other antibodies
8. Other

When multiple test types were available (i.e. from use of different tests), we prioritized as follows:

1. Neutralization
2. CLIA
3. ELISA
4. LFIA
5. Other

When multiple antibody targets were available (i.e. from use of different tests), we prioritized as follows:

1. Multiple antibody targets
2. Anti - Spike
3. Anti - Nucleocapsid

If the above defined prioritization criteria led to estimates left on equal terms, we further introduced test sensitivity and specificity (highest sensitivity and specificity) prioritization criteria. If multiple estimates were present (e.g. an extraction only contained equivalent subgroup data), we further pooled estimate information to generate a single summary estimate.

**The Python code used for our automated estimate prioritization is available on GitHub:** https://github.com/serotracker/iit-backend/blob/8059e9b905395de997f28a1a2dff5def795276ad/app/utils/estimate_prioritization/estimate_prioritization.py

#### S3.2 Additional analysis description

**Cases and vaccination data:**

We drew confirmed case data from the WHO COVID-19 dashboard matched to nine days before the study mid-date to account for time from infection to seroconversion. We used data from Our World in Data dashboard to pull vaccination data, timed fourteen days before the study’s mid date. [[3]](https://www.zotero.org/google-docs/?4NpGBL)

**Meta-regression predictor definitions:**

To examine heterogeneity between study results, we constructed a Poisson generalized linear mixed-effects model with log link function using the glmer function from the lme4 package in R. [[4]](https://www.zotero.org/google-docs/?arlJB5) Categorical covariates in the meta-regression, coded as indicator variables, included geographic scope (reference: national), type of serological test (reference: CLIA assay), UN subregion (reference: Southern Africa), sample frame (reference: household and community samples), population density (reference: low density area). Continuous covariates included cumulative incidence of confirmed cases per hundred in the country of the study.

We categorized each study location as a low or high density area:

- **National studies:** countries with a population density of 75/km^2^ or higher were categorized as high density areas; others as low density areas.
- **Subnational studies:** states, regions or provinces with a population density of 150/km^2^ were categorized as high density areas; others as low density areas.
- **Local studies:** capital cities and other cities with more than 300,000 inhabitants were categorized as high density areas; others as low density areas.

Full details and sources are provided below.

##### Table S2: Population density classification decisions

**National studies**

| **Location** | **Population density (people/km^2^)** | **Source** | **High or low density area** |
| --- | --- | --- | --- |
| Zambia | 24.7 | UN World Population Prospects 2019 | Low density |
| South Africa | 48.9 | UN World Population Prospects 2019 | Low density |
| Cameroon | 56.1 | UN World Population Prospects 2019 | Low density |
| Senegal | 87.0 | UN World Population Prospects 2019 | High density |
| Kenya | 94.4 | UN World Population Prospects 2019 | High density |
| Sierra Leone | 110.5 | UN World Population Prospects 2019 | High density |
| Ethiopia | 115.0 | UN World Population Prospects 2019 | High density |
| Ghana | 136.6 | UN World Population Prospects 2019 | High density |
| Malawi | 202.9 | UN World Population Prospects 2019 | High density |
| Nigeria | 226.3 | UN World Population Prospects 2019 | High density |
| Uganda | 228.9 | UN World Population Prospects 2019 | High density |

**Subnational studies**

| **Location** | **Population density (people/km^2^)** | **Source** | **High or low density area** |
| --- | --- | --- | --- |
| Niger State, Nigeria | 57 | National Bureau of Statistics, 2009; National Population Commission, 2009 | Low density |
| Anambra State, Nigeria | 859 | National Bureau of Statistics, 2009; National Population Commission, 2009 | High density |
| Enugu State, Nigeria | 434 | National Bureau of Statistics, 2009; National Population Commission, 2009 | High density |
| Gombe State, Nigeria | 138 | National Bureau of Statistics, 2009; National Population Commission, 2009 | Low density |
| Nasarawa State, Nigeria | 65 | National Bureau of Statistics, 2009; National Population Commission, 2009 | Low density |
| Lagos State, Nigeria | 2483 | National Bureau of Statistics, 2009; National Population Commission, 2009 | High density |
| Gauteng Province, South Africa | 675 | 2011 National Census | High density |

**Local studies**

| **Location** | **Capital city** | **Source** | **High or low density area** |
| --- | --- | --- | --- |
| Ouagadougou, Burkina Faso | Yes | UN World Urbanization Prospects 2018 | High density |
| Yaoundé, Cameroon | Yes | UN World Urbanization Prospects 2018 | High density |
| Bangui, Central African Republic | Yes | UN World Urbanization Prospects 2018 | High density |
| Kinshasa, Democratic Republic of the Congo | Yes | UN World Urbanization Prospects 2018 | High density |
| Nairobi, Kenya | Yes | UN World Urbanization Prospects 2018 | High density |
| Juba, South Sudan | Yes | UN World Urbanization Prospects 2018 | High density |
| Addis Ababa, Ethiopia | Yes | UN World Urbanization Prospects 2018 | High density |
| Jimma, Ethiopia | No | UN World Urbanization Prospects 2018 | Low density |
| Dire Dawa, Ethiopia | No | UN World Urbanization Prospects 2018 | Low density |
| Harare, Zimbabwe | Yes | UN World Urbanization Prospects 2018 | High density |
| Bancoumana, Doneguebougou, and Sotuba, Mali | No | UN World Urbanization Prospects 2018 | Low density |
| Nampula, Mozambique | No | UN World Urbanization Prospects 2018 | High density |
| Tete, Mozambique | No | UN World Urbanization Prospects 2018 | High density |
| Lichinga, Mozambique | No | UN World Urbanization Prospects 2018 | Low density |
| Chimoio, Mozambique | No | UN World Urbanization Prospects 2018 | High density |
| Massinga, Mozambique | No | UN World Urbanization Prospects 2018 | Low density |
| Maxixe, Mozambique | No | UN World Urbanization Prospects 2018 | Low density |
| Pemba, Mozambique | No | UN World Urbanization Prospects 2018 | Low density |
| Agincourt, South Africa | No | UN World Urbanization Prospects 2018 | Low density |
| Klerksdorp, South Africa | No | UN World Urbanization Prospects 2018 | High density |
| Johannesburg, South Africa | No | UN World Urbanization Prospects 2018 | High density |

### S4: Supplementary Results

#### S4.1 Supplemental tables and figures

##### Table S3: Ratios of seroprevalence to cumulative incidence of confirmed cases by subregion and country, 2020 Q4-2021 Q3

| **Subregion** | **Number of national studies** | **Country** | **Range of seroprevalence to confirmed case ratios,**  **2020 Q4-2021 Q3** |
| --- | --- | --- | --- |
| **Eastern Africa** | 1 | Kenya | 243:1-243:1 |
| **Eastern Africa** | 11 | Malawi | 252:1-696:1 |
| **Eastern Africa** | 1 | Uganda | 176:1-176:1 |
| **Southern Africa** | 7 | South Africa | 10:1-25:1 |
| **Western Africa** | 4 | Ghana | 128:1-219:1 |
| **Western Africa** | 1 | Nigeria | 958:1-958:1 |
| **Western Africa** | 1 | Senegal | 299:1-299:1 |
| **Western Africa** | 1 | Sierra Leone | 57:1-57:1 |
| **Middle Africa** | 1 | Cameroon | 124:1-124:1 |

##### Table S4: Pooled seroprevalence (random-effects model) by UN sub-region, Mar 2020 - Sep 2021

Point estimates and 95% confidence intervals (error bars) are reported for each quarter, as well as the number of studies pooled (n).

| **Subregion** | **Quarter** | **N** | **Effect size** | **95% CI** | **I2** |
| --- | --- | --- | --- | --- | --- |
| **Eastern Africa** | 2020 Q2 | 6 | 0.0216 | [0.0109-0.0422] | 0.9676 |
| **Eastern Africa** | 2020 Q3 | 15 | 0.0419 | [0.0238-0.0728] | 0.9848 |
| **Eastern Africa** | 2020 Q4 | 12 | 0.1339 | [0.0782-0.2197] | 0.9942 |
| **Eastern Africa** | 2021 Q1 | 15 | 0.4204 | [0.3524-0.4916] | 0.9914 |
| **Eastern Africa** | 2021 Q2 | 4 | 0.6506 | [0.6142-0.6854] | 0.5263 |
| **Eastern Africa** | 2021 Q3 | 1 | 0.7014 | [0.646-0.7514] | NA |
| **Middle Africa** | 2020 Q4 | 3 | 0.1795 | [0.1039-0.2923] | 0.9938 |
| **Middle Africa** | 2021 Q1 | 1 | 0.7209 | [0.7029-0.7382] | NA |
| **Middle Africa** | 2021 Q3 | 1 | 0.7547 | [0.7237-0.7833] | NA |
| **Southern Africa** | 2020 Q3 | 7 | 0.1328 | [0.0591-0.2719] | 0.9692 |
| **Southern Africa** | 2020 Q4 | 5 | 0.1884 | [0.1136-0.2961] | 0.9579 |
| **Southern Africa** | 2021 Q1 | 8 | 0.3803 | [0.3125-0.4531] | 0.9736 |
| **Southern Africa** | 2021 Q2 | 3 | 0.4012 | [0.285-0.5297] | 0.9765 |
| **Southern Africa** | 2021 Q3 | 4 | 0.5606 | [0.446-0.6691] | 0.9720 |
| **Western Africa** | 2020 Q2 | 1 | 0.2541 | [0.1965-0.3217] | NA |
| **Western Africa** | 2020 Q3 | 1 | 0.0651 | [0.0563-0.0751] | NA |
| **Western Africa** | 2020 Q4 | 6 | 0.1932 | [0.1443-0.2536] | 0.9859 |
| **Western Africa** | 2021 Q1 | 3 | 0.1489 | [0.0397-0.4258] | 0.9943 |
| **Western Africa** | 2021 Q2 | 2 | 0.6719 | [0.6581-0.6854] | 0.6162 |
| **Western Africa** | 2021 Q3 | 2 | 0.7335 | [0.642-0.8086] | 0.9916 |

##### Figure S1: African Member States with seroprevalence data identified, 2020-2021 (dataset 0: all studies)

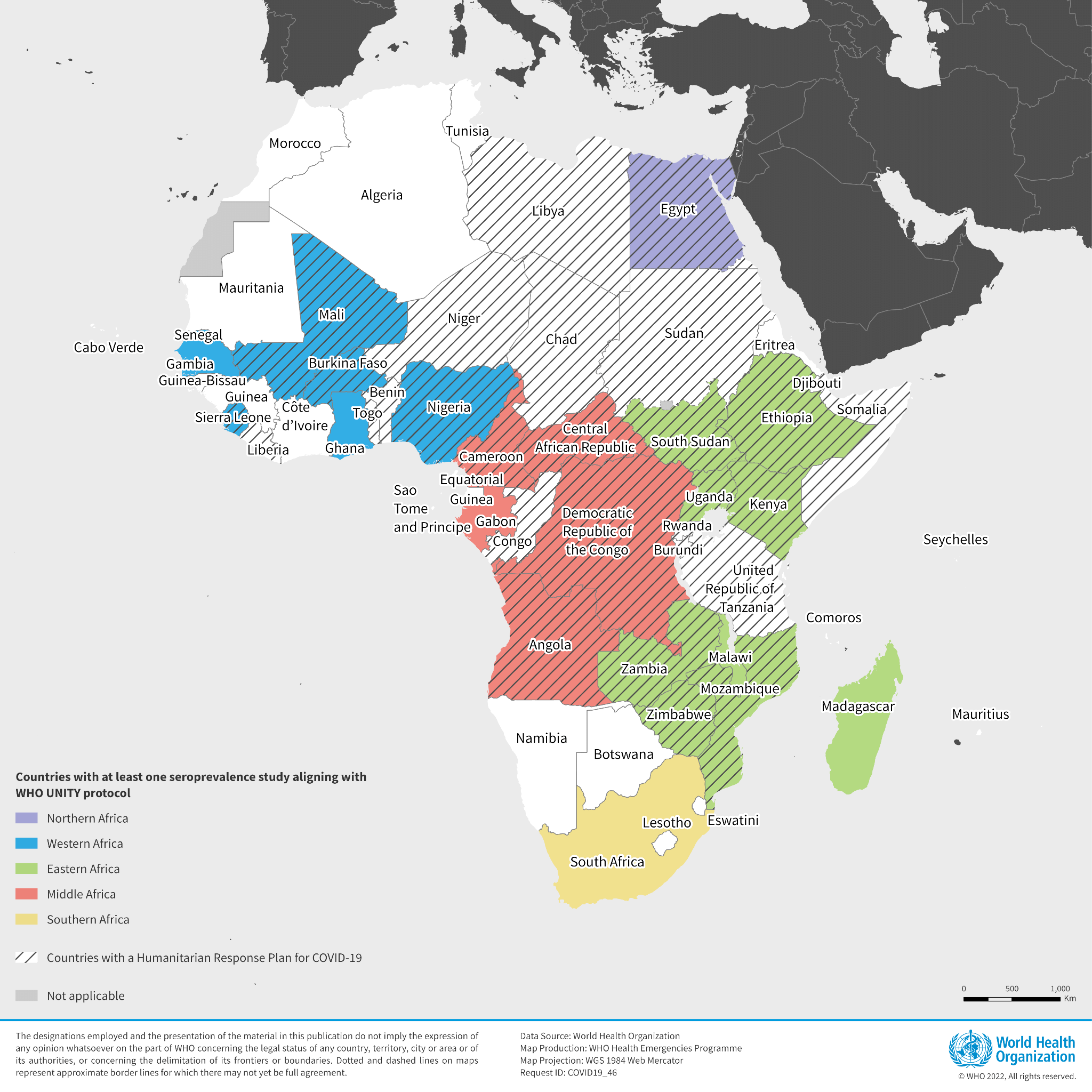

##### Figure S2: Reported seroprevalence, variants of concern, cumulative incidence, and vaccination by countries over time, Mar 2020 - Sep 2021

Plots for all countries are reported in Figure S2. **Top panel, left axis:** Shaded areas represent the relative frequency of major variants of concern (VOC) circulating, based on weekly counts of hCoV-19 genomes submitted to GISAID we have aggregated by month. Weeks with fewer than 10 total submissions in a given country were excluded from the analysis. There were no data available for South Sudan, Democratic Republic of the Congo, Burkina Faso, and Central African Republic. **Top panel, right axis:** New confirmed cases per million people, smoothed using local regression (LOESS). **Middle panel:** Each point is an individual seroprevalence study, and identical shapes represent studies originating from the same cohort or repeated cross-sectional data source. **Bottom panel, left axis:** Cumulative incidence of confirmed cases per 100 people. **Bottom panel, right axis:** Cumulative first dose of any vaccine per 100 people.

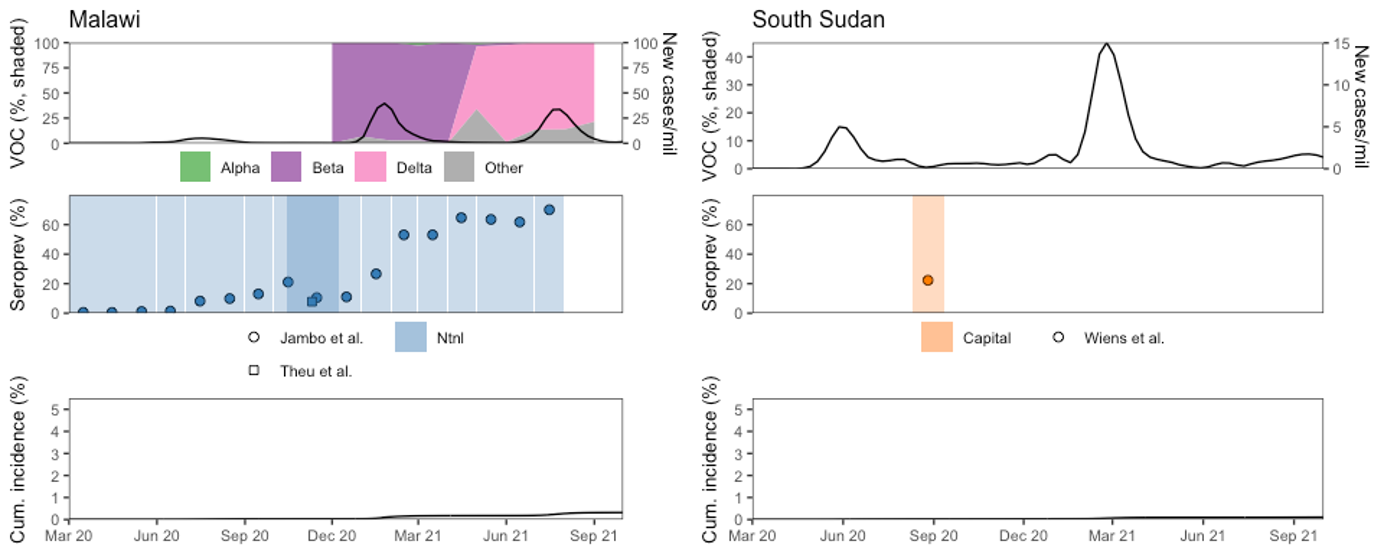

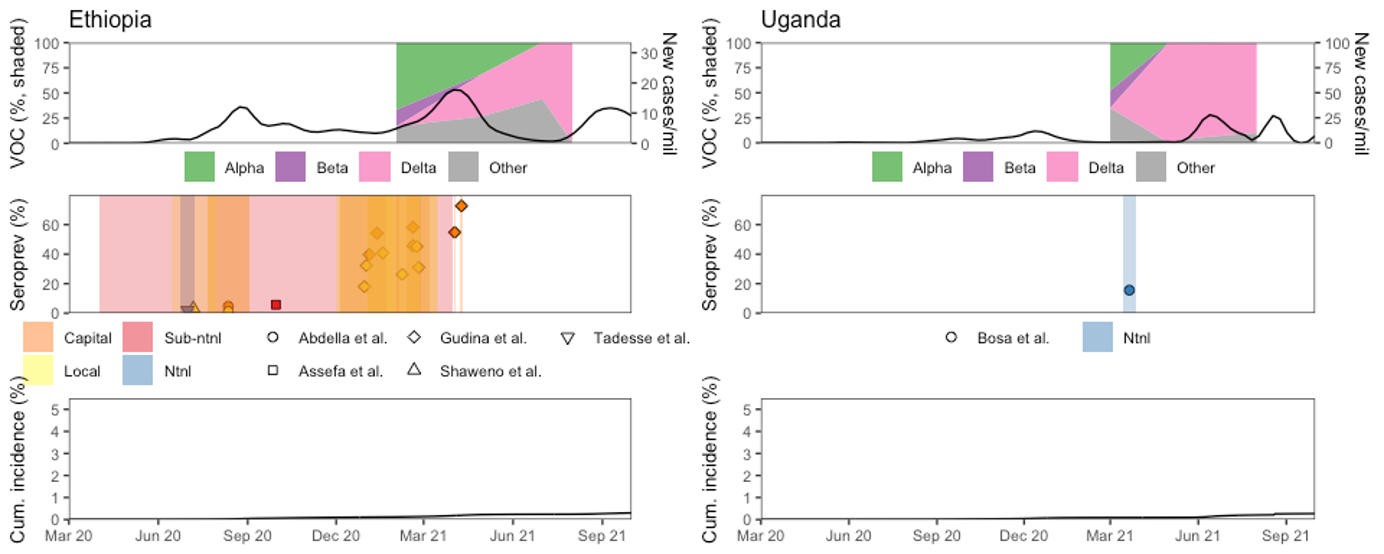

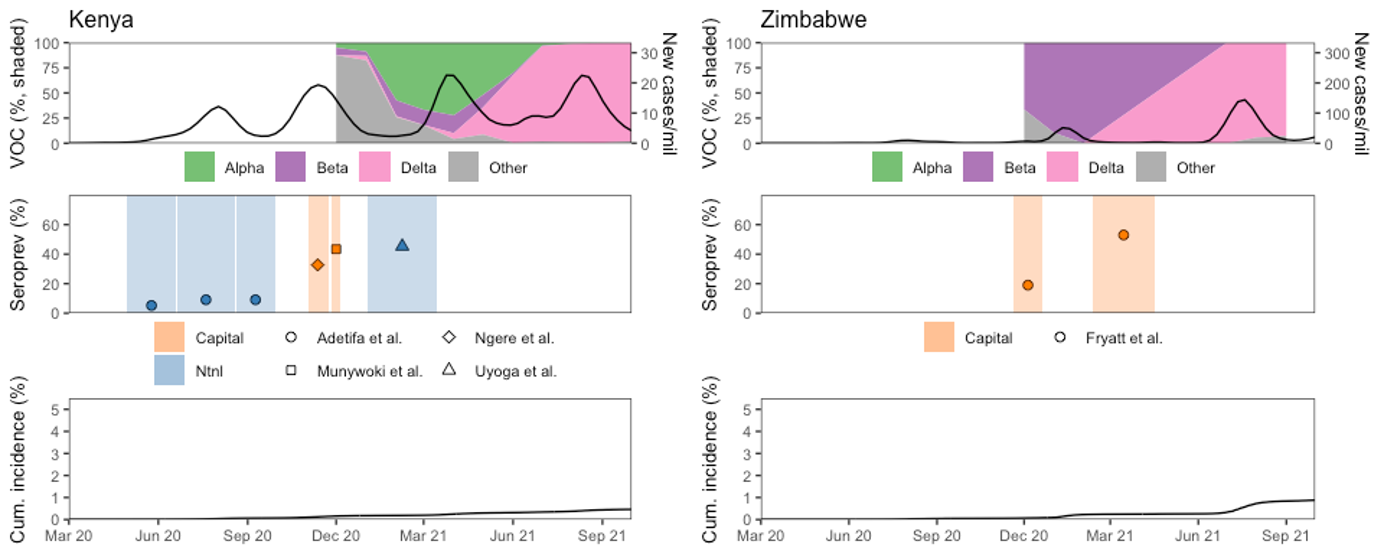

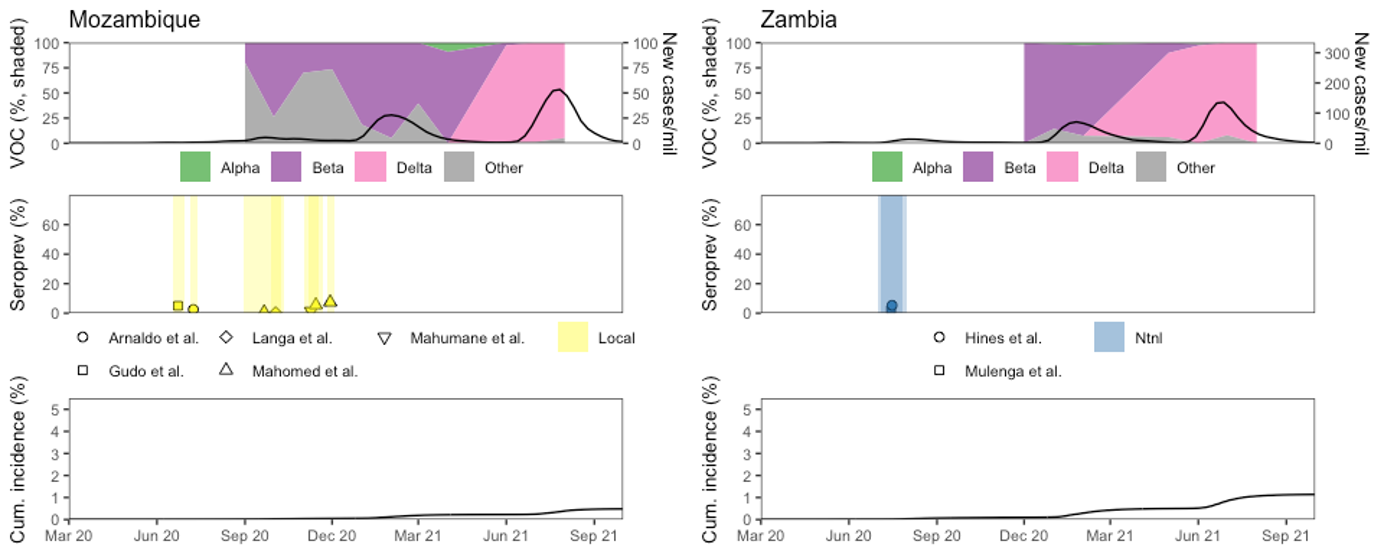

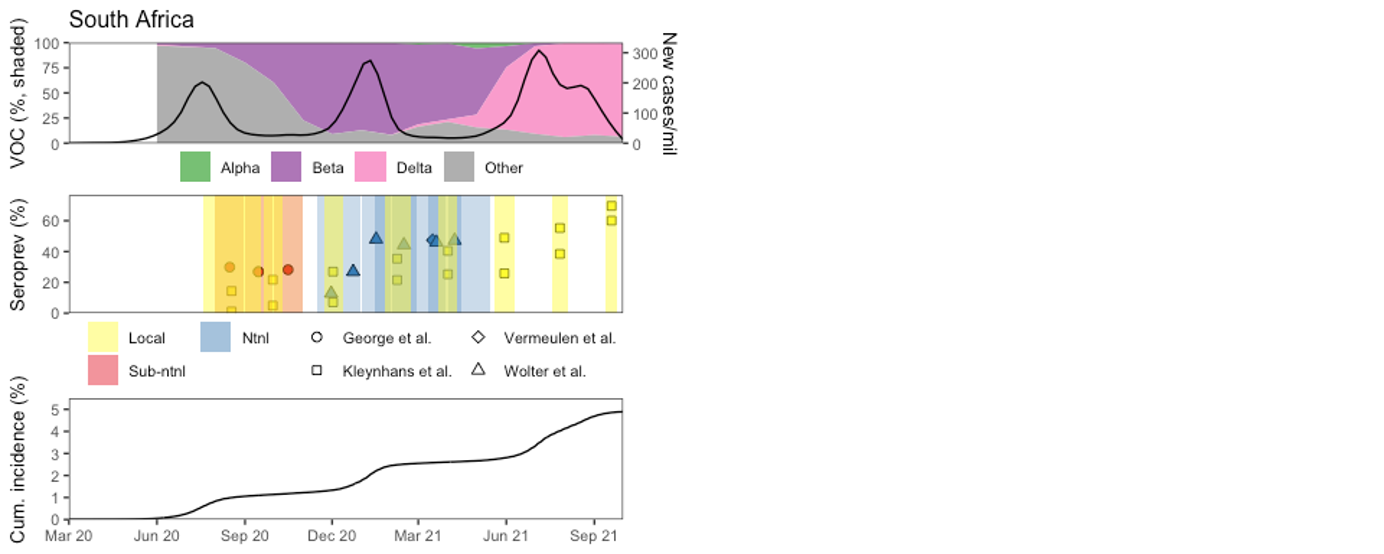

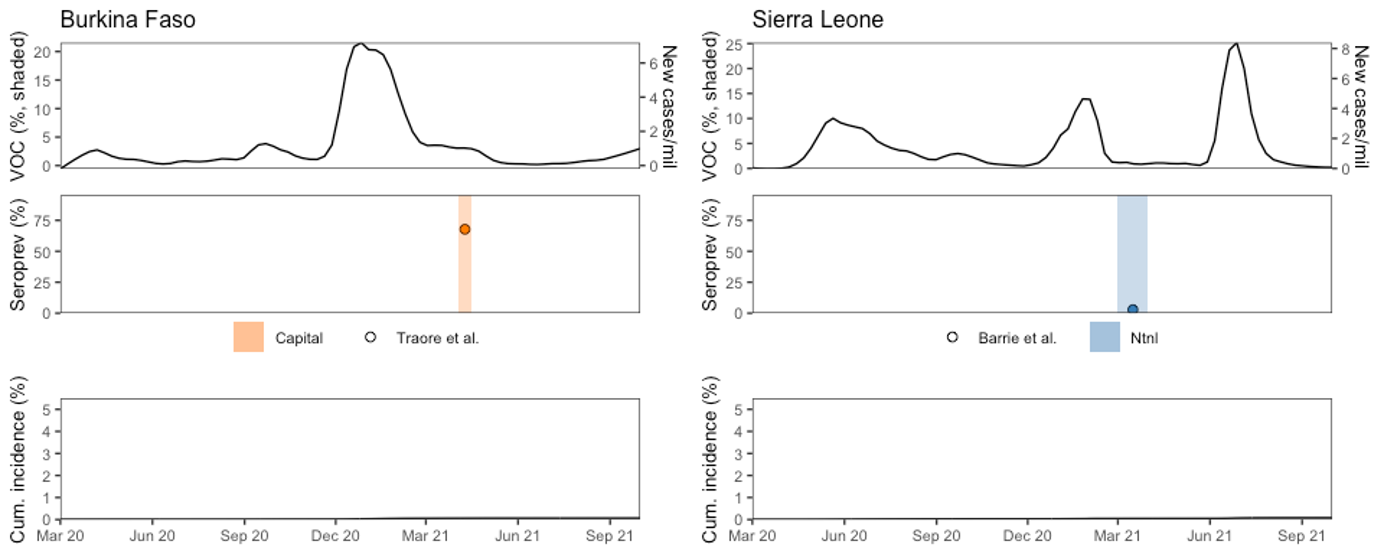

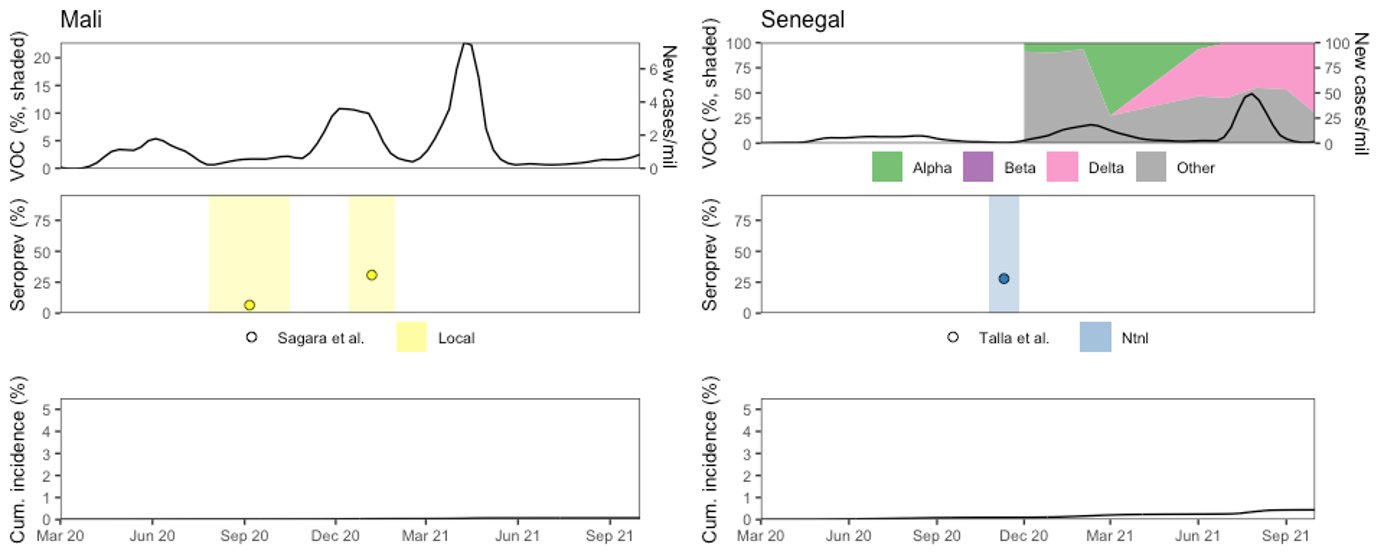

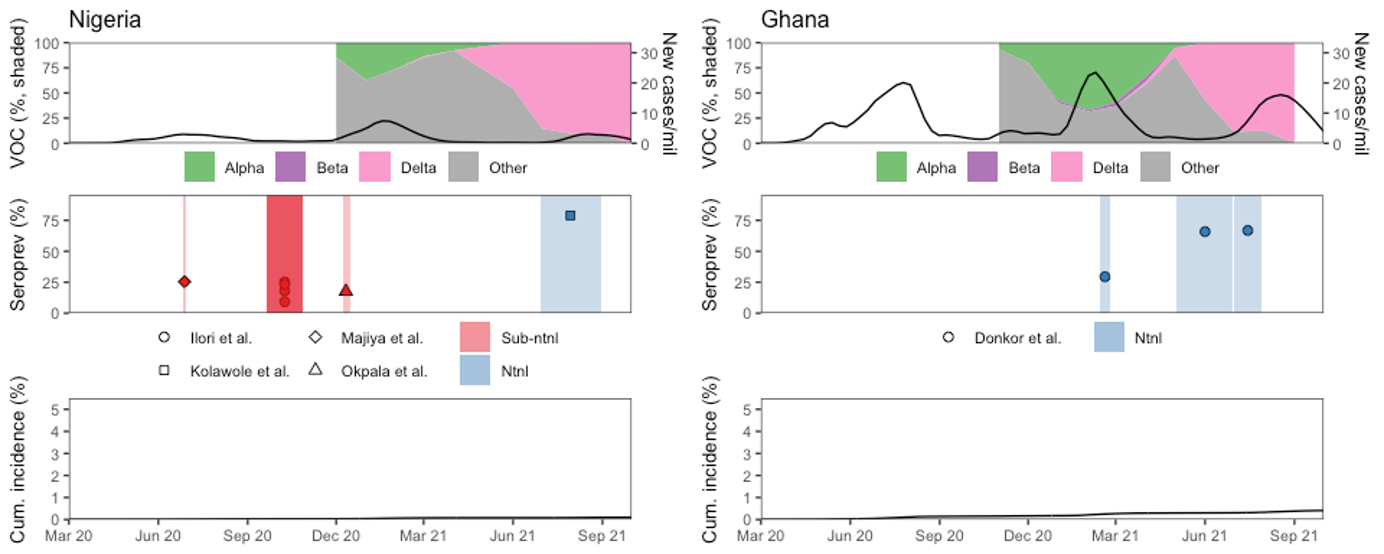

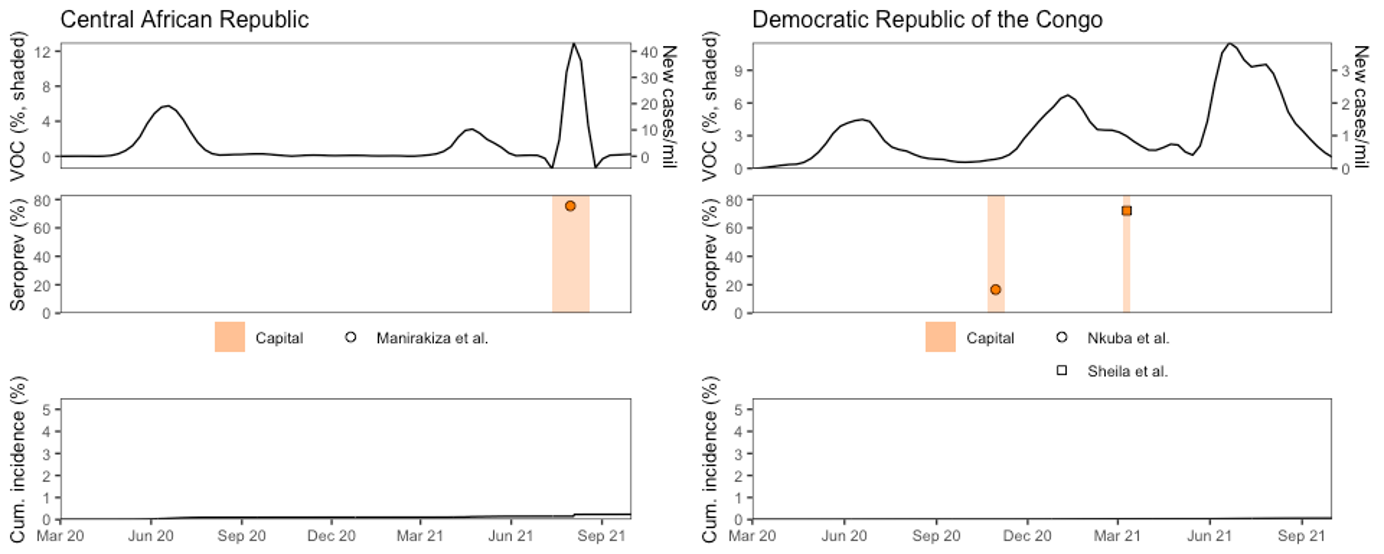

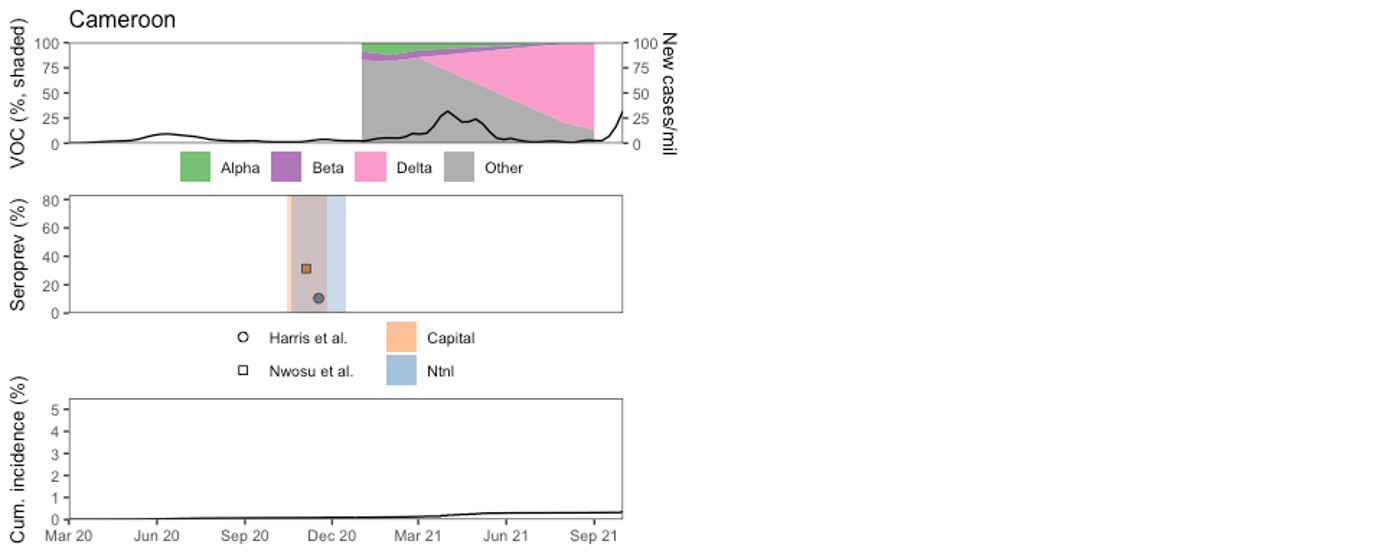

##### Figure S3: Asymptomatic seroprevalence by age and sex

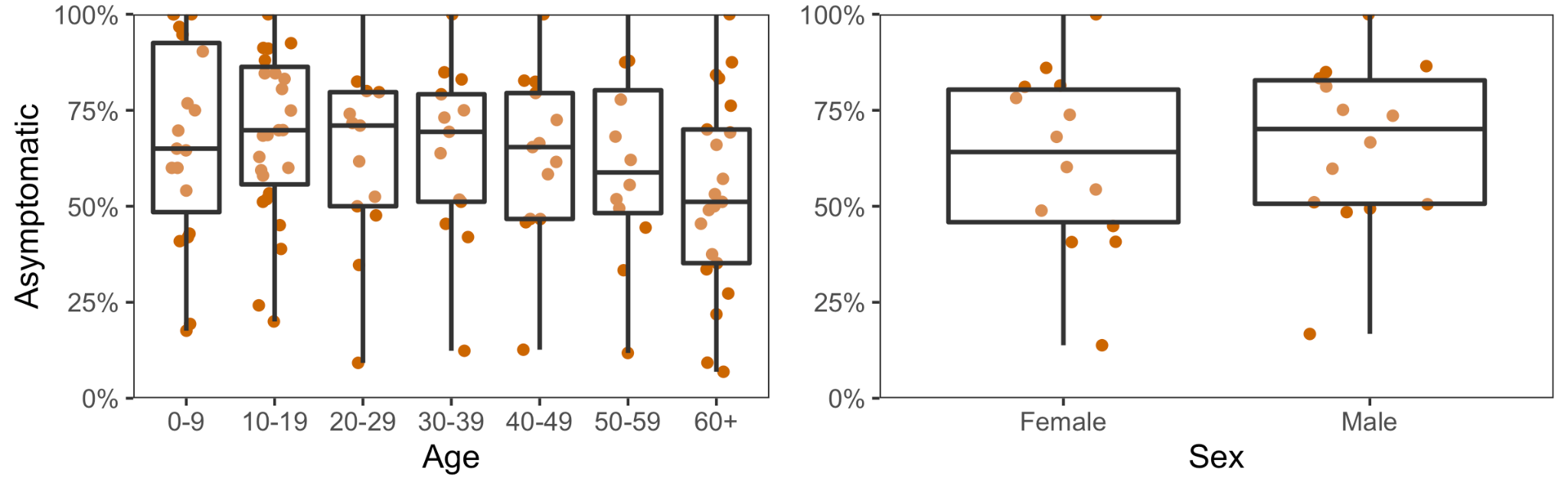

##### Figure S4: Adjusted vs. unadjusted seroprevalence data.

Each point represents a different seroprevalence study. The sensitivity and specificity values for correction were prioritized from the WHO SARS-CoV-2 Test Kit Comparative Study conducted at the NRL Australia,[[5]](https://www.zotero.org/google-docs/?gm4hlu) followed by a multicenter evaluation of 47 commercial SARS-CoV-2 immunoassays by 41 Dutch laboratories,[[6]](https://www.zotero.org/google-docs/?eNlueg) and from independent evaluations by study authors where author designed assays were used.
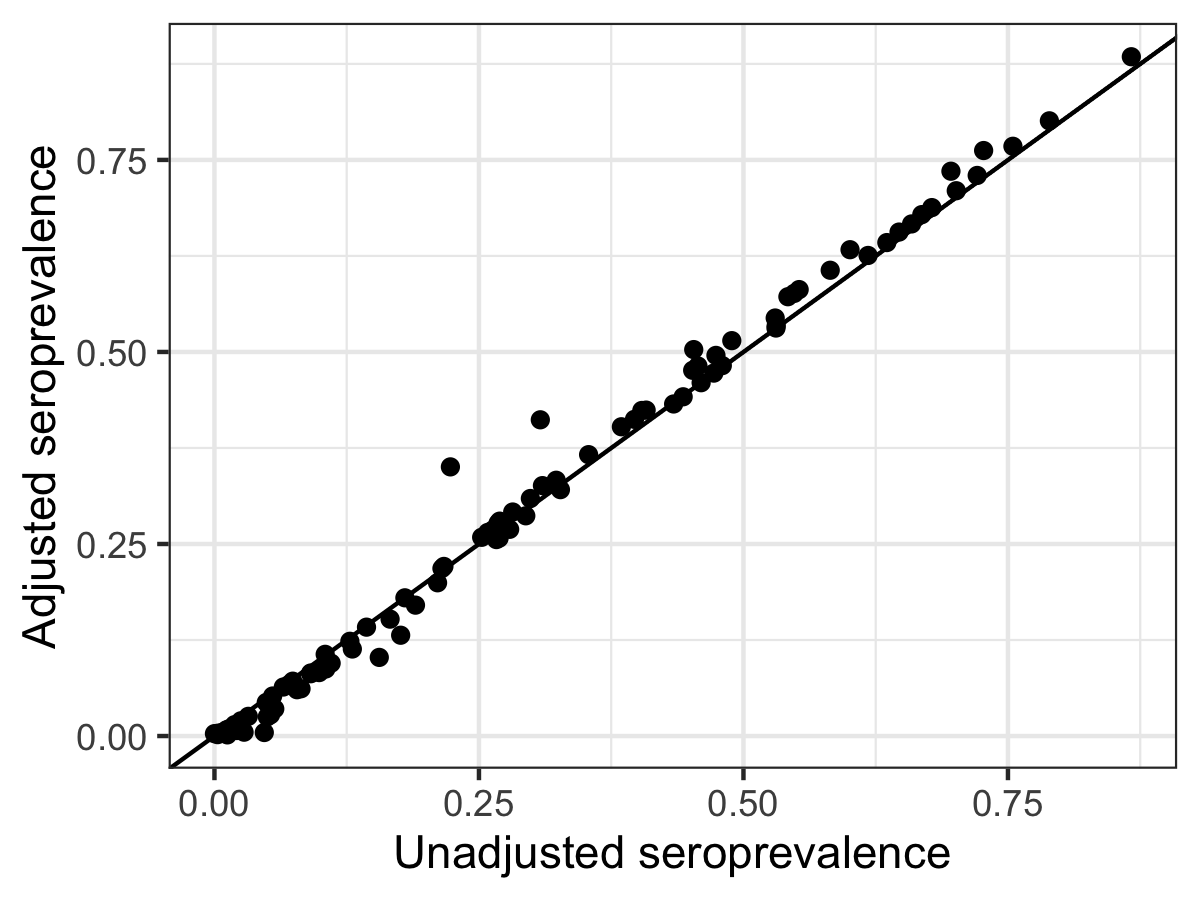

##### Figure S5: Pooled seroprevalence from infection or vaccination (random-effects model) by UN sub-region using adjusted and unadjusted seroprevalence data.

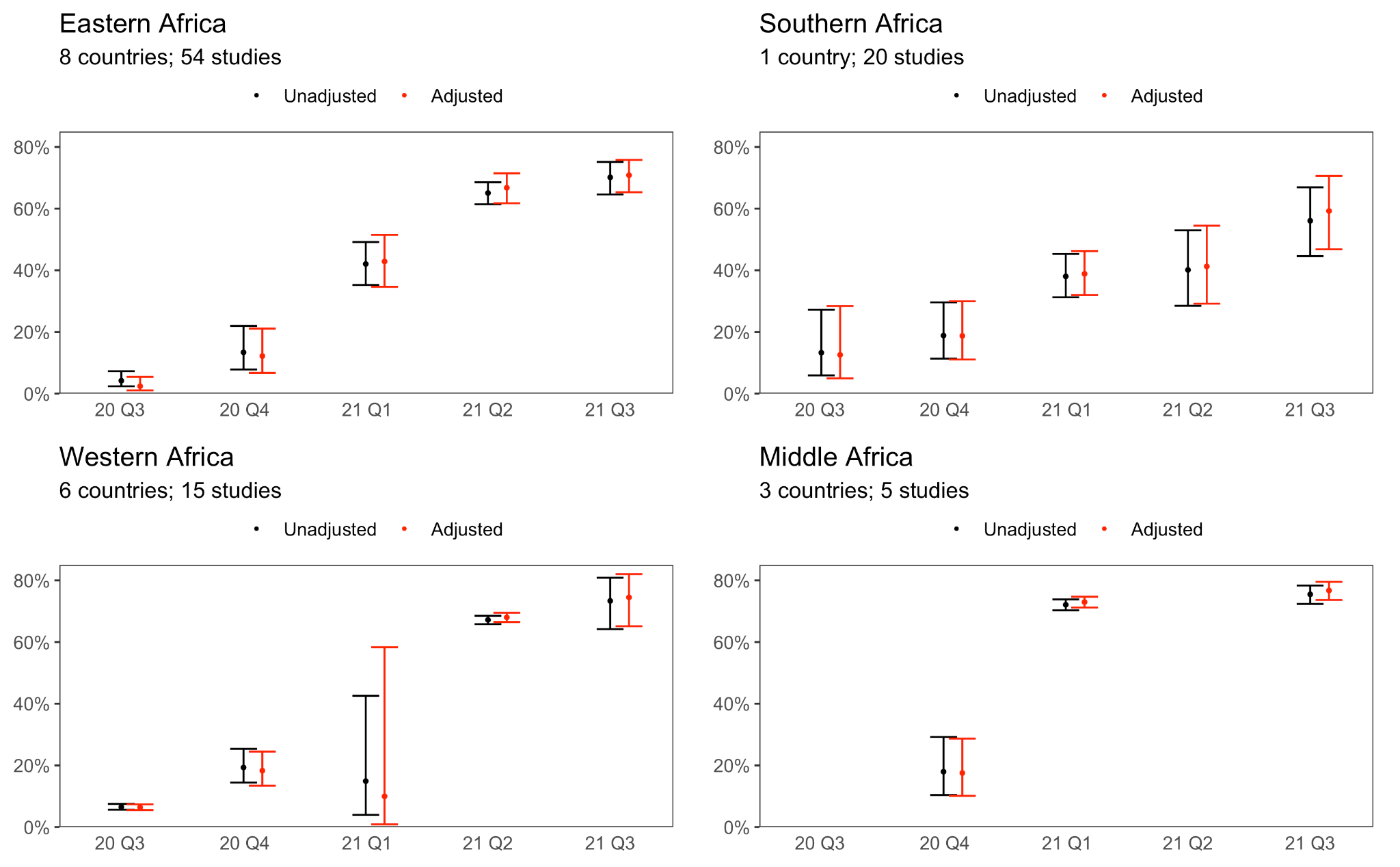

##### Figure S6: Seroprevalence to confirmed case ratios using adjusted and unadjusted seroprevalence data.

| **Subregion** | **Number of national studies** | **Country** | **Range of seroprevalence to confirmed case ratios,**  **2020 Q4-2021 Q3;**  **UNADJUSTED** | **Range of seroprevalence to confirmed case ratios,**  **2020 Q4-2021 Q3;**  **ADJUSTED** |
| --- | --- | --- | --- | --- |
| **Eastern Africa** | 1 | Kenya | 243:1-243:1 | 269:1-269:1 |
| **Eastern Africa** | 11 | Malawi | 252:1-696:1 | 194:1-667:1 |
| **Eastern Africa** | 1 | Uganda | 176:1-176:1 | 116:1-116:1 |
| **Southern Africa** | 7 | South Africa | 10:1-25:1 | 10:1-25:1 |
| **Western Africa** | 4 | Ghana | 128:1-219:1 | 125:1-221:1 |
| **Western Africa** | 1 | Nigeria | 958:1-958:1 | 972:1-972:1 |
| **Western Africa** | 1 | Senegal | 299:1-299:1 | 289:1-289:1 |
| **Western Africa** | 1 | Sierra Leone | 57:1-57:1 | 10:1-10:1 |
| **Middle Africa** | 1 | Cameroon | 124:1-124:1 | 126:1-126:1 |

##### Figure S7: Meta-regression using adjusted seroprevalence data.

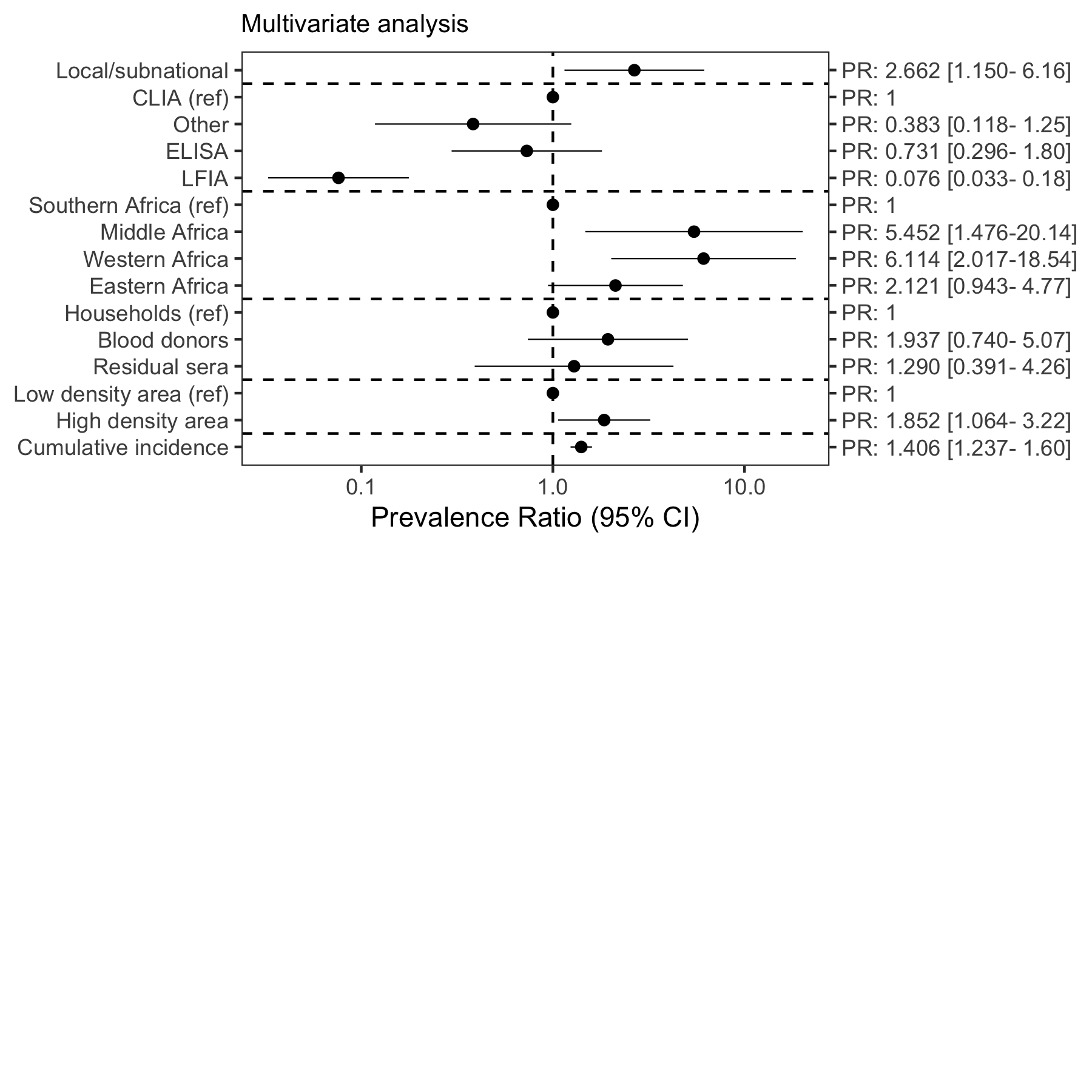

##### Figure S8: Risk of bias summary of studies included in descriptive analysis (dataset 0)

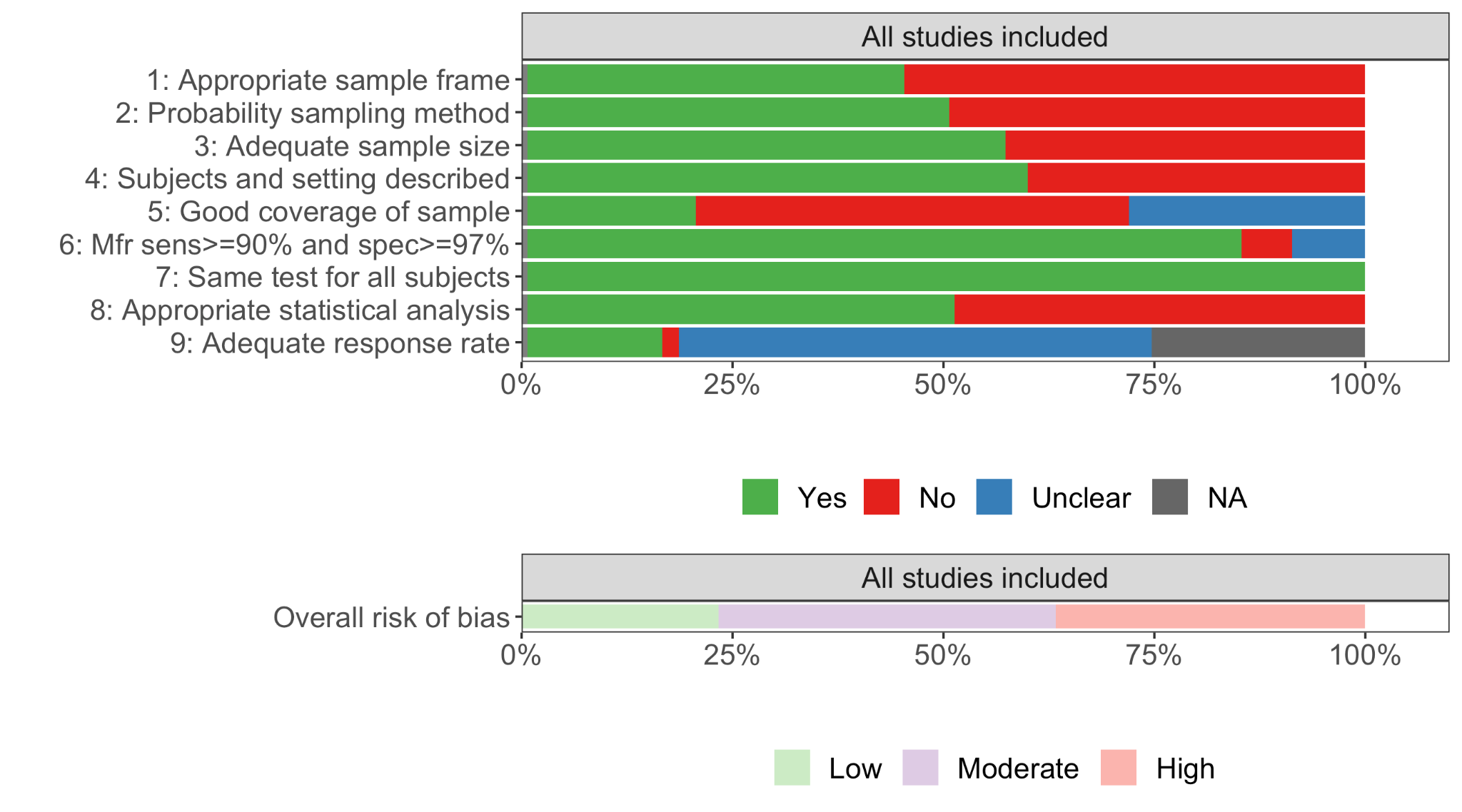

#### S4.2 Studies included in ascertainment analysis, meta-analysis, and descriptive analysis

##### Table S5. Studies used in ascertainment analysis, meta-analysis, and descriptive analysis (dataset 2: national studies rated low and moderate risk of bias).

| **Country (Location)** | **Author (Organization)** | **Sampling Dates (YMD)** | **Geographic scope** | **Overall risk of bias** | **Sample size** | **Age, sex** | **Sampling method** | **Sample frame** | **Seroprevalence (95% CI)** | **Test Manufacturers - Isotypes - Names** | **Test Sens, Spec** | **Sens, Spec Source** |
| --- | --- | --- | --- | --- | --- | --- | --- | --- | --- | --- | --- | --- |
| **Cameroon - HRP** | Tiffany Harris (ICAP at Columbia University)[1] | 2020-10-20 - 2020-12-16 | National | Moderate | 9,332 | Multiple groups, All | Convenience | Household and community samples | 10.5% (95% CI 9.1-12.0%) | Abbott Laboratories - IgG - Abbott Architect SARS-CoV-2 IgG;Beijing Wantai Biological - IgG, IgM, IgA - Wantai SARS-CoV-2 Total Ab ELISA | 0.957, 1 | Complex test algorithm: Manual calculation |
| **Ethiopia - HRP** | Tadesse (Federal Ministry of Health)[2] | 2020-06-24 - 2020-07-08 | National | Low | 16,932 | Multiple groups, All | Probability | Household and community samples | 1.9% (95% CI 1.7-2.1%) | Abbott Laboratories - IgG - Abbott Architect SARS-CoV-2 IgG | 0.99, 1 | Validated by manufacturer |
| **Ghana** | Irene Owusu Donkor (Noguchi Memorial Institute for Medical Rsearch)[3] | 2021-12-12 - 2021-12-19 | National | Moderate | 1,027 | Multiple groups, All | Probability | Household and community samples | 86.7% (95% CI 84.4-88.7%) | Beijing Wantai Biological - IgG, IgM, IgA - Wantai SARS-CoV-2 Total Ab ELISA | 0.967, 0.975 | Validated by manufacturer |
| **Ghana** | Irene Owusu Donkor (Noguchi Memorial Institute for Medical Rsearch)[3] | 2021-07-01 - 2021-07-29 | National | Moderate | 2,275 | Multiple groups, All | Probability | Household and community samples | 66.9% (95% CI 64.9-68.8%) | Beijing Wantai Biological - IgG, IgM, IgA - Wantai SARS-CoV-2 Total Ab ELISA | 0.967, 0.975 | Validated by manufacturer |
| **Ghana** | Irene Owusu Donkor (Noguchi Memorial Institute for Medical Rsearch)[3] | 2021-05-03 - 2021-06-30 | National | Moderate | 2,234 | Multiple groups, All | Probability | Household and community samples | 65.9% (95% CI 63.9-67.9%) | Beijing Wantai Biological - IgG, IgM, IgA - Wantai SARS-CoV-2 Total Ab ELISA | 0.967, 0.975 | Validated by manufacturer |
| **Ghana** | Irene Owusu Donkor (Noguchi Memorial Institute for Medical Research)[3] | 2021-02-13 - 2021-02-23 | National | Moderate | 401 | Multiple groups, All | Probability | Household and community samples | 29.4% (95% CI 25-34.2%) | Beijing Wantai Biological - IgG, IgM, IgA - Wantai SARS-CoV-2 Total Ab ELISA | 0.967, 0.975 | Validated by manufacturer |
| **Kenya - HRP** | Uyoga (KEMRI-Wellcome Trust Research Programme)[4] | 2021-01-03 - 2021-03-15 | National | Moderate | 3,018 | Multiple groups, All | Convenience | Blood donors | 45.3% | NA - IgG - Author designed (ELISA) -Spike | 0.927, 0.99 | Validated by independent authors/third party/non-developers |
| **Kenya - HRP** | Adetifa (KEMRI-Wellcome Trust Research Programme)[5] | 2020-08-20 - 2020-09-30 | National | Moderate | 3,723 | Adults (18-64 years), All | Sequential | Blood donors | 9.1% | NA - IgG - Author designed (ELISA) -Spike | 0.927, 0.99 | Validated by independent authors/third party/non-developers |
| **Malawi** | Jambo (Malawi-Liverpool-Wellcome Trust Clinical Research programme)[6] | 2021-07-01 - 2021-07-31 | National | Low | 288 | Multiple groups, All | Probability | Blood donors | 70.1% | Beijing Wantai Biological - IgG, IgM, IgA - Wantai SARS-CoV-2 Total Ab ELISA;Omega diagnostics - IgG - COVID-19 IgG ELISA | 0.928, 1 | Complex test algorithm: Manual calculation |
| **Malawi** | Jambo (Malawi-Liverpool-Wellcome Trust Clinical Research programme)[6] | 2021-06-01 - 2021-06-30 | National | Low | 319 | Multiple groups, All | Probability | Blood donors | 61.8% | Beijing Wantai Biological - IgG, IgM, IgA - Wantai SARS-CoV-2 Total Ab ELISA;Omega diagnostics - IgG - COVID-19 IgG ELISA | 0.928, 1 | Complex test algorithm: Manual calculation |
| **Malawi** | Jambo (Malawi-Liverpool-Wellcome Trust Clinical Research programme)[6] | 2021-05-01 - 2021-05-31 | National | Low | 298 | Multiple groups, All | Probability | Blood donors | 63.6% | Beijing Wantai Biological - IgG, IgM, IgA - Wantai SARS-CoV-2 Total Ab ELISA;Omega diagnostics - IgG - COVID-19 IgG ELISA | 0.928, 1 | Complex test algorithm: Manual calculation |
| **Malawi** | Jambo (Malawi-Liverpool-Wellcome Trust Clinical Research programme)[6] | 2021-04-01 - 2021-04-30 | National | Low | 296 | Multiple groups, All | Probability | Blood donors | 64.7% | Beijing Wantai Biological - IgG, IgM, IgA - Wantai SARS-CoV-2 Total Ab ELISA;Omega diagnostics - IgG - COVID-19 IgG ELISA | 0.928, 1 | Complex test algorithm: Manual calculation |
| **Malawi** | Jambo (Malawi-Liverpool-Wellcome Trust Clinical Research programme)[6] | 2021-03-01 - 2021-03-31 | National | Low | 298 | Multiple groups, All | Probability | Blood donors | 53.1% | Beijing Wantai Biological - IgG, IgM, IgA - Wantai SARS-CoV-2 Total Ab ELISA;Omega diagnostics - IgG - COVID-19 IgG ELISA | 0.928, 1 | Complex test algorithm: Manual calculation |
| **Malawi** | Jambo (Malawi-Liverpool-Wellcome Trust Clinical Research programme)[6] | 2021-02-01 - 2021-02-28 | National | Low | 365 | Multiple groups, All | Probability | Blood donors | 53.1% | Beijing Wantai Biological - IgG, IgM, IgA - Wantai SARS-CoV-2 Total Ab ELISA;Omega diagnostics - IgG - COVID-19 IgG ELISA | 0.928, 1 | Complex test algorithm: Manual calculation |
| **Malawi** | Jambo (Malawi-Liverpool-Wellcome Trust Clinical Research programme)[6] | 2021-01-01 - 2021-01-31 | National | Low | 303 | Multiple groups, All | Probability | Blood donors | 26.7% | Beijing Wantai Biological - IgG, IgM, IgA - Wantai SARS-CoV-2 Total Ab ELISA;Omega diagnostics - IgG - COVID-19 IgG ELISA | 0.928, 1 | Complex test algorithm: Manual calculation |
| **Malawi** | Jambo (Malawi-Liverpool-Wellcome Trust Clinical Research programme)[6] | 2020-12-01 - 2020-12-31 | National | Low | 263 | Multiple groups, All | Probability | Blood donors | 11% | Beijing Wantai Biological - IgG, IgM, IgA - Wantai SARS-CoV-2 Total Ab ELISA;Omega diagnostics - IgG - COVID-19 IgG ELISA | 0.928, 1 | Complex test algorithm: Manual calculation |
| **Malawi** | Jambo (Malawi-Liverpool-Wellcome Trust Clinical Research programme)[6] | 2020-11-01 - 2020-11-30 | National | Low | 365 | Multiple groups, All | Probability | Blood donors | 10.5% | Beijing Wantai Biological - IgG, IgM, IgA - Wantai SARS-CoV-2 Total Ab ELISA;Omega diagnostics - IgG - COVID-19 IgG ELISA | 0.928, 1 | Complex test algorithm: Manual calculation |
| **Malawi** | Matthews Kagoli/Joe Theu (Public Health Institute of Malawi(PHIM) MOH)[7] | 2020-10-14 - 2020-12-08 | National | Low | 4,261 | Multiple groups, All | Probability | Household and community samples | 7.8% | Beijing Wantai Biological - IgG, IgM, IgA - Wantai SARS-CoV-2 Total Ab ELISA | 0.967, 0.975 | Validated by manufacturer |
| **Malawi** | Jambo (Malawi-Liverpool-Wellcome Trust Clinical Research programme)[6] | 2020-10-01 - 2020-10-31 | National | Low | 324 | Multiple groups, All | Probability | Blood donors | 21.1% | Beijing Wantai Biological - IgG, IgM, IgA - Wantai SARS-CoV-2 Total Ab ELISA;Omega diagnostics - IgG - COVID-19 IgG ELISA | 0.928, 1 | Complex test algorithm: Manual calculation |
| **Malawi** | Jambo (Malawi-Liverpool-Wellcome Trust Clinical Research programme)[6] | 2020-09-01 - 2020-09-30 | National | Low | 309 | Multiple groups, All | Probability | Blood donors | 13% | Beijing Wantai Biological - IgG, IgM, IgA - Wantai SARS-CoV-2 Total Ab ELISA;Omega diagnostics - IgG - COVID-19 IgG ELISA | 0.928, 1 | Complex test algorithm: Manual calculation |
| **Malawi** | Jambo (Malawi-Liverpool-Wellcome Trust Clinical Research programme)[6] | 2020-08-01 - 2020-08-31 | National | Low | 192 | Multiple groups, All | Probability | Blood donors | 9.9% | Beijing Wantai Biological - IgG, IgM, IgA - Wantai SARS-CoV-2 Total Ab ELISA;Omega diagnostics - IgG - COVID-19 IgG ELISA | 0.928, 1 | Complex test algorithm: Manual calculation |
| **Malawi** | Jambo (Malawi-Liverpool-Wellcome Trust Clinical Research programme)[6] | 2020-07-01 - 2020-07-31 | National | Low | 204 | Multiple groups, All | Probability | Blood donors | 8.2% | Beijing Wantai Biological - IgG, IgM, IgA - Wantai SARS-CoV-2 Total Ab ELISA;Omega diagnostics - IgG - COVID-19 IgG ELISA | 0.928, 1 | Complex test algorithm: Manual calculation |
| **Malawi** | Jambo (Malawi-Liverpool-Wellcome Trust Clinical Research programme)[6] | 2020-06-01 - 2020-06-30 | National | Low | 242 | Multiple groups, All | Probability | Blood donors | 1.4% | Beijing Wantai Biological - IgG, IgM, IgA - Wantai SARS-CoV-2 Total Ab ELISA;Omega diagnostics - IgG - COVID-19 IgG ELISA | 0.928, 1 | Complex test algorithm: Manual calculation |
| **Malawi** | Jambo (Malawi-Liverpool-Wellcome Trust Clinical Research programme)[6] | 2020-05-01 - 2020-05-31 | National | Low | 194 | Multiple groups, All | Probability | Blood donors | 1.1% | Beijing Wantai Biological - IgG, IgM, IgA - Wantai SARS-CoV-2 Total Ab ELISA;Omega diagnostics - IgG - COVID-19 IgG ELISA | 0.928, 1 | Complex test algorithm: Manual calculation |
| **Malawi** | Jambo (Malawi-Liverpool-Wellcome Trust Clinical Research programme)[6] | 2020-04-01 - 2020-04-30 | National | Low | 205 | Multiple groups, All | Probability | Blood donors | 0.5% | Beijing Wantai Biological - IgG, IgM, IgA - Wantai SARS-CoV-2 Total Ab ELISA;Omega diagnostics - IgG - COVID-19 IgG ELISA | 0.928, 1 | Complex test algorithm: Manual calculation |
| **Malawi** | Jambo (Malawi-Liverpool-Wellcome Trust Clinical Research programme)[6] | 2020-01-01 - 2020-01-31 | National | Low | 227 | Multiple groups, All | Probability | Blood donors | 0% | Beijing Wantai Biological - IgG, IgM, IgA - Wantai SARS-CoV-2 Total Ab ELISA;Omega diagnostics - IgG - COVID-19 IgG ELISA | 0.928, 1 | Complex test algorithm: Manual calculation |
| **Malawi** | Jambo (Malawi-Liverpool-Wellcome Trust Clinical Research programme)[6] | 2020-02-01 - 2020-02-29 | National | Low | 194 | Multiple groups, All | Probability | Blood donors | 0.3% | Beijing Wantai Biological - IgG, IgM, IgA - Wantai SARS-CoV-2 Total Ab ELISA;Omega diagnostics - IgG - COVID-19 IgG ELISA | 0.928, 1 | Complex test algorithm: Manual calculation |
| **Malawi** | Jambo (Malawi-Liverpool-Wellcome Trust Clinical Research programme)[6] | 2020-03-01 - 2020-03-31 | National | Low | 199 | Multiple groups, All | Probability | Blood donors | 0.5% | Beijing Wantai Biological - IgG, IgM, IgA - Wantai SARS-CoV-2 Total Ab ELISA;Omega diagnostics - IgG - COVID-19 IgG ELISA | 0.928, 1 | Complex test algorithm: Manual calculation |
| **Nigeria - HRP** | Olatunji Kolawole (Ministerial Expert Advisory Committee on COVID-19- Health Sector Response)[8] | 2021-06-29 - 2021-08-31 | National | Moderate | 4,904 | Multiple groups, All | Probability | Residual sera | 78.9% (95% CI 77.7-80%) | Beijing Wantai Biological - IgG, IgM, IgA - Wantai SARS-CoV-2 Total Ab ELISA | 0.967, 0.975 | Validated by manufacturer |
| **Senegal** | Talla (Institut Pasteur de Dakar)[9] | 2020-10-25 - 2020-11-26 | National | Low | 1,422 | Multiple groups, All | Probability | Household and community samples | 27.9% (95% CI 25.6-30.3%) | Omega diagnostics - IgG - COVID-19 IgG ELISA , ID.Vet - IgG - ID Screen , Beijing Wantai Biological - IgG, IgM, IgA - Wantai SARS-CoV-2 Total Ab ELISA | N/A | Complex algorithm: Manual review |
| **Sierra Leone - HRP** | Barrie (Harvard Medical School)[10] | 2021-03-01 - 2021-03-31 | National | Low | 1,893 | Multiple groups, All | Probability | Household and community samples | 2.8% (95% CI 2.1-3.6%) | Hangzhou Biotest Biotech Co., Ltd - IgG, IgM - RightSign COVID-19 IgG/IgM Rapid Test Cassette | 0.914, 1 | Validated by manufacturer |
| **South Africa** | Wolter (National Health Laboratory Service)[11] | 2021-04-01 - 2021-04-15 | National | Moderate | 1,849 | Multiple groups, All | Probability | Household and community samples | 47.2% | Beijing Wantai Biological - IgG, IgM, IgA - Wantai SARS-CoV-2 Total Ab ELISA , Roche Diagnostics - IgG, IgM, IgA - Elecsys® Anti‐SARS‐CoV‐2 (N) | 1, 0.97 | Complex algorithm: manual calculation |
| **South Africa** | Vermeulen (South African National Blood Service)[12] | 2021-01-15 - 2021-05-15 | National | Moderate | 16,762 | Multiple groups, All | Sequential | Blood donors | 47.4% | Roche Diagnostics - IgG, IgM, IgA - Elecsys® Anti‐SARS‐CoV‐2 (N) | 1, 0.995 | Validated by manufacturer |
| **Uganda - HRP** | Dr. Henry Kyobe Bosa (Ministry of Health/ UPDF)[55] | 2021-03-14 - 2021-03-28 | National | Moderate | 7,029 | Multiple groups, All | Probability | Household and community samples | 15.6% (95% CI 14.7-16.4%) | Hangzhou Biotest Biotech Co., Ltd - IgG, IgM - RightSign COVID-19 IgG/IgM Rapid Test Cassette | 0.914, 1 | Validated by manufacturer |
| **Zambia - HRP** | Mulenga (Zambia Ministry of Health)[13] | 2020-07-04 - 2020-07-27 | National | Low | 2,704 | Multiple groups, All | Probability | Household and community samples | 2.1% | EUROIMMUN AG - IgG - Anti-SARS-CoV-2 ELISA (IgG) | 0.9, 1 | Validated by manufacturer |
| **Zambia - HRP** | Hines (US Centers for Disease Control and Prevention)[14] | 2020-07-02 - 2020-07-31 | National | Moderate | 1,657 | Multiple groups, All | Convenience | Residual sera | 5.3% (95% CI 4.3-6.5%) | EUROIMMUN AG - IgG - Anti-SARS-CoV-2 ELISA (IgG) | 0.9, 1 | Validated by manufacturer |

##### Table S6. Studies used in meta-analysis and descriptive analysis (dataset 1: local and sub-national studies rated low and moderate risk of bias. For national studies in dataset 1, see Table S5).

| **Country (Location)** | **Author (Organization)** | **Sampling Dates (YMD)** | **Geographic scope** | **Overall risk of bias** | **Sample size** | **Age, sex** | **Sampling method** | **Sample frame** | **Seroprevalence (95% CI)** | **Test Manufacturers - Isotypes - Names** | **Test Sens, Spec** | **Sens, Spec Source** |
| --- | --- | --- | --- | --- | --- | --- | --- | --- | --- | --- | --- | --- |
| **Burkina Faso - HRP (Ouagadougou & Bobo-Dioulasso)** | Traore (National Institute of Public Health)[15] | 2021-04-03 - 2021-04-16 | Local | Moderate | 5,240 | Multiple groups, All | Probability | Household and community samples | 67.8% (95% CI 66.5-69.1%) | Beijing Wantai Biological - IgG, IgM, IgA - Wantai SARS-CoV-2 Total Ab ELISA | 0.967, 0.975 | Validated by manufacturer |
| **Cameroon - HRP (Yaoundé)** | Nwosu (Hopital Central de Yaounde)[16] | 2020-10-14 - 2020-11-26 | Local | Low | 971 | Multiple groups, All | Probability | Household and community samples | 31.3% | Abbott Laboratories - IgG, IgM - Panbio COVID-19 IgG/IgM rapid test device | 0.978, 0.928 | Validated by manufacturer |
| **Central African Republic - HRP (Bangui, the capital of the Central African Republic)** | Alexandre Manirakiza (Institut Pasteur of Bangui)[17] | 2021-07-12 - 2021-08-20 | Local | Moderate | 799 | Multiple groups, All | Probability | Household and community samples | 75.5% (95% CI 72.3-78.4%) | Beijing Wantai Biological - IgG, IgM, IgA - Wantai SARS-CoV-2 Total Ab ELISA | 0.967, 0.975 | Validated by manufacturer |
| **Democratic Republic of the Congo - HRP (All the 35 health districts of Kinshasa)** | Makiala Mandanda Sheila (Institut National de Recherche Biomédicale)[18] | 2021-03-06 - 2021-03-14 | Subnational | Moderate | 2,476 | Multiple groups, All | Probability | Household and community samples | 72.1% (95% CI 70.3-73.9%) | Beijing Wantai Biological - IgG, IgM, IgA - Wantai SARS-CoV-2 Total Ab ELISA | 0.967, 0.975 | Validated by manufacturer |
| **Democratic Republic of the Congo - HRP (Kinshasa)** | Nkuba (Institut National de Recherche Biomédicale)[19] | 2020-10-22 - 2020-11-08 | Local | Low | 1,080 | Multiple groups, All | Probability | Household and community samples | 16.6% | NA - - Author designed (Luminex) | 1, 0.997 | Validated by developers |
| **Ethiopia - HRP (Addis Ababa)** | Gudina (University of Munich)[20] | 2021-04-08 - 2021-04-10 | Local | Moderate | 176 | Multiple groups, All | Convenience | Household and community samples | 72.7% (95% CI 64.9-78.6%) | Roche Diagnostics - IgG, IgM, IgA - Elecsys® Anti‐SARS‐CoV‐2 (N) | 1, 0.995 | Validated by manufacturer |
| **Ethiopia - HRP (Addis Ababa)** | Gudina (University of Munich)[20] | 2021-04-01 - 2021-04-03 | Local | Moderate | 188 | Multiple groups, All | Convenience | Household and community samples | 54.8% (95% CI 47.4-62%) | Roche Diagnostics - IgG, IgM, IgA - Elecsys® Anti‐SARS‐CoV‐2 (N) | 1, 0.995 | Validated by manufacturer |
| **Ethiopia - HRP (Jimma)** | Gudina (University of Munich)[20] | 2021-02-04 - 2021-03-16 | Local | Moderate | 100 | Multiple groups, All | Convenience | Household and community samples | 31% (95% CI 22.1-41%) | Roche Diagnostics - IgG, IgM, IgA - Elecsys® Anti‐SARS‐CoV‐2 (N) | 1, 0.995 | Validated by manufacturer |
| **Ethiopia - HRP (Jimma)** | Gudina (University of Munich)[20] | 2021-02-02 - 2021-03-15 | Local | Moderate | 166 | Multiple groups, All | Convenience | Household and community samples | 45.2% (95% CI 37.5-53.1%) | Roche Diagnostics - IgG, IgM, IgA - Elecsys® Anti‐SARS‐CoV‐2 (N) | 1, 0.995 | Validated by manufacturer |
| **Ethiopia - HRP (Addis Ababa)** | Gudina (University of Munich)[20] | 2021-02-11 - 2021-02-26 | Local | Moderate | 151 | Multiple groups, All | Convenience | Household and community samples | 58.2% (95% CI 49.3-65.6%) | Roche Diagnostics - IgG, IgM, IgA - Elecsys® Anti‐SARS‐CoV‐2 (N) | 1, 0.995 | Validated by manufacturer |
| **Ethiopia - HRP (Addis Ababa)** | Gudina (University of Munich)[20] | 2021-02-01 - 2021-03-07 | Local | Moderate | 149 | Multiple groups, All | Convenience | Household and community samples | 45.7% (95% CI 37.5-54%) | Roche Diagnostics - IgG, IgM, IgA - Elecsys® Anti‐SARS‐CoV‐2 (N) | 1, 0.995 | Validated by manufacturer |
| **Ethiopia - HRP (Jimma)** | Gudina (University of Munich)[20] | 2021-01-21 - 2021-02-25 | Local | Moderate | 133 | Multiple groups, All | Convenience | Household and community samples | 26.3% (95% CI 18.4-33.8%) | Roche Diagnostics - IgG, IgM, IgA - Elecsys® Anti‐SARS‐CoV‐2 (N) | 1, 0.995 | Validated by manufacturer |
| **Ethiopia - HRP (Jimma)** | Gudina (University of Munich)[20] | 2021-01-02 - 2021-02-04 | Local | Moderate | 191 | Multiple groups, All | Convenience | Household and community samples | 40.8% (95% CI 33.3-47.6%) | Roche Diagnostics - IgG, IgM, IgA - Elecsys® Anti‐SARS‐CoV‐2 (N) | 1, 0.995 | Validated by manufacturer |
| **Ethiopia - HRP (Addis Ababa)** | Gudina (University of Munich)[20] | 2021-01-02 - 2021-01-22 | Local | Moderate | 218 | Multiple groups, All | Convenience | Household and community samples | 54.2% (95% CI 47.3-60.9%) | Roche Diagnostics - IgG, IgM, IgA - Elecsys® Anti‐SARS‐CoV‐2 (N) | 1, 0.995 | Validated by manufacturer |
| **Ethiopia - HRP (Addis Ababa)** | Gudina (University of Munich)[20] | 2020-12-05 - 2021-02-04 | Local | Moderate | 224 | Multiple groups, All | Convenience | Household and community samples | 39.7% (95% CI 32.8-46%) | Roche Diagnostics - IgG, IgM, IgA - Elecsys® Anti‐SARS‐CoV‐2 (N) | 1, 0.995 | Validated by manufacturer |
| **Ethiopia - HRP (Jimma)** | Gudina (University of Munich)[20] | 2020-12-01 - 2021-02-01 | Local | Moderate | 297 | Multiple groups, All | Convenience | Household and community samples | 32.3% (95% CI 26.7-37.6%) | Roche Diagnostics - IgG, IgM, IgA - Elecsys® Anti‐SARS‐CoV‐2 (N) | 1, 0.995 | Validated by manufacturer |
| **Ethiopia - HRP (Jimma)** | Gudina (University of Munich)[20] | 2020-12-03 - 2021-01-27 | Local | Moderate | 238 | Multiple groups, All | Convenience | Household and community samples | 18% (95% CI 13-23.1%) | Roche Diagnostics - IgG, IgM, IgA - Elecsys® Anti‐SARS‐CoV‐2 (N) | 1, 0.995 | Validated by manufacturer |
| **Ethiopia - HRP (Harari,Oromia)** | Assefa (The London School of Hygiene & Tropical Medicine)[21] | 2020-04-01 - 2021-03-31 | Subnational | Moderate | 1,447 | Adults (18-64 years), Female | Probability | Pregnant or parturient women | 5.7% (95% CI 4.5-7%) | Beijing Wantai Biological - IgG, IgM, IgA - Wantai Rapid Test for Total Antibody to SARS-CoV-2 | 0.947, 0.989 | Validated by manufacturer |
| **Ethiopia - HRP (Addis Ababa)** | Abdella (Ethiopian Public Health Institute)[22] | 2020-07-22 - 2020-09-02 | Local | Low | 956 | Multiple groups, All | Probability | Household and community samples | 4.7% (95% CI 3.5-6.2%) | Core Technology Co. Ltd - IgG, IgM - Coretests COVID-19 IgM/IgG Ab Test | 0.939, 0.981 | Validated by manufacturer |
| **Ethiopia - HRP (Jimma)** | Abdella (Ethiopian Public Health Institute)[22] | 2020-07-22 - 2020-09-02 | Local | Low | 900 | Multiple groups, All | Probability | Household and community samples | 1.2% (95% CI 0.5-2%) | Core Technology Co. Ltd - IgG, IgM - Coretests COVID-19 IgM/IgG Ab Test | 0.939, 0.981 | Validated by manufacturer |
| **Ethiopia - HRP (Dire Dawa city)** | Shaweno (Jimma University Institute of Health)[23] | 2020-06-15 - 2020-07-30 | Local | Moderate | 684 | Multiple groups, All | Probability | Household and community samples | 3.2% (95% CI 1.9-4.7%) | Abbott Laboratories - IgG - Abbott Architect SARS-CoV-2 IgG | 0.99, 1 | Validated by manufacturer |
| **Kenya - HRP (Nairobi)** | Munywoki (Kenya Medical Research Institute)[24] | 2020-11-27 - 2020-12-05 | Local | Moderate | 511 | Multiple groups, All | Probability | Household and community samples | 43.4% (95% CI 38.9-47.7%) | Beijing Wantai Biological - IgG, IgM, IgA - Wantai SARS-CoV-2 Total Ab ELISA | 0.967, 0.975 | Validated by manufacturer |
| **Kenya - HRP (Nairobi)** | Ngere (Washington State University)[25] | 2020-11-02 - 2020-11-23 | Local | Low | 1,164 | Multiple groups, All | Probability | Household and community samples | 32.7% | Beijing Wantai Biological - IgG, IgM, IgA - Wantai SARS-CoV-2 Total Ab ELISA | 0.967, 0.975 | Validated by manufacturer |
| **Mali - HRP (Bancoumana,Doneguebougou,Sotuba)** | Sagara (University of Sciences )[26] | 2020-12-14 - 2021-01-29 | Local | Moderate | 2,353 | Multiple groups, All | Convenience | Household and community samples | 30.8% (95% CI 28.9-32.7%) | NA - IgG - Author designed (ELISA) -Spike | 0.739, 0.994 | Validated by developers |
| **Mali - HRP (Bancoumana,Doneguebougou,Sotuba)** | Sagara (University of Sciences)[26] | 2020-07-28 - 2020-10-16 | Local | Moderate | 2,659 | Multiple groups, All | Convenience | Household and community samples | 6.5% (95% CI 5.6-7.5%) | NA - IgG, IgM - Author designed (ELISA) -Spike | 0.739, 0.994 | Validated by developers |
| **Mozambique - HRP (Massinga)** | Mahomed (Republica de Mocambique Ministerio da Saude)[27] | 2020-11-26 - 2020-12-03 | Local | Moderate | 1,577 | Multiple groups, All | Probability | Household and community samples | 7.4% (95% CI 6.1-8.8%) | Qingdao Hightop Biotech Co., Ltd - IgG, IgM - Hightop COVID-19 IgM/IgG Ab Rapid Test Kit | 0.942, 0.939 | Validated by manufacturer |
| **Mozambique - HRP (Maxixe)** | Mahomed (Republica de Mocambique Ministerio da Saude)[27] | 2020-11-07 - 2020-11-21 | Local | Moderate | 3,974 | Multiple groups, All | Probability | Household and community samples | 5.5% (95% CI 4.8-6.2%) | Qingdao Hightop Biotech Co., Ltd - IgG, IgM - Hightop COVID-19 IgM/IgG Ab Rapid Test Kit | 0.942, 0.939 | Validated by manufacturer |
| **Mozambique - HRP (Chimoio)** | Mahumane (Republica de Mocambique Ministerio da Saude)[28] | 2020-11-02 - 2020-11-17 | Local | Moderate | 9,756 | Multiple groups, All | Probability | Household and community samples | 1.4% (95% CI 1.2-1.6%) | Qingdao Hightop Biotech Co., Ltd - IgG, IgM - Hightop COVID-19 IgM/IgG Ab Rapid Test Kit | 0.942, 0.939 | Validated by manufacturer |
| **Mozambique - HRP (Lichinga)** | Langa (Republica de Mocambique Ministerio da Saude)[29] | 2020-09-28 - 2020-10-09 | Local | Moderate | 1,635 | Multiple groups, All | Probability | Household and community samples | 0.3% (95% CI 0.1-0.6%) | Qingdao Hightop Biotech Co., Ltd - IgG, IgM - Hightop COVID-19 IgM/IgG Ab Rapid Test Kit | 0.942, 0.939 | Validated by manufacturer |
| **Mozambique - HRP (Tete)** | Mahomed (Republica de Mocambique Ministerio da Saude)[30] | 2020-08-31 - 2020-10-12 | Local | Moderate | 1,946 | Multiple groups, All | Probability | Household and community samples | 0.7% (95% CI 0.4-1.1%) | Qingdao Hightop Biotech Co., Ltd - IgG, IgM - Hightop COVID-19 IgM/IgG Ab Rapid Test Kit | 0.942, 0.939 | Validated by manufacturer |
| **Mozambique - HRP (Pemba)** | Arnaldo (República de Moçambique Ministério da Saúde)[31] | 2020-07-06 - 2020-07-13 | Local | Moderate | 1,360 | Multiple groups, All | Probability | Household and community samples | 2.5% (95% CI 1.7-3.5%) | Qingdao Hightop Biotech Co., Ltd - IgG, IgM - Hightop COVID-19 IgM/IgG Ab Rapid Test Kit | 0.942, 0.939 | Validated by manufacturer |
| **Mozambique - HRP (Nampula)** | Gudo (Republica de Mocambique Ministerio da Saude)[32] | 2020-06-17 - 2020-06-30 | Local | Moderate | 1,769 | Multiple groups, All | Probability | Household and community samples | 5% (95% CI 4-6.1%) | SD Biosensor - IgG, IgM - Standard Q COVID-19 IgM/IgG Duo rapid immunochromatography test kit | 0.991, 0.951 | Validated by manufacturer |
| **Nigeria - HRP (Anambra State)** | Okpala (Anambra State Ministry of Health)[33] | 2020-12-08 - 2020-12-15 | Subnational | Moderate | 3,142 | Multiple groups, All | Probability | Household and community samples | 17.6% (95% CI 16.3-18.9%) | Hangzhou Realy Tech Co., Ltd - IgG, IgM - Realy-Tech 2019 nCOV/COVID-19 IgG/IgM Rapid Test | 0.967, 0.937 | Validated by manufacturer |
| **Nigeria - HRP (Enugu State)** | Stafford (UMB); Steinhardt (CDC); Ilori (NCDC); Audu (NIMR) (University of Maryland, Baltimore)[34] | 2020-09-21 - 2020-10-27 | Subnational | Low | 2,147 | Multiple groups, All | Probability | Household and community samples | 25.2% (95% CI 21.7-28.8%) | Abbott Laboratories - IgG - Abbott Architect SARS-CoV-2 IgG;EUROIMMUN AG - IgG - SARS-CoV-2-NCP-IgG ELISA;Luminex Corp - IgG - Multiplexed Particle Flow Cytometry Assay; | N/A | Complex test algorithm: Manual review |
| **Nigeria - HRP (Gombe State)** | Stafford (UMB); Steinhardt (CDC); Ilori (NCDC); Audu (NIMR) (University of Maryland, Baltimore)[34] | 2020-09-21 - 2020-10-27 | Subnational | Low | 3,432 | Multiple groups, All | Probability | Household and community samples | 9.3% (95% CI 6.9-11.6%) | Abbott Laboratories - IgG - Abbott Architect SARS-CoV-2 IgG;EUROIMMUN AG - IgG - SARS-CoV-2-NCP-IgG ELISA;Luminex Corp - IgG - Multiplexed Particle Flow Cytometry Assay | N/A | Complex test algorithm: Manual review |
| **Nigeria - HRP (Nasarawa State)** | Stafford (UMB); Steinhardt (CDC); Ilori (NCDC); Audu (NIMR) (University of Maryland, Baltimore)[34] | 2020-09-21 - 2020-10-27 | Subnational | Low | 2,657 | Multiple groups, All | Probability | Household and community samples | 18% (95% CI 14.3-21.7%) | Abbott Laboratories - IgG - Abbott Architect SARS-CoV-2 IgG;EUROIMMUN AG - IgG - SARS-CoV-2-NCP-IgG ELISA;Luminex Corp - IgG - Multiplexed Particle Flow Cytometry Assay | N/A | Complex test algorithm: Manual review |
| **Nigeria - HRP (Lagos State)** | Stafford (UMB); Steinhardt (CDC); Ilori (NCDC); Audu (NIMR) (University of Maryland, Baltimore)[34] | 2020-09-21 - 2020-10-27 | Subnational | Low | 2,393 | Multiple groups, All | Probability | Household and community samples | 23.2% (95% CI 20-26.5%) | Abbott Laboratories - IgG - Abbott Architect SARS-CoV-2 IgG;EUROIMMUN AG - IgG - SARS-CoV-2-NCP-IgG ELISA;Luminex Corp - IgG - Multiplexed Particle Flow Cytometry Assay | N/A | Complex test algorithm: Manual review |
| **Nigeria - HRP (Niger State)** | Majiya (Ibrahim Badamasi Babangida University)[35] | 2020-06-26 - 2020-06-30 | Subnational | Moderate | 185 | Multiple groups, All | Probability | Household and community samples | 25.4% (95% CI 19.3-32.3%) | Cambridge Network - IgG, IgM - COVID-19 IgG and IgM Rapid Test | 1, 1 | Validated by independent authors/third party/non-developers |
| **South Africa (Agincourt)** | Kleynhans (National Institute for Communicable Diseases)[54] | 2021-09-13 - 2021-09-25 | Local | Moderate | 541 | Multiple groups, All | Probability | Household and community samples | 60.1% (95% CI 55.8-64.2%) | Roche Diagnostics - IgG, IgM, IgA - Elecsys® Anti‐SARS‐CoV‐2 (N) | 1, 0.995 | Validated by manufacturer |
| **South Africa (Klerksdorp)** | Kleynhans (National Institute for Communicable Diseases)[54] | 2021-09-13 - 2021-09-25 | Local | Moderate | 487 | Multiple groups, All | Probability | Household and community samples | 69.6% (95% CI 65.1-73.5%) | Roche Diagnostics - IgG, IgM, IgA - Elecsys® Anti‐SARS‐CoV‐2 (N) | 1, 0.995 | Validated by manufacturer |
| **South Africa (Agincourt)** | Kleynhans (National Institute for Communicable Diseases)[54] | 2021-07-19 - 2021-08-05 | Local | Moderate | 572 | Multiple groups, All | Probability | Household and community samples | 38.5% (95% CI 34.5-42.6%) | Roche Diagnostics - IgG, IgM, IgA - Elecsys® Anti‐SARS‐CoV‐2 (N) | 1, 0.995 | Validated by manufacturer |
| **South Africa (Klerksdorp)** | Kleynhans (National Institute for Communicable Diseases)[54] | 2021-07-19 - 2021-08-05 | Local | Moderate | 494 | Multiple groups, All | Probability | Household and community samples | 55.3% (95% CI 50.8-59.7%) | Roche Diagnostics - IgG, IgM, IgA - Elecsys® Anti‐SARS‐CoV‐2 (N) | 1, 0.995 | Validated by manufacturer |
| **South Africa (Agincourt)** | Kleynhans (National Institute for Communicable Diseases)[54] | 2021-05-20 - 2021-06-09 | Local | Moderate | 573 | Multiple groups, All | Probability | Household and community samples | 25.8% (95% CI 22.3-29.6%) | Roche Diagnostics - IgG, IgM, IgA - Elecsys® Anti‐SARS‐CoV‐2 (N) | 1, 0.995 | Validated by manufacturer |
| **South Africa (Klerksdorp)** | Kleynhans (National Institute for Communicable Diseases)[54] | 2021-05-20 - 2021-06-09 | Local | Moderate | 499 | Multiple groups, All | Probability | Household and community samples | 48.9% (95% CI 44.4-53.4%) | Roche Diagnostics - IgG, IgM, IgA - Elecsys® Anti‐SARS‐CoV‐2 (N) | 1, 0.995 | Validated by manufacturer |
| **South Africa (Agincourt)** | Kleynhans (National Institute for Communicable Diseases)[54] | 2021-03-22 - 2021-04-11 | Local | Moderate | 586 | Multiple groups, All | Probability | Household and community samples | 25.3% (95% CI 21.8-29%) | Roche Diagnostics - IgG, IgM, IgA - Elecsys® Anti‐SARS‐CoV‐2 (N) | 1, 0.995 | Validated by manufacturer |
| **South Africa (Klerksdorp)** | Kleynhans (National Institute for Communicable Diseases)[54] | 2021-03-22 - 2021-04-11 | Local | Moderate | 505 | Multiple groups, All | Probability | Household and community samples | 40.4% (95% CI 36.1-44.8%) | Roche Diagnostics - IgG, IgM, IgA - Elecsys® Anti‐SARS‐CoV‐2 (N) | 1, 0.995 | Validated by manufacturer |
| **South Africa (Agincourt)** | Kleynhans (National Institute for Communicable Diseases)[54] | 2021-01-25 - 2021-02-21 | Local | Moderate | 586 | Multiple groups, All | Probability | Household and community samples | 21.5% (95% CI 18.1-24.9%) | Roche Diagnostics - IgG, IgM, IgA - Elecsys® Anti‐SARS‐CoV‐2 (N) | 1, 0.995 | Validated by manufacturer |
| **South Africa (Klerksdorp)** | Kleynhans (National Institute for Communicable Diseases)[54] | 2021-01-25 - 2021-02-21 | Local | Moderate | 509 | Multiple groups, All | Probability | Household and community samples | 35.4% (95% CI 31-39.5%) | Roche Diagnostics - IgG, IgM, IgA - Elecsys® Anti‐SARS‐CoV‐2 (N) | 1, 0.995 | Validated by manufacturer |
| **South Africa (Agincourt)** | Kleynhans (National Institute for Communicable Diseases)[54] | 2020-11-23 - 2020-12-12 | Local | Moderate | 566 | Multiple groups, All | Probability | Household and community samples | 7.1% (95% CI 5.1-9.5%) | Roche Diagnostics - IgG, IgM, IgA - Elecsys® Anti‐SARS‐CoV‐2 (N) | 1, 0.995 | Validated by manufacturer |
| **South Africa (Klerksdorp)** | Kleynhans (National Institute for Communicable Diseases)[54] | 2020-11-23 - 2020-12-12 | Local | Moderate | 523 | Multiple groups, All | Probability | Household and community samples | 27% (95% CI 23.2-31%) | Roche Diagnostics - IgG, IgM, IgA - Elecsys® Anti‐SARS‐CoV‐2 (N) | 1, 0.995 | Validated by manufacturer |
| **South Africa (Gauteng)** | George (National Health Laboratory Service)[36] | 2020-10-01 - 2020-10-31 | Subnational | Moderate | 1,664 | Multiple groups, All | Convenience | Residual sera | 28.2% | Roche Diagnostics - IgG, IgM, IgA - Elecsys® Anti‐SARS‐CoV‐2 (N) | 1, 0.995 | Validated by manufacturer |
| **South Africa (Agincourt)** | Kleynhans (National Institute for Communicable Diseases)[54] | 2020-09-21 - 2020-10-10 | Local | Moderate | 490 | Multiple groups, All | Probability | Household and community samples | 4.9% (95% CI 3.2-7.2%) | Roche Diagnostics - IgG, IgM, IgA - Elecsys® Anti‐SARS‐CoV‐2 (N) | 1, 0.995 | Validated by manufacturer |
| **South Africa (Klerksdorp)** | Kleynhans (National Institute for Communicable Diseases)[54] | 2020-09-21 - 2020-10-10 | Local | Moderate | 498 | Multiple groups, All | Probability | Household and community samples | 21.7% (95% CI 18.1-25.6%) | Roche Diagnostics - IgG, IgM, IgA - Elecsys® Anti‐SARS‐CoV‐2 (N) | 1, 0.995 | Validated by manufacturer |
| **South Africa (Johannesburg)** | George (National Health Laboratory Service)[37] | 2020-08-01 - 2020-10-31 | Local | Low | 4,393 | Multiple groups, All | Probability | Residual sera | 27% | Roche Diagnostics - IgG, IgM, IgA - Elecsys® Anti‐SARS‐CoV‐2 (N) | 1, 0.995 | Validated by manufacturer |
| **South Africa (Agincourt)** | Kleynhans (National Institute for Communicable Diseases)[54] | 2020-07-20 - 2020-09-17 | Local | Moderate | 439 | Multiple groups, All | Probability | Household and community samples | 1.1% (95% CI 0.4-2.6%) | Roche Diagnostics - IgG, IgM, IgA - Elecsys® Anti‐SARS‐CoV‐2 (N) | 1, 0.995 | Validated by manufacturer |
| **South Africa (Klerksdorp)** | Kleynhans (National Institute for Communicable Diseases)[54] | 2020-07-20 - 2020-09-17 | Local | Moderate | 501 | Multiple groups, All | Probability | Household and community samples | 14.4% (95% CI 11.4-17.8%) | Roche Diagnostics - IgG, IgM, IgA - Elecsys® Anti‐SARS‐CoV‐2 (N) | 1, 0.995 | Validated by manufacturer |
| **South Sudan - HRP (Juba)** | Wiens (World Health Organization)[38] | 2020-08-10 - 2020-09-11 | Local | Low | 2,214 | Multiple groups, All | Probability | Household and community samples | 22.3% (95% CI 20.5-24.1%) | NA - IgG - Author designed (ELISA) -Spike | 0.655, 1 | Validated by developers |
| **Zimbabwe - HRP (Harare)** | Fryatt (Biomedical Research and Training Institute)[39] | 2021-02-10 - 2021-04-17 | Local | Moderate | 1,530 | Multiple groups, All | Probability | Household and community samples | 53% (95% CI 50.4-55.5%) | NA - - Author designed (type unknown) | 0.992, 0.9865 | Validated by developers |
| **Zimbabwe - HRP (Harare)** | Fryatt (Biomedical Research and Training Institute)[39] | 2020-11-20 - 2020-12-20 | Local | Moderate | 620 | Multiple groups, All | Probability | Household and community samples | 19% (95% CI 15.9-22.2%) | NA - - Author designed (type unknown) | 0.992, 0.9865 | Validated by developers |

##### Table S7. Studies used in descriptive analysis (dataset 0: studies rated high risk of bias. For studies rated low and moderate risk of bias, see Tables S5 and S6).

| **Country (Location)** | **Author (Organization)** | **Sampling Dates (YMD)** | **Geographic scope** | **Overall risk of bias** | **Sample size** | **Age, sex** | **Sampling method** | **Sample frame** | **Seroprevalence (95% CI)** | **Test Manufacturers - Isotypes - Names** | **Test Sens, Spec** | **Sens, Spec Source** |
| --- | --- | --- | --- | --- | --- | --- | --- | --- | --- | --- | --- | --- |
| **Angola - HRP (Luanda)** | Sebastiao (Instituto Nacional de Investigação em Saúde)[40] | 2020-09-01 - 2020-09-30 | Local | High | 55 | Adults (18-64 years), All | Convenience | Blood donors | 20% | Biomerieux - IgG, IgM, IgA - VIDAS Multiparametric immunoassay system for medium throughput | 1, 1 | Validated by manufacturer |
| **Ethiopia - HRP (Addis Ababa)** | Kempen (Kuwait University)[41] | 2020-05-18 - 2020-05-21 | Local | High | 99 | Multiple groups, All | Convenience | Residual sera | 3% (95% CI 0.2-7.1%) | Abbott Laboratories - IgG - Abbott Architect SARS-CoV-2 IgG | 0.99, 1 | Validated by manufacturer |
| **Gabon (Libreville)** | Nzoghe (Centre Hospitalier Universitaire (CHU) ‐ Mère‐ Enfant)[42] | 2020-07-15 - 2020-10-15 | Local | High | 1,495 | Multiple groups, All | Convenience | Residual sera | 36.2% (95% CI 33.7-38.7%) | Biomerieux - IgG, IgM, IgA - VIDAS Multiparametric immunoassay system for medium throughput , Roche Diagnostics - IgG, IgM, IgA - Elecsys® Anti‐SARS‐CoV‐2 (N) | 1, 0.995 | Complex algorithm: Manual calculation |
| **Kenya - HRP** | Nyagwange (KEMRI-Wellcome Trust Research Programme)[43] | 2021-08-01 - 2021-08-31 | National | High | 176 | Adults (18-64 years), All | Convenience | Blood donors | 83% (95% CI 76.6-88.2%) | Beijing Wantai Biological - IgG, IgM, IgA - Wantai SARS-CoV-2 Total Ab ELISA | 0.967, 0.975 | Validated by manufacturer |
| **Kenya - HRP (Kilifi)** | Lucinde (KEMRI-Wellcome Trust)[44] | 2020-11-01 - 2020-11-24 | Local | High | 154 | Adults (18-64 years), Female | Sequential | Pregnant or parturient women | 10.4% (95% CI 6.1-16.3%) | NA - IgG - Author designed (ELISA) -Spike | 0.927, 0.99 | Validated by independent authors/third party/non-developers |
| **Kenya - HRP (Kilifi)** | Lucinde (KEMRI-Wellcome Trust)[44] | 2020-10-01 - 2020-10-31 | Local | High | 183 | Adults (18-64 years), Female | Sequential | Pregnant or parturient women | 1.6% (95% CI 0.1-3.9%) | NA - IgG - Author designed (ELISA) -Spike | 0.927, 0.99 | Validated by independent authors/third party/non-developers |
| **Kenya - HRP (Kilifi)** | Lucinde (KEMRI-Wellcome Trust)[44] | 2020-09-18 - 2020-09-30 | Local | High | 82 | Adults (18-64 years), Female | Sequential | Pregnant or parturient women | 0% (95% CI 0-4.4%) | NA - IgG - Author designed (ELISA) -Spike | 0.927, 0.99 | Validated by independent authors/third party/non-developers |
| **Kenya - HRP (Nairobi)** | Lucinde (KEMRI-Wellcome Trust)[44] | 2020-07-30 - 2020-08-25 | Local | High | 196 | Adults (18-64 years), Female | Sequential | Pregnant or parturient women | 46.4% (95% CI 39.3-53.7%) | NA - IgG - Author designed (ELISA) -Spike | 0.927, 0.99 | Validated by independent authors/third party/non-developers |
| **Kenya - HRP** | Nyagwange (KEMRI-Wellcome Trust Research Programme)[43] | 2020-05-01 - 2020-05-31 | National | High | 176 | Adults (18-64 years), All | Convenience | Blood donors | 2.3% (95% CI 0.4-4.9%) | Beijing Wantai Biological - IgG, IgM, IgA - Wantai SARS-CoV-2 Total Ab ELISA | 0.967, 0.975 | Validated by manufacturer |
| **Kenya - HRP (Western Region)** | Crowell (Walter Reed Army Institute of Research)[45] | 2020-01-01 - 2020-03-19 | Subnational | High | 582 | Adults (18-64 years), All | Convenience | Residual sera | 3.3% (95% CI 2-5.1%) | Bio-rad - IgG, IgM, IgA - Platelia SARS-CoV-2 Total Ab assay | 0.993, 0.98 | Validated by manufacturer |
| **Madagascar (Antananarivo)** | Razafimahatratra (Institut Pasteur de Madagascar)[46] | 2021-05-01 - 2021-05-26 | Local | High | 500 | Adults (18-64 years), All | Sequential | Blood donors | 64.8% (95% CI 60.4-69%) | Beijing Wantai Biological - IgG, IgM, IgA - Wantai SARS-CoV-2 Total Ab ELISA | 0.967, 0.975 | Validated by manufacturer |
| **Madagascar (Antananarivo)** | Razafimahatratra (Institut Pasteur de Madagascar)[46] | 2021-04-01 - 2021-04-30 | Local | High | 493 | Adults (18-64 years), All | Sequential | Blood donors | 58.4% (95% CI 53.7-62.6%) | Beijing Wantai Biological - IgG, IgM, IgA - Wantai SARS-CoV-2 Total Ab ELISA | 0.967, 0.975 | Validated by manufacturer |
| **Madagascar (Antananarivo)** | Razafimahatratra (Institut Pasteur de Madagascar)[46] | 2021-03-01 - 2021-03-31 | Local | High | 497 | Adults (18-64 years), All | Sequential | Blood donors | 49.9% (95% CI 45.4-54.4%) | Beijing Wantai Biological - IgG, IgM, IgA - Wantai SARS-CoV-2 Total Ab ELISA | 0.967, 0.975 | Validated by manufacturer |
| **Madagascar (Antananarivo)** | Razafimahatratra (Institut Pasteur de Madagascar)[46] | 2021-02-01 - 2021-02-28 | Local | High | 319 | Adults (18-64 years), All | Sequential | Blood donors | 50.5% (95% CI 44.8-56.1%) | Beijing Wantai Biological - IgG, IgM, IgA - Wantai SARS-CoV-2 Total Ab ELISA | 0.967, 0.975 | Validated by manufacturer |
| **Madagascar (Antananarivo)** | Razafimahatratra (Institut Pasteur de Madagascar)[46] | 2021-01-01 - 2021-01-31 | Local | High | 389 | Adults (18-64 years), All | Sequential | Blood donors | 44.7% (95% CI 39.5-49.6%) | Beijing Wantai Biological - IgG, IgM, IgA - Wantai SARS-CoV-2 Total Ab ELISA | 0.967, 0.975 | Validated by manufacturer |
| **Madagascar (Antananarivo)** | Razafimahatratra (Institut Pasteur de Madagascar)[46] | 2020-12-01 - 2020-12-31 | Local | High | 177 | Adults (18-64 years), All | Sequential | Blood donors | 22% (95% CI 15.7-28.3%) | ID.Vet - IgG - ID Screen | 0.952, 0.998 | Validated by manufacturer |
| **Madagascar (Antananarivo)** | Schoenhals (Institut Pasteur de Madagascar)[47] | 2020-11-01 - 2020-11-30 | Local | High | 502 | Adults (18-64 years), All | Convenience | Blood donors | 31.3% (95% CI 27.2-35.5%) | ID.Vet - IgG - ID Screen | 0.952, 0.998 | Validated by manufacturer |
| **Madagascar (Fianarantsoa)** | Schoenhals (Institut Pasteur de Madagascar)[47] | 2020-11-01 - 2020-11-30 | Local | High | 172 | Adults (18-64 years), All | Convenience | Blood donors | 12.8% (95% CI 8.2-18.7%) | ID.Vet - IgG - ID Screen | 0.952, 0.998 | Validated by manufacturer |
| **Madagascar (Mahajanga)** | Schoenhals (Institut Pasteur de Madagascar)[47] | 2020-11-01 - 2020-11-30 | Local | High | 30 | Adults (18-64 years), All | Convenience | Blood donors | 16.7% (95% CI 5.6-34.7%) | ID.Vet - IgG - ID Screen | 0.952, 0.998 | Validated by manufacturer |
| **Madagascar (Toamasina)** | Schoenhals (Institut Pasteur de Madagascar)[47] | 2020-11-01 - 2020-11-30 | Local | High | 282 | Adults (18-64 years), All | Convenience | Blood donors | 18.4% (95% CI 13.8-23.1%) | ID.Vet - IgG - ID Screen | 0.952, 0.998 | Validated by manufacturer |
| **Madagascar (Toliara)** | Schoenhals (Institut Pasteur de Madagascar)[47] | 2020-11-01 - 2020-11-30 | Local | High | 199 | Adults (18-64 years), All | Convenience | Blood donors | 18.1% (95% CI 13-24.2%) | ID.Vet - IgG - ID Screen | 0.952, 0.998 | Validated by manufacturer |
| **Madagascar (Antananarivo)** | Razafimahatratra (Institut Pasteur de Madagascar)[46] | 2020-11-01 - 2020-11-30 | Local | High | 503 | Adults (18-64 years), All | Sequential | Blood donors | 31.2% (95% CI 27-35.3%) | ID.Vet - IgG - ID Screen | 0.952, 0.998 | Validated by manufacturer |
| **Madagascar (Antananarivo)** | Schoenhals (Institut Pasteur de Madagascar)[47] | 2020-10-01 - 2020-10-31 | Local | High | 502 | Adults (18-64 years), All | Convenience | Blood donors | 36% (95% CI 31.7-40.2%) | ID.Vet - IgG - ID Screen | 0.952, 0.998 | Validated by manufacturer |
| **Madagascar (Fianarantsoa)** | Schoenhals (Institut Pasteur de Madagascar)[47] | 2020-10-01 - 2020-10-31 | Local | High | 152 | Adults (18-64 years), All | Convenience | Blood donors | 15.1% (95% CI 9.3-21.1%) | ID.Vet - IgG - ID Screen | 0.952, 0.998 | Validated by manufacturer |
| **Madagascar (Mahajanga)** | Schoenhals (Institut Pasteur de Madagascar)[47] | 2020-10-01 - 2020-10-31 | Local | High | 26 | Adults (18-64 years), All | Convenience | Blood donors | 26.9% (95% CI 9-43.6%) | ID.Vet - IgG - ID Screen | 0.952, 0.998 | Validated by manufacturer |
| **Madagascar (Toamasina)** | Schoenhals (Institut Pasteur de Madagascar)[47] | 2020-10-01 - 2020-10-31 | Local | High | 199 | Adults (18-64 years), All | Convenience | Blood donors | 26.1% (95% CI 19.7-32.3%) | ID.Vet - IgG - ID Screen | 0.952, 0.998 | Validated by manufacturer |
| **Madagascar (Toliara)** | Schoenhals (Institut Pasteur de Madagascar)[47] | 2020-10-01 - 2020-10-31 | Local | High | 139 | Adults (18-64 years), All | Convenience | Blood donors | 22.3% (95% CI 15.1-29.4%) | ID.Vet - IgG - ID Screen | 0.952, 0.998 | Validated by manufacturer |
| **Madagascar (Antananarivo)** | Razafimahatratra (Institut Pasteur de Madagascar)[46] | 2020-10-01 - 2020-10-31 | Local | High | 497 | Adults (18-64 years), All | Sequential | Blood donors | 35.8% (95% CI 31.4-40%) | ID.Vet - IgG - ID Screen | 0.952, 0.998 | Validated by manufacturer |
| **Madagascar (Antananarivo)** | Schoenhals (Institut Pasteur de Madagascar)[47] | 2020-09-01 - 2020-09-30 | Local | High | 481 | Adults (18-64 years), All | Convenience | Blood donors | 38.5% (95% CI 34.1-43%) | ID.Vet - IgG - ID Screen | 0.952, 0.998 | Validated by manufacturer |
| **Madagascar (Fianarantsoa)** | Schoenhals (Institut Pasteur de Madagascar)[47] | 2020-09-01 - 2020-09-30 | Local | High | 173 | Adults (18-64 years), All | Convenience | Blood donors | 18.5% (95% CI 13-25.1%) | ID.Vet - IgG - ID Screen | 0.952, 0.998 | Validated by manufacturer |
| **Madagascar (Mahajanga)** | Schoenhals (Institut Pasteur de Madagascar)[47] | 2020-09-01 - 2020-09-30 | Local | High | 53 | Adults (18-64 years), All | Convenience | Blood donors | 37.7% (95% CI 23.1-50.2%) | ID.Vet - IgG - ID Screen | 0.952, 0.998 | Validated by manufacturer |
| **Madagascar (Toamasina)** | Schoenhals (Institut Pasteur de Madagascar)[47] | 2020-09-01 - 2020-09-30 | Local | High | 213 | Adults (18-64 years), All | Convenience | Blood donors | 29.1% (95% CI 22.7-35.2%) | ID.Vet - IgG - ID Screen | 0.952, 0.998 | Validated by manufacturer |
| **Madagascar (Toliara)** | Schoenhals (Institut Pasteur de Madagascar)[47] | 2020-09-01 - 2020-09-30 | Local | High | 185 | Adults (18-64 years), All | Convenience | Blood donors | 20.5% (95% CI 14.5-26.5%) | ID.Vet - IgG - ID Screen | 0.952, 0.998 | Validated by manufacturer |
| **Madagascar (Fianarantsoa)** | Schoenhals (Institut Pasteur de Madagascar)[47] | 2020-08-01 - 2020-08-31 | Local | High | 120 | Adults (18-64 years), All | Convenience | Blood donors | 9.2% (95% CI 4.7-15.8%) | ID.Vet - IgG - ID Screen | 0.952, 0.998 | Validated by manufacturer |
| **Madagascar (Mahajanga)** | Schoenhals (Institut Pasteur de Madagascar)[47] | 2020-08-01 - 2020-08-31 | Local | High | 107 | Adults (18-64 years), All | Convenience | Blood donors | 35.5% (95% CI 25.6-44.4%) | ID.Vet - IgG - ID Screen | 0.952, 0.998 | Validated by manufacturer |
| **Madagascar (Toamasina)** | Schoenhals (Institut Pasteur de Madagascar)[47] | 2020-08-01 - 2020-08-31 | Local | High | 227 | Adults (18-64 years), All | Convenience | Blood donors | 40.1% (95% CI 33.7-46.8%) | ID.Vet - IgG - ID Screen | 0.952, 0.998 | Validated by manufacturer |
| **Madagascar (Toliara)** | Schoenhals (Institut Pasteur de Madagascar)[47] | 2020-08-01 - 2020-08-31 | Local | High | 167 | Adults (18-64 years), All | Convenience | Blood donors | 7.2% (95% CI 3.8-12.2%) | ID.Vet - IgG - ID Screen | 0.952, 0.998 | Validated by manufacturer |
| **Madagascar (Antananarivo)** | Schoenhals (Institut Pasteur de Madagascar)[47] | 2020-08-01 - 2020-08-30 | Local | High | 432 | Adults (18-64 years), All | Convenience | Blood donors | 35.2% (95% CI 30.7-39.9%) | ID.Vet - IgG - ID Screen | 0.952, 0.998 | Validated by manufacturer |
| **Madagascar (Antananarivo)** | Schoenhals (Institut Pasteur de Madagascar)[47] | 2020-07-01 - 2020-07-31 | Local | High | 428 | Adults (18-64 years), All | Convenience | Blood donors | 29% (95% CI 24.7-33.5%) | ID.Vet - IgG - ID Screen | 0.952, 0.998 | Validated by manufacturer |
| **Madagascar (Mahajanga)** | Schoenhals (Institut Pasteur de Madagascar)[47] | 2020-07-01 - 2020-07-31 | Local | High | 98 | Adults (18-64 years), All | Convenience | Blood donors | 1% (95% CI 0-3.7%) | ID.Vet - IgG - ID Screen | 0.952, 0.998 | Validated by manufacturer |
| **Madagascar (Toamasina)** | Schoenhals (Institut Pasteur de Madagascar)[47] | 2020-07-01 - 2020-07-31 | Local | High | 182 | Adults (18-64 years), All | Convenience | Blood donors | 43.4% (95% CI 35.6-50.4%) | ID.Vet - IgG - ID Screen | 0.952, 0.998 | Validated by manufacturer |
| **Madagascar (Toliara)** | Schoenhals (Institut Pasteur de Madagascar)[47] | 2020-07-01 - 2020-07-31 | Local | High | 147 | Adults (18-64 years), All | Convenience | Blood donors | 0.7% (95% CI 0-3.7%) | ID.Vet - IgG - ID Screen | 0.952, 0.998 | Validated by manufacturer |
| **Madagascar (Fianarantsoa)** | Schoenhals (Institut Pasteur de Madagascar)[47] | 2020-07-01 - 2020-07-30 | Local | High | 142 | Adults (18-64 years), All | Convenience | Blood donors | 0% (95% CI 0-2.6%) | ID.Vet - IgG - ID Screen | 0.952, 0.998 | Validated by manufacturer |
| **Madagascar (Antananarivo)** | Schoenhals (Institut Pasteur de Madagascar)[47] | 2020-06-01 - 2020-06-30 | Local | High | 308 | Adults (18-64 years), All | Convenience | Blood donors | 3.3% (95% CI 1.6-5.9%) | ID.Vet - IgG - ID Screen | 0.952, 0.998 | Validated by manufacturer |
| **Madagascar (Fianarantsoa)** | Schoenhals (Institut Pasteur de Madagascar)[47] | 2020-06-01 - 2020-06-30 | Local | High | 129 | Adults (18-64 years), All | Convenience | Blood donors | 0.8% (95% CI 0-4.2%) | ID.Vet - IgG - ID Screen | 0.952, 0.998 | Validated by manufacturer |
| **Madagascar (Mahajanga)** | Schoenhals (Institut Pasteur de Madagascar)[47] | 2020-06-01 - 2020-06-30 | Local | High | 50 | Adults (18-64 years), All | Convenience | Blood donors | 0% (95% CI 0-7.1%) | ID.Vet - IgG - ID Screen | 0.952, 0.998 | Validated by manufacturer |
| **Madagascar (Toamasina)** | Schoenhals (Institut Pasteur de Madagascar)[47] | 2020-06-01 - 2020-06-30 | Local | High | 82 | Adults (18-64 years), All | Convenience | Blood donors | 39% (95% CI 27.3-49.2%) | ID.Vet - IgG - ID Screen | 0.952, 0.998 | Validated by manufacturer |
| **Madagascar (Toliara)** | Schoenhals (Institut Pasteur de Madagascar)[47] | 2020-06-01 - 2020-06-30 | Local | High | 87 | Adults (18-64 years), All | Convenience | Blood donors | 1.2% (95% CI 0-6.2%) | ID.Vet - IgG - ID Screen | 0.952, 0.998 | Validated by manufacturer |
| **Madagascar (Antananarivo)** | Schoenhals (Institut Pasteur de Madagascar)[47] | 2020-05-01 - 2020-05-31 | Local | High | 598 | Adults (18-64 years), All | Convenience | Blood donors | 0.2% (95% CI 0-0.9%) | ID.Vet - IgG - ID Screen | 0.952, 0.998 | Validated by manufacturer |
| **Madagascar (Antananarivo)** | Schoenhals (Institut Pasteur de Madagascar)[47] | 2020-04-01 - 2020-04-30 | Local | High | 157 | Adults (18-64 years), All | Convenience | Blood donors | 0% (95% CI 0-2.3%) | ID.Vet - IgG - ID Screen | 0.952, 0.998 | Validated by manufacturer |
| **Madagascar (Antananarivo)** | Schoenhals (Institut Pasteur de Madagascar)[47] | 2020-03-01 - 2020-03-30 | Local | High | 496 | Adults (18-64 years), All | Convenience | Blood donors | 0.8% (95% CI 0.1-1.8%) | ID.Vet - IgG - ID Screen | 0.952, 0.998 | Validated by manufacturer |
| **Nigeria - HRP (Enugu,Abuja)** | Ifeorah (Molecular Pathology Institute)[48] | 2020-08-01 - 2020-08-31 | Local | High | 113 | Adults (18-64 years), All | Convenience | Blood donors | 42% (95% CI 32.4-51.2%) | NovaTec Immundiagnostics GmbH - IgG - NovaLisa® SARS-CoV-2 IgG | 1, 0.98 | Validated by manufacturer |
| **South Africa (Johannesburg)** | Fairlie (Wits Reproductive Health and HIV Institute)[49] | 2021-03-17 - 2021-06-09 | Local | High | 500 | Multiple groups, All | Convenience | Pregnant or parturient women | 64% (95% CI 59.6-68.2%) | Beijing Wantai Biological - IgG, IgM, IgA - Wantai SARS-CoV-2 Total Ab ELISA;Roche Diagnostics, Basel, Switzerland - IgG, IgM, IgA - Elecsys® Anti‐SARS‐CoV‐2 | 1, 0.97 | Complex test algorithm: Manual calculation |
| **South Africa (Cape Town)** | Hsiao ( University of Cape Town)[50] | 2020-06-15 - 2020-08-07 | Local | High | 2,791 | Adults (18-64 years), All | Sequential | Residual sera | 40.2% (95% CI 38.3-42%) | Roche Diagnostics - IgG, IgM, IgA - Elecsys® Anti‐SARS‐CoV‐2 (N) | 1, 0.995 | Validated by manufacturer |
| **Egypt - HRP (Cairo)** | Abdelmaksoud (Ain-Shams University)[51] | 2020-09-06 - 2020-10-31 | Local | High | 100 | Adults (18-64 years), All | Probability | Blood donors | 38% (95% CI 28.5-48.3%) | AMEDA Labordiagnostik GmbH - IgG, IgM - AMP Rapid Test SARS-CoV-2 IgG/IgM | 1, 0.975 | Validated by manufacturer |
| **Egypt - HRP (Cairo)** | Girgis (Ain-Shams University)[52] | 2020-05-05 - 2020-10-31 | Local | High | 2,927 | Multiple groups, All | Sequential | Residual sera | 30% (95% CI 28.3-31.6%) | Roche Diagnostics - IgG, IgM, IgA - Elecsys® Anti‐SARS‐CoV‐2 (N) | 1, 0.995 | Validated by manufacturer |

#### S4.3 Risk of bias ratings for all studies

| **Country** | **Author (Organization)** | **Item 1: Appropriate sample frame** | **Item 2: Probability sampling method** | **Item 3: Adequate sample size** | **Item 4: Subjects & setting described** | **Item 5: Good coverage of sample** | **Item 6: Sens>=90%, Spec>=97%** | **Item 7: Same test for all subjects** | **Item 8: Appropriate statistical analysis** | **Item 9: Adequate response rate** | **Overall risk of bias** |
| --- | --- | --- | --- | --- | --- | --- | --- | --- | --- | --- | --- |
| **Burkina Faso** | Dr Isidore Traore (National Institute of Public Health)[15] | Yes | Yes | Yes | Yes | Yes | Yes | Yes | No | Yes | Moderate |
| **Cameroon** | Kene Nwosu (Hopital Central de Yaounde)[16] | Yes | Yes | Yes | Yes | Yes | No | Yes | Yes | Yes | Low |
| **Cameroon** | Tiffany Harris (ICAP at Columbia University)[1] | Yes | No | Yes | Yes | Yes | Yes | Yes | Yes | Yes | Moderate |
| **Central African Republic** | Alexandre Manirakiza (Institut Pasteur of Bangui)[53] | Yes | Yes | Yes | Yes | Yes | Yes | Yes | No | Unclear | Moderate |
| **Democratic Republic of the Congo** | Antoine N Nkuba (Institut National de Recherche Biomédicale)[19] | Yes | Yes | Yes | Yes | Yes | Unclear | Yes | Yes | Yes | Low |
| **Democratic Republic of the Congo** | Sheila Makiala Mandanda (Institut National de Recherche Biomédicale)[18] | Yes | Yes | Yes | Yes | Unclear | Yes | Yes | No | Yes | Moderate |
| **Ethiopia** | John Kempen (Kuwait University)[41] | No | No | No | Yes | No | Yes | Yes | No | Unclear | High |
| **Ethiopia** | Enyew Birru Tadesse (Federal Ministry of Health)[2] | Yes | Yes | Yes | Yes | Yes | Yes | Yes | Yes | Unclear | Low |
| **Ethiopia** | Tamrat Shaweno (Jimma University Institute of Health)[23] | Yes | Yes | Yes | Yes | Yes | Yes | Yes | No | Yes | Moderate |
| **Ethiopia** | Saro Abdella (Ethiopian Public Health Institute)[22] | Yes | Yes | Yes | Yes | Yes | Yes | Yes | Yes | Unclear | Low |
| **Ethiopia** | Saro Abdella (Ethiopian Public Health Institute)[22] | Yes | Yes | Yes | Yes | Yes | Yes | Yes | Yes | Unclear | Low |
| **Ethiopia** | Nega Assefa (The London School of Hygiene & Tropical Medicine)[21] | No | Yes | Yes | No | Unclear | Yes | Yes | No | Unclear | Moderate |
| **Gabon** | Amandine Mveang Nzoghe (Centre Hospitalier Universitaire (CHU) ‐ Mère‐ Enfant)[42] | No | No | Yes | No | Unclear | Yes | Yes | No | Unclear | High |
| **Ghana** | Irene Owusu Donkor (Noguchi Memorial Institute for Medical Rsearch)[3] | Yes | Yes | Yes | Yes | Yes | Yes | Yes | No | Yes | Moderate |
| **Ghana** | Irene Owusu Donkor (Noguchi Memorial Institute for Medical Rsearch)[3] | Yes | Yes | Yes | Yes | Yes | Yes | Yes | No | Yes | Moderate |
| **Ghana** | Irene Owusu Donkor (Noguchi Memorial Institute for Medical Rsearch)[3] | Yes | Yes | Yes | Yes | Yes | Yes | Yes | No | Yes | Moderate |
| **Kenya** | Trevor Crowell (Walter Reed Army Institute of Research)[45] | No | No | No | Yes | Unclear | Yes | Yes | No | N/A | High |
| **Kenya** | James Nyagwange (KEMRI-Wellcome Trust Research Programme)[43] | No | No | No | No | No | Yes | Yes | No | N/A |  |
| **Kenya** | R Lucinde (KEMRI-Wellcome Trust)[44] | No | No | No | No | Unclear | Unclear | Yes | No | Unclear | High |
| **Kenya** | R Lucinde (KEMRI-Wellcome Trust)[44] | No | No | No | No | Unclear | Unclear | Yes | No | Unclear | High |
| **Kenya** | Ifedayo M. O. Adetifa (KEMRI-Wellcome Trust Research Programme)[5] | Yes | No | Yes | Yes | No | Unclear | Yes | Yes | N/A | Moderate |
| **Kenya** | R Lucinde (KEMRI-Wellcome Trust)[44] | No | No | No | No | Unclear | Unclear | Yes | No | Unclear | High |
| **Kenya** | Isaac Ngere (Washington State University)[25] | Yes | Yes | Yes | Yes | Yes | Yes | Yes | Yes | Yes | Low |
| **Kenya** | R Lucinde (KEMRI-Wellcome Trust)[44] | No | No | No | No | Unclear | Unclear | Yes | No | Unclear | High |
| **Kenya** | Patrick Munywoki (Kenya Medical Research Institute)[24] | Yes | Yes | No | No | Yes | Yes | Yes | Yes | Yes | Moderate |
| **Kenya** | Sophie Uyoga (KEMRI-Wellcome Trust Research Programme)[4] | No | No | Yes | Yes | No | Unclear | Yes | Yes | N/A | Moderate |
| **Kenya** | James Nyagwange (KEMRI-Wellcome Trust Research Programme)[43] | No | No | No | No | No | Yes | Yes | No | N/A |  |
| **Madagascar** | Matthieu Schoenhals (Institut Pasteur de Madagascar)[47] | No | No | No | No | No | Yes | Yes | Yes | Unclear | High |
| **Madagascar** | Matthieu Schoenhals (Institut Pasteur de Madagascar)[47] | No | No | No | No | No | Yes | Yes | Yes | Unclear | High |
| **Madagascar** | Matthieu Schoenhals (Institut Pasteur de Madagascar)[47] | No | No | No | No | No | Yes | Yes | Yes | Unclear | High |
| **Madagascar** | Matthieu Schoenhals (Institut Pasteur de Madagascar)[47] | No | No | No | No | No | Yes | Yes | Yes | Unclear | High |
| **Madagascar** | Matthieu Schoenhals (Institut Pasteur de Madagascar)[47] | No | No | No | No | No | Yes | Yes | Yes | Unclear | High |
| **Madagascar** | Matthieu Schoenhals (Institut Pasteur de Madagascar)[47] | No | No | No | No | No | Yes | Yes | Yes | Unclear | High |
| **Madagascar** | Matthieu Schoenhals (Institut Pasteur de Madagascar)[47] | No | No | No | No | No | Yes | Yes | Yes | Unclear | High |
| **Madagascar** | Matthieu Schoenhals (Institut Pasteur de Madagascar)[47] | No | No | No | No | No | Yes | Yes | Yes | Unclear | High |
| **Madagascar** | Matthieu Schoenhals (Institut Pasteur de Madagascar)[47] | No | No | No | No | No | Yes | Yes | Yes | Unclear | High |
| **Madagascar** | Matthieu Schoenhals (Institut Pasteur de Madagascar)[47] | No | No | No | No | No | Yes | Yes | Yes | Unclear | High |
| **Madagascar** | Matthieu Schoenhals (Institut Pasteur de Madagascar)[47] | No | No | No | No | No | Yes | Yes | Yes | Unclear | High |
| **Madagascar** | Matthieu Schoenhals (Institut Pasteur de Madagascar)[47] | No | No | No | No | No | Yes | Yes | Yes | Unclear | High |
| **Madagascar** | Matthieu Schoenhals (Institut Pasteur de Madagascar)[47] | No | No | No | No | No | Yes | Yes | Yes | Unclear | High |
| **Madagascar** | Matthieu Schoenhals (Institut Pasteur de Madagascar)[47] | No | No | No | No | No | Yes | Yes | Yes | Unclear | High |
| **Madagascar** | Matthieu Schoenhals (Institut Pasteur de Madagascar)[47] | No | No | No | No | No | Yes | Yes | Yes | Unclear | High |
| **Madagascar** | Matthieu Schoenhals (Institut Pasteur de Madagascar)[47] | No | No | No | No | No | Yes | Yes | Yes | Unclear | High |
| **Madagascar** | Matthieu Schoenhals (Institut Pasteur de Madagascar)[47] | No | No | No | No | No | Yes | Yes | Yes | Unclear | High |
| **Madagascar** | Matthieu Schoenhals (Institut Pasteur de Madagascar)[47] | No | No | No | No | No | Yes | Yes | Yes | Unclear | High |
| **Madagascar** | Matthieu Schoenhals (Institut Pasteur de Madagascar)[47] | No | No | No | No | No | Yes | Yes | Yes | Unclear | High |
| **Madagascar** | Matthieu Schoenhals (Institut Pasteur de Madagascar)[47] | No | No | No | No | No | Yes | Yes | Yes | Unclear | High |
| **Madagascar** | Matthieu Schoenhals (Institut Pasteur de Madagascar)[47] | No | No | No | No | No | Yes | Yes | Yes | Unclear | High |
| **Madagascar** | Matthieu Schoenhals (Institut Pasteur de Madagascar)[47] | No | No | No | No | No | Yes | Yes | Yes | Unclear | High |
| **Madagascar** | Matthieu Schoenhals (Institut Pasteur de Madagascar)[47] | No | No | No | No | No | Yes | Yes | Yes | Unclear | High |
| **Madagascar** | Matthieu Schoenhals (Institut Pasteur de Madagascar)[47] | No | No | No | No | No | Yes | Yes | Yes | Unclear | High |
| **Madagascar** | Matthieu Schoenhals (Institut Pasteur de Madagascar)[47] | No | No | No | No | No | Yes | Yes | Yes | Unclear | High |
| **Madagascar** | Matthieu Schoenhals (Institut Pasteur de Madagascar)[47] | No | No | No | No | No | Yes | Yes | Yes | Unclear | High |
| **Madagascar** | Matthieu Schoenhals (Institut Pasteur de Madagascar)[47] | No | No | No | No | No | Yes | Yes | Yes | Unclear | High |
| **Madagascar** | Matthieu Schoenhals (Institut Pasteur de Madagascar)[47] | No | No | No | No | No | Yes | Yes | Yes | Unclear | High |
| **Madagascar** | Solohery Razafimahatratra (Institut Pasteur de Madagascar)[46] | No | No | No | Yes | Unclear | Yes | Yes | No | N/A | High |
| **Madagascar** | Matthieu Schoenhals (Institut Pasteur de Madagascar)[47] | No | No | No | No | No | Yes | Yes | Yes | Unclear | High |
| **Madagascar** | Matthieu Schoenhals (Institut Pasteur de Madagascar)[47] | No | No | No | No | No | Yes | Yes | Yes | Unclear | High |
| **Madagascar** | Matthieu Schoenhals (Institut Pasteur de Madagascar)[47] | No | No | No | No | No | Yes | Yes | Yes | Unclear | High |
| **Madagascar** | Matthieu Schoenhals (Institut Pasteur de Madagascar)[47] | No | No | No | No | No | Yes | Yes | Yes | Unclear | High |
| **Madagascar** | Matthieu Schoenhals (Institut Pasteur de Madagascar)[47] | No | No | No | No | No | Yes | Yes | Yes | Unclear | High |
| **Madagascar** | Solohery Razafimahatratra (Institut Pasteur de Madagascar)[46] | No | No | No | Yes | Unclear | Yes | Yes | No | N/A | High |
| **Madagascar** | Solohery Razafimahatratra (Institut Pasteur de Madagascar)[46] | No | No | No | Yes | Unclear | Yes | Yes | No | N/A | High |
| **Madagascar** | Solohery Razafimahatratra (Institut Pasteur de Madagascar)[46] | No | No | No | Yes | Unclear | Yes | Yes | No | N/A | High |
| **Madagascar** | Solohery Razafimahatratra (Institut Pasteur de Madagascar)[46] | No | No | No | Yes | Unclear | Yes | Yes | No | N/A | High |
| **Madagascar** | Solohery Razafimahatratra (Institut Pasteur de Madagascar)[46] | No | No | No | Yes | Unclear | Yes | Yes | No | N/A | High |
| **Madagascar** | Solohery Razafimahatratra (Institut Pasteur de Madagascar)[46] | No | No | No | Yes | Unclear | Yes | Yes | No | N/A | High |
| **Madagascar** | Solohery Razafimahatratra (Institut Pasteur de Madagascar)[46] | No | No | No | Yes | Unclear | Yes | Yes | No | N/A | High |
| **Malawi** | Jambo (Malawi-Liverpool-Wellcome Trust Clinical Research programme, Liverpool School of Tropical Medicine)[6] | No | Yes | Yes | Yes | No | Yes | Yes | Yes | N/A | Low |
| **Malawi** | Jambo (Malawi-Liverpool-Wellcome Trust Clinical Research programme, Liverpool School of Tropical Medicine)[6] | No | Yes | Yes | Yes | No | Yes | Yes | Yes | N/A | Low |
| **Malawi** | Jambo (Malawi-Liverpool-Wellcome Trust Clinical Research programme, Liverpool School of Tropical Medicine)[6] | No | Yes | Yes | Yes | No | Yes | Yes | Yes | N/A | Low |
| **Malawi** | Jambo (Malawi-Liverpool-Wellcome Trust Clinical Research programme, Liverpool School of Tropical Medicine)[6] | No | Yes | Yes | Yes | No | Yes | Yes | Yes | N/A | Low |
| **Malawi** | Jambo (Malawi-Liverpool-Wellcome Trust Clinical Research programme, Liverpool School of Tropical Medicine)[6] | No | Yes | Yes | Yes | No | Yes | Yes | Yes | N/A | Low |
| **Malawi** | Jambo (Malawi-Liverpool-Wellcome Trust Clinical Research programme, Liverpool School of Tropical Medicine)[6] | No | Yes | Yes | Yes | No | Yes | Yes | Yes | N/A | Low |
| **Malawi** | Jambo (Malawi-Liverpool-Wellcome Trust Clinical Research programme, Liverpool School of Tropical Medicine)[6] | No | Yes | Yes | Yes | No | Yes | Yes | Yes | N/A | Low |
| **Malawi** | Jambo (Malawi-Liverpool-Wellcome Trust Clinical Research programme, Liverpool School of Tropical Medicine)[6] | No | Yes | Yes | Yes | No | Yes | Yes | Yes | N/A | Low |
| **Malawi** | Jambo (Malawi-Liverpool-Wellcome Trust Clinical Research programme, Liverpool School of Tropical Medicine)[6] | No | Yes | Yes | Yes | No | Yes | Yes | Yes | N/A | Low |
| **Malawi** | Jambo (Malawi-Liverpool-Wellcome Trust Clinical Research programme, Liverpool School of Tropical Medicine)[6] | No | Yes | Yes | Yes | No | Yes | Yes | Yes | N/A | Low |
| **Malawi** | Jambo (Malawi-Liverpool-Wellcome Trust Clinical Research programme, Liverpool School of Tropical Medicine)[6] | No | Yes | Yes | Yes | No | Yes | Yes | Yes | N/A | Low |
| **Malawi** | Jambo (Malawi-Liverpool-Wellcome Trust Clinical Research programme, Liverpool School of Tropical Medicine)[6] | No | Yes | Yes | Yes | No | Yes | Yes | Yes | N/A | Low |
| **Malawi** | Jambo (Malawi-Liverpool-Wellcome Trust Clinical Research programme, Liverpool School of Tropical Medicine)[6] | No | Yes | Yes | Yes | No | Yes | Yes | Yes | N/A | Low |
| **Malawi** | Jambo (Malawi-Liverpool-Wellcome Trust Clinical Research programme, Liverpool School of Tropical Medicine)[6] | No | Yes | Yes | Yes | No | Yes | Yes | Yes | N/A | Low |
| **Malawi** | Jambo (Malawi-Liverpool-Wellcome Trust Clinical Research programme, Liverpool School of Tropical Medicine)[6] | No | Yes | Yes | Yes | No | Yes | Yes | Yes | N/A | Low |
| **Malawi** | Jambo (Malawi-Liverpool-Wellcome Trust Clinical Research programme, Liverpool School of Tropical Medicine)[6] | No | Yes | Yes | Yes | No | Yes | Yes | Yes | N/A | Low |
| **Malawi** | Jambo (Malawi-Liverpool-Wellcome Trust Clinical Research programme, Liverpool School of Tropical Medicine)[6] | No | Yes | Yes | Yes | No | Yes | Yes | Yes | N/A | Low |
| **Malawi** | Jambo (Malawi-Liverpool-Wellcome Trust Clinical Research programme, Liverpool School of Tropical Medicine)[6] | No | Yes | Yes | Yes | No | Yes | Yes | Yes | N/A | Low |
| **Malawi** | Jambo (Malawi-Liverpool-Wellcome Trust Clinical Research programme, Liverpool School of Tropical Medicine)[6] | No | Yes | Yes | Yes | No | Yes | Yes | Yes | N/A | Low |
| **Malawi** | Matthews Kagoli/Joe Theu (Public Health Institute of Malawi(PHIM) MOH)[7] | Yes | Yes | Yes | Yes | Yes | Yes | Yes | Yes | Yes | Low |
| **Mali** | Issaka Sagara (University of Sciences)[26] | Yes | No | Yes | Yes | Yes | Unclear | Yes | Yes | Yes | Moderate |
| **Mali** | Issaka Sagara (University of Sciences )[26] | Yes | No | Yes | Yes | Yes | Unclear | Yes | Yes | Yes | Moderate |
| **Mozambique** | Eduardo Samo Gudo (Republica de Mocambique Ministerio da Saude)[32] | Yes | Yes | Yes | No | Unclear | No | Yes | No | Unclear | Moderate |
| **Mozambique** | Paulo Arnaldo (República de Moçambique Ministério da Saúde)[31] | Yes | Yes | Yes | Yes | Yes | No | Yes | No | No | Moderate |
| **Mozambique** | Mussagy Mahomed (Republica de Mocambique Ministerio da Saude)[30] | Yes | Yes | Yes | Yes | Unclear | No | Yes | No | Unclear | Moderate |
| **Mozambique** | Jeronimo Langa (Republica de Mocambique Ministerio da Saude)[29] | Yes | Yes | Yes | Yes | Unclear | No | Yes | No | Unclear | Moderate |
| **Mozambique** | Arlete Mahumane (Republica de Mocambique Ministerio da Saude)[28] | Yes | Yes | Yes | Yes | Yes | No | Yes | No | Unclear | Moderate |
| **Mozambique** | Mussagy Mahomed (Republica de Mocambique Ministerio da Saude)[27] | Yes | Yes | Yes | Yes | Unclear | No | Yes | No | Unclear | Moderate |
| **Mozambique** | Mussagy Mahomed (Republica de Mocambique Ministerio da Saude)[27] | Yes | Yes | Yes | Yes | Unclear | No | Yes | No | Unclear | Moderate |
| **Nigeria** | Hussaini Majiya (Ibrahim Badamasi Babangida University)[35] | Yes | Yes | No | No | Unclear | Unclear | Yes | No | Unclear | Moderate |
| **Nigeria** | Ijeoma Ifeorah (Molecular Pathology Institute)[48] | No | No | Yes | Yes | Unclear | Yes | Yes | No | N/A | High |
| **Nigeria** | OgoChukwu Vincent Okpala (Anambra State Ministry of Health)[33] | Yes | Yes | Yes | Yes | No | No | Yes | No | N/A | Moderate |
| **Nigeria** | Kristen A. Stafford (UMB); Laura Steinhardt (CDC); Elsie Ilori (NCDC); Rosemary Audu (NIMR) (University of Maryland, Baltimore)[34] | Yes | Yes | Yes | Yes | Yes | Yes | Yes | Yes | Yes | Low |
| **Nigeria** | Kristen A. Stafford (UMB); Laura Steinhardt (CDC); Elsie Ilori (NCDC); Rosemary Audu (NIMR) (University of Maryland, Baltimore)[34] | Yes | Yes | Yes | Yes | Yes | Yes | Yes | Yes | Yes | Low |
| **Nigeria** | Kristen A. Stafford (UMB); Laura Steinhardt (CDC); Elsie Ilori (NCDC); Rosemary Audu (NIMR) (University of Maryland, Baltimore)[34] | Yes | Yes | Yes | Yes | Yes | Yes | Yes | Yes | Yes | Low |
| **Nigeria** | Kristen A. Stafford (UMB); Laura Steinhardt (CDC); Elsie Ilori (NCDC); Rosemary Audu (NIMR) (Nigeria Institute for Medical Research)[34] | Yes | Yes | Yes | Yes | Yes | Yes | Yes | Yes | Yes | Low |
| **Nigeria** | Olatunji Kolawole (Ministerial Expert Advisory Committee on COVID-19- Health Sector Response)[8] | No | Yes | Yes | Yes | Yes | Yes | Yes | No | Yes | Moderate |
| **Senegal** | Cheikh Talla (Institut Pasteur de Dakar)[9] | Yes | Yes | Yes | Yes | Unclear | Yes | Yes | Yes | Yes | Low |
| **Sierra Leone** | Mohamed Bailor Barrie (Harvard Medical School)[10] | Yes | Yes | Yes | Yes | Unclear | Yes | Yes | Yes | Unclear | Low |
| **South Africa** | Marvin Hsiao ( University of Cape Town)[50] | No | No | Yes | Yes | No | Yes | Yes | No | N/A | High |
| **South Africa** | Jaya George (National Health Laboratory Service)[37] | No | Yes | Yes | Yes | Unclear | Yes | Yes | Yes | Yes | Low |
| **South Africa** | Jaya George (National Health Laboratory Service)[36] | No | No | Yes | Yes | Yes | Yes | Yes | Yes | N/A | Moderate |
| **South Africa** | Nicole Wolter (National Health Laboratory Service)[11] | Yes | Yes | Yes | Yes | Yes | Yes | Yes | No | No | Moderate |
| **South Africa** | Marion Vermeulen (South African National Blood Service)[12] | No | No | Yes | No | Unclear | Yes | Yes | Yes | N/A | Moderate |
| **South Africa** | Jackie Kleynhans (National Institute for Communicable Diseases)[54] | Yes | Yes | Yes | Yes | No | Yes | Yes | No | Unclear | Moderate |
| **South Africa** | Jackie Kleynhans (National Institute for Communicable Diseases)[54] | Yes | Yes | Yes | Yes | No | Yes | Yes | No | Unclear | Moderate |
| **South Africa** | Jackie Kleynhans (National Institute for Communicable Diseases)[54] | Yes | Yes | Yes | Yes | No | Yes | Yes | No | Unclear | Moderate |
| **South Africa** | Jackie Kleynhans (National Institute for Communicable Diseases)[54] | Yes | Yes | Yes | Yes | No | Yes | Yes | No | Unclear | Moderate |
| **South Africa** | Jackie Kleynhans (National Institute for Communicable Diseases)[54] | Yes | Yes | Yes | Yes | No | Yes | Yes | No | Unclear | Moderate |
| **South Africa** | Jackie Kleynhans (National Institute for Communicable Diseases)[54] | Yes | Yes | Yes | Yes | No | Yes | Yes | No | Unclear | Moderate |
| **South Africa** | Jackie Kleynhans (National Institute for Communicable Diseases)[54] | Yes | Yes | Yes | Yes | No | Yes | Yes | No | Unclear | Moderate |
| **South Africa** | Jackie Kleynhans (National Institute for Communicable Diseases)[54] | Yes | Yes | Yes | Yes | No | Yes | Yes | No | Unclear | Moderate |
| **South Africa** | Jackie Kleynhans (National Institute for Communicable Diseases)[54] | Yes | Yes | Yes | Yes | No | Yes | Yes | No | Unclear | Moderate |
| **South Africa** | Jackie Kleynhans (National Institute for Communicable Diseases)[54] | Yes | Yes | Yes | Yes | No | Yes | Yes | No | Unclear | Moderate |
| **South Africa** | Jackie Kleynhans (National Institute for Communicable Diseases)[54] | Yes | Yes | Yes | Yes | No | Yes | Yes | No | Unclear | Moderate |
| **South Africa** | Jackie Kleynhans (National Institute for Communicable Diseases)[54] | Yes | Yes | Yes | Yes | No | Yes | Yes | No | Unclear | Moderate |
| **South Africa** | Jackie Kleynhans (National Institute for Communicable Diseases)[54] | Yes | Yes | Yes | Yes | No | Yes | Yes | No | Unclear | Moderate |
| **South Africa** | Jackie Kleynhans (National Institute for Communicable Diseases)[54] | Yes | Yes | Yes | Yes | No | Yes | Yes | No | Unclear | Moderate |
| **South Africa** | Jackie Kleynhans (National Institute for Communicable Diseases)[54] | Yes | Yes | Yes | Yes | No | Yes | Yes | No | Unclear | Moderate |
| **South Africa** | Jackie Kleynhans (National Institute for Communicable Diseases)[54] | Yes | Yes | Yes | Yes | No | Yes | Yes | No | Unclear | Moderate |
| **South Africa** | Lee Fairlie (Wits Reproductive Health and HIV Institute)[49] | No | No | Yes | Yes | Unclear | Yes | Yes | No | No | High |
| **South Sudan** | Kirsten E Wiens (World Health Organization)[38] | Yes | Yes | Yes | No | Unclear | Unclear | Yes | Yes | Unclear | Low |
| **Uganda** | Dr. Henry Kyobe Bosa (Ministry of Health/ UPDF)[55] | Yes | Yes | Yes | Yes | Yes | Yes | Yes | No | Unclear | Moderate |
| **Zambia** | Lloyd B Mulenga (Zambia Ministry of Health)[13] | Yes | Yes | Yes | Yes | Yes | Yes | Yes | Yes | Yes | Low |
| **Zambia** | Jonas Z Hines (US Centers for Disease Control and Prevention)[14] | Yes | No | Yes | No | No | Yes | Yes | No | Unclear | Moderate |
| **Zimbabwe** | Arun Fryatt (Biomedical Research and Training Institute)[39] | No | Yes | Yes | Yes | Yes | Unclear | Yes | No | Yes | Moderate |
| **Zimbabwe** | Arun Fryatt (Biomedical Research and Training Institute)[39] | No | Yes | Yes | Yes | Yes | Unclear | Yes | No | Yes | Moderate |
| **Egypt** | Samia Girgis (Ain-Shams University)[52] | No | No | Yes | Yes | Unclear | Yes | Yes | No | Unclear | High |
| **Egypt** | Sahar Abdelmaksoud (Ain-Shams University)[51] | No | Yes | No | No | No | Yes | Yes | No | N/A | High |

27	Mussagy Mahomed. Inquérito Sero-epidemiológico de SARS-CoV-2 na Cidade de Maxixe e Vila de Massinga (InCOVID 2020). República de Moçambique Ministério da Saúde 2020.

28	Arlete Mahumane. Inquérito Sero-epidemiológico de SARS-CoV-2 na Cidade de Chimoio (InCOVID 2020). República de Moçambique Ministério da Saúde 2020.

29	Jerónimo Langa. Inquérito Sero-epidemiológico de SARS-CoV-2 na Cidade de Lichinga (InCOVID 2020). República de Moçambique Ministério da Saúde 2020.

30	Mussagy Mahomed. Inquérito Sero-epidemiológico de SARS-CoV-2 na Cidade de Tete (InCOVID 2020). República de Moçambique Ministério da Saúde 2020.

31	Paulo Arnaldo. Inquérito Sero-epidemiológico de SARS-CoV-2 na Cidade de Pemba (InCOVID 2020). República de Moçambique Ministério da Saúde 2020.

32	Eduardo Samo Gudo. Inquérito Sero-epidemiológico de SARS-CoV-2 na Cidade de Nampula. República de Moçambique Ministério da Saúde 2020.

33	Population seroprevalence of SARS-CoV-2 antibodies in Anambra state, South-East, Nigeria. *International Journal of Infectious Diseases* Published Online First: July 2021. doi:[10.1016/j.ijid.2021.07.040](https://doi.org/10.1016/j.ijid.2021.07.040)

34	Ilori E, Stafford K, Steinhardt L, *et al.* Nigeria COVID-19 Household Seroprevalence Survey. 2021. doi:[10.5281/ZENODO.5644231](https://doi.org/10.5281/ZENODO.5644231)

35	Majiya H, Aliyu-Paiko M, Balogu VT, *et al.* Seroprevalence of COVID-19 in Niger State. *medRxiv* Published Online First: August 2020. doi:[10.1101/2020.08.04.20168112](https://doi.org/10.1101/2020.08.04.20168112)

36	George JA, Khoza S, Mayne E, *et al.* Sentinel seroprevalence of SARS-CoV-2 in Gauteng Province, South Africa, August - October 2020. *South African Medical Journal* 2021;**111**:1078–83. doi:[10.7196/SAMJ.2021.v111i11.15669](https://doi.org/10.7196/SAMJ.2021.v111i11.15669)

37	George JA, Khoza S, Mayne E, *et al.* Sentinel seroprevalence of SARS-CoV-2 in the Gauteng province, South Africa August to October 2020. *medRxiv* 2021;2021.04.27.21256099. doi:[10.1101/2021.04.27.21256099](https://doi.org/10.1101/2021.04.27.21256099)

38	Wiens KE, Mawien PN, Rumunu J, *et al.* Seroprevalence of Severe Acute Respiratory Syndrome Coronavirus 2 IgG in Juba, South Sudan, 2020. *Emerging Infectious Diseases* 2021;**27**. doi:[10.3201/eid2706.210568](https://doi.org/10.3201/eid2706.210568)

39	Fryatt A, Simms V, Bandason T, *et al.* Community SARS-CoV-2 seroprevalence before and after the second wave of SARS-CoV-2 infection in Harare, Zimbabwe. *EClinicalMedicine* 2021;**41**:101172. doi:[10.1016/j.eclinm.2021.101172](https://doi.org/10.1016/j.eclinm.2021.101172)

40	Sebastião CS, Galangue M, Gaston C, *et al.* Seroprevalence of anti-SARS-CoV-2 antibodies and risk factors among healthy blood donors in Luanda, Angola. *BMC Infectious Diseases* 2021;**21**:1131. doi:[10.1186/s12879-021-06814-0](https://doi.org/10.1186/s12879-021-06814-0)

41	Kempen JH, Abashawl A, Suga HK, *et al.* SARS-CoV-2 Serosurvey in Addis Ababa, Ethiopia. *The American Journal of Tropical Medicine and Hygiene* 2020;**103**:2022–3. doi:[10.4269/ajtmh.20-0816](https://doi.org/10.4269/ajtmh.20-0816)

42	Mveang Nzoghe A, Leboueny M, Kuissi Kamgaing E, *et al.* Circulating anti-SARS-CoV-2 nucleocapsid (N)-protein antibodies and anti-SARS-CoV-2 spike (S)-protein antibodies in an African setting: Herd immunity, not there yet! *BMC Research Notes* 2021;**14**:152. doi:[10.1186/s13104-021-05570-3](https://doi.org/10.1186/s13104-021-05570-3)

43	Nyagwange J, Kutima B, Mwai K, *et al.* Comparative performance of WANTAI ELISA for total immunoglobulin to receptor binding protein and an ELISA for IgG to spike protein in detecting SARS-CoV-2 antibodies in Kenyan populations. *Journal of Clinical Virology* 2022;**146**:105061. doi:[10.1016/j.jcv.2021.105061](https://doi.org/10.1016/j.jcv.2021.105061)

44	Lucinde R, Mugo D, Bottomley C, *et al.* Sero-surveillance for IgG to SARS-CoV-2 at antenatal care clinics in two Kenyan referral hospitals. *medRxiv* 2021;2021.02.05.21250735. doi:[10.1101/2021.02.05.21250735](https://doi.org/10.1101/2021.02.05.21250735)

45	Crowell TA, Daud II, Maswai J, *et al.* SARS-CoV-2 antibody prevalence in people with and without HIV in rural western Kenya, January to March 2020. *AIDS* Published Online First: September 2021. doi:[10.1097/QAD.0000000000003054](https://doi.org/10.1097/QAD.0000000000003054)

46	Razafimahatratra SL, Ndiaye MDB, Rasoloharimanana LT, *et al.* Seroprevalence of ancestral and Beta SARS-CoV-2 antibodies in Malagasy blood donors. *The Lancet Global Health* 2021;**9**:e1363–4. doi:[10.1016/S2214-109X(21)00361-2](https://doi.org/10.1016/S2214-109X(21)00361-2)

47	Schoenhals M, Rabenindrina N, Rakotondramanga JM, *et al.* SARS-CoV-2 antibody seroprevalence follow-up in Malagasy blood donors during the 2020 COVID-19 Epidemic. *EBioMedicine* 2021;**68**:103419. doi:[10.1016/j.ebiom.2021.103419](https://doi.org/10.1016/j.ebiom.2021.103419)

48	Sero-pravelence of SARS CoV-2 IgM and IgG Antibodies Amongst Blood Donors in Nigeria. Published Online First: January 2021. doi:[10.21203/rs.3.rs-151037/v1](https://doi.org/10.21203/rs.3.rs-151037/v1)

49	Sawry S, le Roux JG, Mbatha P, *et al.* High prevalence of SARS-CoV-2 antibodies in pregnant women in the inner city of Johannesburg, Gauteng Province, South Africa. 2021. doi:[10.5281/ZENODO.5643228](https://doi.org/10.5281/ZENODO.5643228)

50	Hsiao M, Davies M-A, Kalk E, *et al.* SARS-CoV-2 seroprevalence in the Cape Town Metropolitan sub-districts after the peak of infections. *NICD COVID-19 Special Public Health Surveillance Bulletin* 2020;**18**:9.

51	Abdelmaksoud S. Seropositive coronavirus disease 2019 detection among apparently healthy voluntary blood donors: A preliminary step to catch asymptomatic cases as a potential viral spread route. 2021;**46**:65–9. doi:[10.4103/ejh.ejh_59_20](https://doi.org/10.4103/ejh.ejh_59_20)

52	Girgis SA, Hafez HM, Elarab HE, *et al.* SARS-CoV-2 PCR positivity rate and seroprevalence of related antibodies among a sample of patients in Cairo: Pre-wave 2 results of a screening program in a university hospital. *PLOS ONE* 2021;**16**:e0254581. doi:[10.1371/journal.pone.0254581](https://doi.org/10.1371/journal.pone.0254581)

53	Manirakiza A, Malaka C, Yambiyo BM, *et al.* Seroprevalence of anti-SARS-CoV-2 antibodies among communities in Bangui, Central African Republic. 2021. doi:[10.5281/zenodo.5794867](https://doi.org/10.5281/zenodo.5794867)

54	Cohen, Cheryl, Kleynhans, Jackie, von Gottberg, Anne, *et al.* SARS-CoV-2 Seroprevalence in a Rural and Urban Household Cohort, South Africa, July 2020–September 2021. 2022. doi:[10.5281/ZENODO.5898414](https://doi.org/10.5281/ZENODO.5898414)

55	Bosa, Henry Kyobe, Ario, Alex R., Wamala, Robert. Population-based, Age- and Gender- Stratified Sero-Survey Study for SARS-CoV-2 in Uganda. 2022. doi:[10.5281/zenodo.5961443](https://doi.org/10.5281/zenodo.5961443)
